# Health care use attributable to COVID-19: A propensity matched national electronic health records cohort study of 249,390 people in Wales, UK

**DOI:** 10.1101/2022.04.21.22274152

**Authors:** J Kennedy, M Parker, M Seaborne, M Mhereeg, A Walker, V Walker, S Denaxas, N Kennedy, S.V Katikireddi, S Brophy

## Abstract

**Background:** To determine the extent and nature of changes in infected patients healthcare utilization, we studied healthcare contact in the 1-4 weeks and 5-24 weeks following a COVID-19 diagnosis compared to propensity matched controls.

**Methods:** Survival analysis was used for time to death and first clinical outcomes including clinical terminology concepts for post-viral illness, fatigue, embolism, respiratory conditions, mental and developmental conditions, fit note, or hospital attendance. Increased instantaneous risk for the occurrence of an outcome for positive individuals was quantified using hazard ratios (HR) from Cox Regression and absolute risk was quantified using relative risk (RR) from life table analysis.

**Results:** Compared to matched individuals testing negative, surviving positive community-tested patients had a higher risk of post-viral illness (HR: 4.57, 95%CI: 1.77-11.80, p=0.002), fatigue (HR: 1.47, 95%CI: 1.24-1.75, p<0.001) and embolism (HR: 1.51, 95%CI: 1.13-2.02, p=0.005) at 5-24 weeks post-diagnosis. In the four weeks after COVID-19 higher rates of sick notes were being issued for community-tested (HR: 3.04, 95%CI: 0.88 to 10.50, p<0.079); the risk was reduced after four weeks, compared to controls. Overall healthcare attendance for anxiety, depression was less likely in those with COVID-19 in the first four weeks (HR: 0.83, 95%CI: 0.73-1.06, p=0.007). After four weeks, anxiety, depression is less likely to occur for the positive community-tested individuals (HR: 0.87, 95%CI: 0.77-1.00, p=0.048), but more likely for positive hospital-tested individuals (HR: 1.16, 95%CI: 1.00-1.45, p=0.053). Although statistical associations between positive infection and post-infection healthcare use are clear, the absolute use of healthcare is very.

**Conclusions:** Community COVID-19 disease is associated with increased risks of post-viral illness, fatigue, embolism, depression, anxiety and respiratory conditions. Despite these elevated risks, the absolute healthcare burden is low. Either very small proportions of people experience adverse outcomes following COVID-19 or they are not presenting to healthcare.

**Trial registration:** Data held in SAIL databank are anonymised and therefore, no ethical approval is required. All data in SAIL has the permission from the relevant Caldicott Guardian or Data Protection Officer and SAIL-related projects are required to obtain Information Governance Review Panel (IGRP) approval. The IGRP approval number for this study is 1259.

## Introduction

Considerable concerns exist about chronic, debilitating, and varied symptoms experienced by people who have had coronavirus disease (COVID-19) caused by severe acute respiratory syndrome coronavirus 2 (SARS-CoV-2) infection (1). However, the natural history of morbidity and healthcare use after infection and disease remains unclear. While most people who experience COVID-19 recover quickly, an unknown minority experience prolonged symptoms that manifest as a range of post-COVID-19 illness (1).

As the pandemic continues, meeting the needs of the increasing numbers of people who have recovered from COVID-19 remains important. Recovery from any severe disease can often be protracted. However, there is increasing evidence that even those who were not hospitalised with COVID-19 infection, may have longer-term health consequences, such as chronic fatigue and respiratory issues (1). Hence, a substantial number of people are experiencing persistent symptoms such as pain, heart palpitations, breathlessness, cognitive impairment, and fatigue (2). Many symptoms have been reported following COVID-19 (3), with evidence from a symptom tracker in the UK suggesting the existence of six different syndromes. However, consensus on what clusters of sequelae exist is not available. The highly varied nature of symptoms and experiences reported by patients has made standardised diagnosis difficult. In particular, accurate clinical coding of ‘Long COVID’ has been lacking, thereby impeding research efforts (4). However, age, self-reported health status before the onset of symptoms, self-reported pre-existing comorbidities and the number of symptoms during the infection were found to significantly predict the number of symptoms patients with long COVID may experience at follow-up (5). The most common symptoms presented to the GP, four weeks after infection, were joint pain (2.5%), anxiety (1.2%) and prescription of non-steroidal anti-inflammatory drugs (1.2%) using routine medical record data (6).

Health systems internationally have been under extreme pressure due to the COVID-19 pandemic. Many countries have faced large demands for healthcare, resulting in elective care being postponed and many patients foregoing or delaying necessary healthcare. These stresses have led to large waiting lists in the UK. The large numbers of SARS-CoV-2 infections could lead to a further demand on healthcare because of long COVID. At present, there is limited data (6) available to inform health systems about the scale of demand that might be expected and what services might be sought. However, establishing the extent to which these conditions are attributable to COVID-19 or reflect disease burden among the general population can be difficult. Misclassification may also occur because of the general misunderstanding of long-term consequences of COVID-19 and the likelihood that clinicians may attribute unrelated illness, or worsening of existing symptoms, to COVID-19.

Research to establish the natural history of COVID-19 disease over the medium- and long-term can inform understanding of the long-term effect of COVID and potentially inform expectations about future health system demands. This study therefore aims to develop an understanding of the burden on the healthcare system attributable to COVID-19, quantify the length of time of excess resource use, and identify the different diagnostic clusters that underpin any excess healthcare use.

## Methods

### Study design

To investigate primary and secondary healthcare use after a positive reverse transcription – polymerase chain reaction (RT-PCR) test result, we followed a target trial (also called an ‘emulated trial’) design (7,8), e.g. emulating a randomised control trial using observational data, by ensuring cases and controls are as similar as possible at baseline and only the exposure (e.g COVID positive/negative) differs between the groups. In addition, given the consequences of COVID-19 may differ depending on the severity of the initial disease, we defined two different exposed groups, reflecting different severity of disease: a) Received a positive RT-PCR at a community testing site; and b) received a positive RT-PCR at a hospital testing site (See appendix A for what constitutes a ‘community’ and ‘hospital’ testing site). For each exposed group, we defined two alternative control groups. First, we followed a test negative design (i.e., comparing people who tested positive to those testing negative) to better account for potential under-ascertainment and variable testing. We then compared people who tested negative to controls who had not yet been tested at that time. This latter analysis provided greater statistical power for analysis but was potentially at greater risk of bias due to differential testing. For example, those with any respiratory symptoms would have a COVID-19 test so the non-tested group were predominantly non-symptomatic patients.

### Study population (28/02/2020 to 26/08/2021)

The study used the Secure Anonymised Information Linkage (SAIL) Databank in Wales (9), which includes nation-wide electronic health records from primary and secondary care. The SAIL databank is a data repository which allows person-based data linkage across datasets. This databank includes Welsh GP data, hospital in- and out-patient records, as well as mortality data collected by the Office of National Statistics (ONS). SAIL holds over a billion anonymised records and has Welsh population coverage for 100% of hospital data and 90% of GPs. It uses a split-file approach to ensure anonymisation and overcome issues of confidentiality and disclosure in health-related data warehousing. Demographic data are sent to a partner organisation, NHS Wales Informatics Service, where identifiable information is removed; clinical data are sent directly to the SAIL Databank and an individual is assigned an encrypted anonymised linking field (ALF). The ALF is used to link anonymised individuals across datasets, facilitating longitudinal analysis of an individual’s journey through multiple health, education and social datasets (9).

The data linked in this study (Figure 1) were; Welsh Demographic Service to identify all patients registered with a GP practice and identify when people move in and out of Wales, Primary care GP dataset to identify healthcare contacts in general practice, data collected by GPs are captured via Read Codes version 2 (5-character alphanumeric codes related to diagnosis, medication and process of care codes) (10), the hospital inpatient and outpatient data collected in the Patient Episode Database for Wales, which contains clinical information regarding patients’ hospital admissions, discharges, diagnoses and operations using the International Classification of Diseases (ICD-10) clinical classification system. The ONS Mortality dataset contains demographic data, place of death and underlying cause of death (also ICD-10) and test results from the laboratory management information system to identify individuals who have had a laboratory COVID-19 test and the test result.

**Figure 1:**
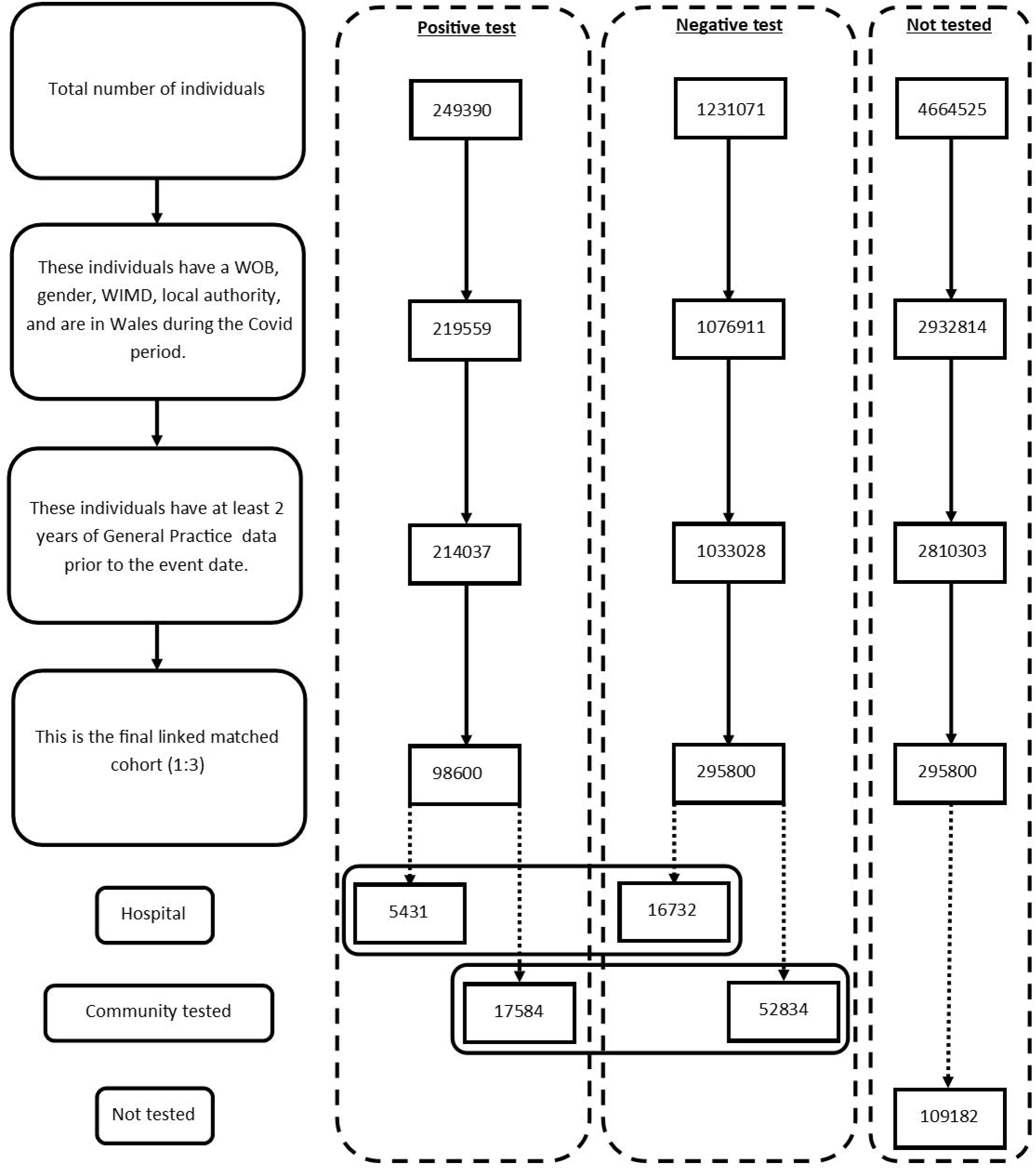
Flow Diagram of participants.

### Identifying SARS-CoV-2

The study identified all people in Wales registered with a Welsh GP and stratified them as having had a SARS-CoV-2 positive test result, a negative result, or no SARS-CoV-2 test between 28/02/2020 and 26/08/2021. Two exposed study cohorts were defined: A) Tested positive for SARS-CoV-2 in the community and B) Tested positive for SARS-CoV-2 in hospital, based on the location data associated with their testing result. Two sets of controls were created through propensity score matching (see below) for each of these exposure groups: C) Controls who tested negative for SARS-CoV-2 (individuals that have negative tests prior to being positive are valid for linking to other individuals which are positive), and D) Controls who had not tested positive for COVID at the index time (but may test positive later and become a case or test negative and enter group A at a later date). Study cohorts identified included: A) Exposed (Case), those tested positive for SARS-CoV-2 when in hospital or in the community B) Controls A who tested negative for SARS-CoV-2 in hospital or the community (individuals that have negative tests prior to being positive are valid for linking to other individuals which are positive) and, C) Control group B, those who had not been tested for COVID at the index time (but may test later and become cases or enter group A at a later date).

### Propensity matching

Exposure to SARS-CoV-2 infection is defined as starting from the date of having a positive RT-PCR test result. A positive hospital case was matched with three controls for control A (test negative) also tested in hospital and three controls from control B (not tested at the index date). A positive community case was matched with three controls for control A tested in the community and control B (not tested). Follow-up of the exposure period was censored for the control participant if the control goes on to have a positive test. Controls who have a positive test were then eligible to be included in the analysis within the exposed group. Cases and controls were propensity matched on; Welsh Index of multiple deprivation quintile (WIMD), comorbidities using the Charlson Comorbidity Index (CCI), number of people in the household, number of previous SARS-CoV-2 tests. In addition to the propensity score the following features were utilised to make the control group; gender, local authority area, week of birth (+/- 1 year from date), and location of the test (community/hospital/other).

### Data cleaning

Data were checked for patterns of missingness and implausible values for all analytical variables investigated. A record of reasons for exclusion from analysis was maintained. Individuals with no recorded test location (excluding the untested population) were excluded from the analysis.

### Outcomes

The primary outcome was to determine whether testing positive for SARS-CoV-2 results in different use of primary and secondary care in the first 6 months following the test, compared to those who had currently not tested positive.

### Statistical analysis

Descriptive statistics were gathered on the negative test, positive test, and untested populations to assess the adequacy of the propensity matching. Frequency of deaths, primary and secondary care visits were tracked each month and adjusted for the population size accounting for individuals who had been censored. General groups of codes used to define the clinical outcomes of this study can be seen in appendix B (full list in appendix E). Sick notes only refer to those issued by the GP and not self-certified notes. Whether the codes originated in primary or secondary and additional notes are also included in this appendix (E). Outcomes include death, first secondary care visit, diabetes, embolism, fatigue, mental and behavioural disorders, respiratory conditions, post viral illness and sick notes. Alternatively, an individual could be censored. Reasons for censorship include death (from any cause), end of follow-up period (28 days or 168 days), end of study period (01/08/2021), changing COVID-19 test status through a confirmed RT-PCR test or leaving Wales.

Secondary analyses examined a) health care use b) the length of time by which the excess risk associated with SARS-CoV-2 infection and COVID-19 disease has ended.

### Survival Analysis

Survival analysis was used to examine the time between an individual’s first RT-PCR and the first occurrence of an outcome or endpoint. The time between the index date and the endpoint was calculated for each different outcome independently. Age has been calculated using the week of birth and the date of first test or index date divided by 365.25 to provide the age in years.

Cox proportional hazard models were used to produce hazard ratios (HR) to quantify likelihood the first instantaneous occurrence of an outcome within: i.0 - 28 days (1 month), ii. 29 - 168 days (5 months) following a RT-PCR. The negative test population result was set as the reference group for the positive and untested population. The untested population was compared with the negative group to understand bias in who attends for testing as those who are untested are also unlikely to attend for healthcare for other conditions. The negative test population was also set as the reference group when the data are stratified by test location (community or hospital). Follow up starts on the day of being classified as exposed (or the date of being matched for controls). Follow-up ends on the first of: experiencing the outcome of interest or being censored.

Individual models were run for each outcome, time frame (1-4 weeks & 5-24 weeks) and location (combined location, hospital only and community only). Dataset conditions were dependent on the time frame being studied: i.1-4 weeks: the end of the follow up period was 28 days from the index date, ii.5 - 24 weeks: the follow up period was between 29 and 168 days from the index date. If an RT-PCR positive individual died within the first 28 days all their propensity matched partners were also removed from the analysis. Life table analysis uses the full follow up period between day 0 and 168 from the first test or index date.

### Life Table Analysis

Risk ratios (RR) showing the relative risk of an outcome every 4 weeks compared to a reference group were calculated through life table analysis. The analysis creates a ratio of absolute risk (AR) for each outcome adjusting the population size as individuals are censored. The reference groups for those tested in the community and hospital settings are negative tests in their respective environments. The reference group for the untested population was negative tests in the community only as this was to explore the bias in health care use for those who have not been tested.

### Software

The data handling and preparation for survival analysis, descriptive statistics and life table analysis were performed in an SQL database (SAIL) using Eclipse (11) and tabulated in Microsoft Excel for database extraction. Final data preparations specific to survival analysis were performed in RStudio 2021.09.0 such as setting reference groups for the Cox proportional hazard models (12). Survival analysis was performed in R studio using packages ‘Survminer’ (13) and ‘Survival’ (14). ‘Love Plots’ (appendix C) were created in R using the package ‘cobalt’ (15). Risk ratio and confidence intervals calculations were performed in Microsoft Excel (Version 2201.) and hazard ratio plots (figures 5 - 8) were also manually constructed in Microsoft Excel.

### Ethical approval

Data held in the SAIL databank are anonymised and therefore, no NHS ethical approval is required. All data contained in SAIL has the permission from the relevant Caldicott Guardian or Data Protection Officer and SAIL-related projects are required to obtain Information Governance Review Panel (IGRP) approval. The IGRP approval number for this study is 1259.

## Results

### Demographic of Case Controls

There were 249,390 people who had a positive SARS-CoV-2 test between 28/02/2020 and 26/08/2021. When these were propensity matched with controls this number reduced to 41,838 individuals, thus removing 83% of the data. The dataset was then further restricted by removing all matches for whom their test location was matched as missing.

Three matched cohorts are used in this study; COVID-19 test positive (case), COVID-19 test negative (control) and not tested (control). 23,015 and 69,566 individuals were identified to have had a positive and negative test respectively. Appendix C shows ‘Love Plots’ for the standardised mean distribution before and after the propensity matching had occurred. Censorship patterns were checked and were similar across the cohorts.

### Death Following RT-PCR

The mortality rate was higher within the first four weeks of those who tested positive (n = 3019.77) compared with individuals who tested negative (n = 980.36) or who were untested (n = 128.23) (see figure 2). The untested population maintains a low mortality frequency for the entire follow up period (77.29 - 140.55). The mortality frequency in positive and negative controls converges at 5-8 weeks and declines towards the end of the follow-up period. The negative population had a marginally higher frequency during the 5–24-week period.

**Figure 2.**
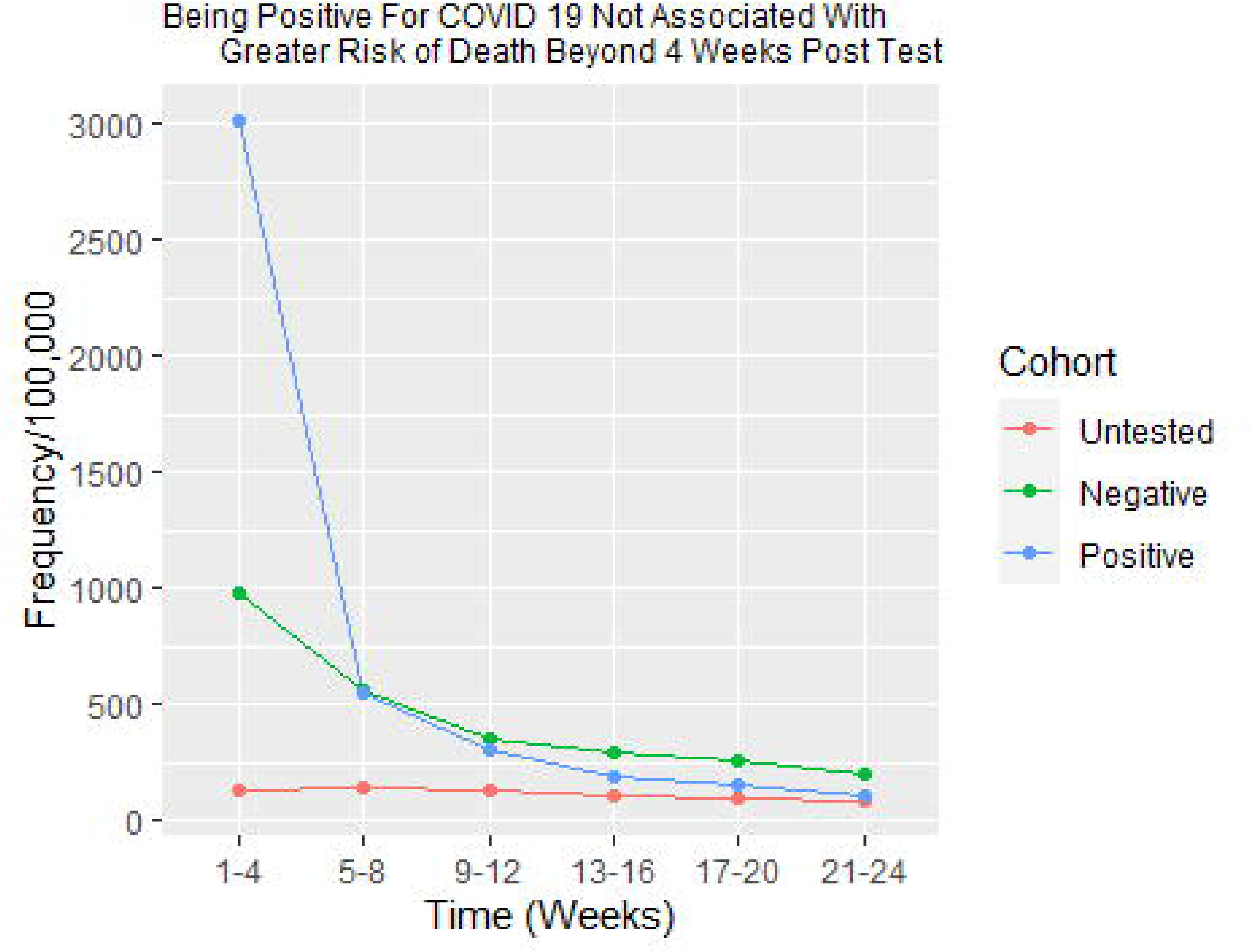
Deaths Per 100,000 Every 4 Weeks Following a COVID-19 Test or the Index Date (Untested)

These findings are also observed in the cox proportional hazard models (see figures 5 - 8). Positive individuals were significantly more likely to die within 28 days of a positive test in both the community (HR: 4.10, 95%CI: 3.38 to 4.97, p<0.001) and hospital (HR: 2.83, 95%CI: 2.50 to 3.22, p<0.001) environments when compared controls. Untested individuals were significantly less likely to die (HR: 0.13, 95%CI: 0.11 to 0.16, p<0.001) compared with tested. After 5 weeks, positive individuals were significantly less likely to die if tested in the community (HR: 0.72, 95%CI: 0.58 to 0.91, p=0.005) more likely to die if tested in hospital (HR: 1.22, 95%CI: 1.01 to 1.47, p=0.037. Untested individuals were much less likely to die than those tested after 5 weeks (HR: 0.36, 95%CI: 0.32 to 0.41, p<0.001)

Risk ratios from the life table analysis support the observed trends and can be seen in table 2. They illustrate that individuals are more likely to die following a positive test in both the hospital (RR: 2.77, 95%CI: 2.45 to 3.14) and community (RR: 4.06, 95%CI: 3.35 to 4.92) environment in the first four weeks. After 5 weeks an individual was less likely to die if receiving a positive test in the community or if no test had been taken by this point.

**Table 1:** Demographic profile of propensity matched infected and non-infected individuals analysed.

Survival Probability Time to Event
|  | Case (+ ve test) |  | Control (- ve test) |  | Control |
| --- | --- | --- | --- | --- | --- |
|  | Community | Hospital | Community | Hospital | Untested |
| Total, n (%) | 17,584 (24.97) | 5,431 (24.50) | 52,834 (75.03) | 16,732 (75.50) | 109,182<br>(100.00) |
| Males, n (%) | 8,222 (46.76) | 2,770 (51.00) | 24,856 (47.05) | 8,361 (49.97) | 54,435<br>(49.86) |
| WIMD Score 1 or<br>2, n (%) | 3,945 (22.44) | 1,233 (22.70) | 11,862 (22.45) | 3,441 (20.56) | 21,694<br>(19.87) |
| Mean Age (Years<br>(Std.D)) | 42.56 (23.47) | 51.90 (25.83) | 42.74 (23.56) | 50.66 (22.92) | 43.17<br>(23.95) |
| Mean Charlson<br>Index (Std.D) | 0.60 (1.00) | 1.13 (1.53) | 0.55 (1.01) | 1.03 (1.47) | 0.53 (0.89) |

**Table 2.**
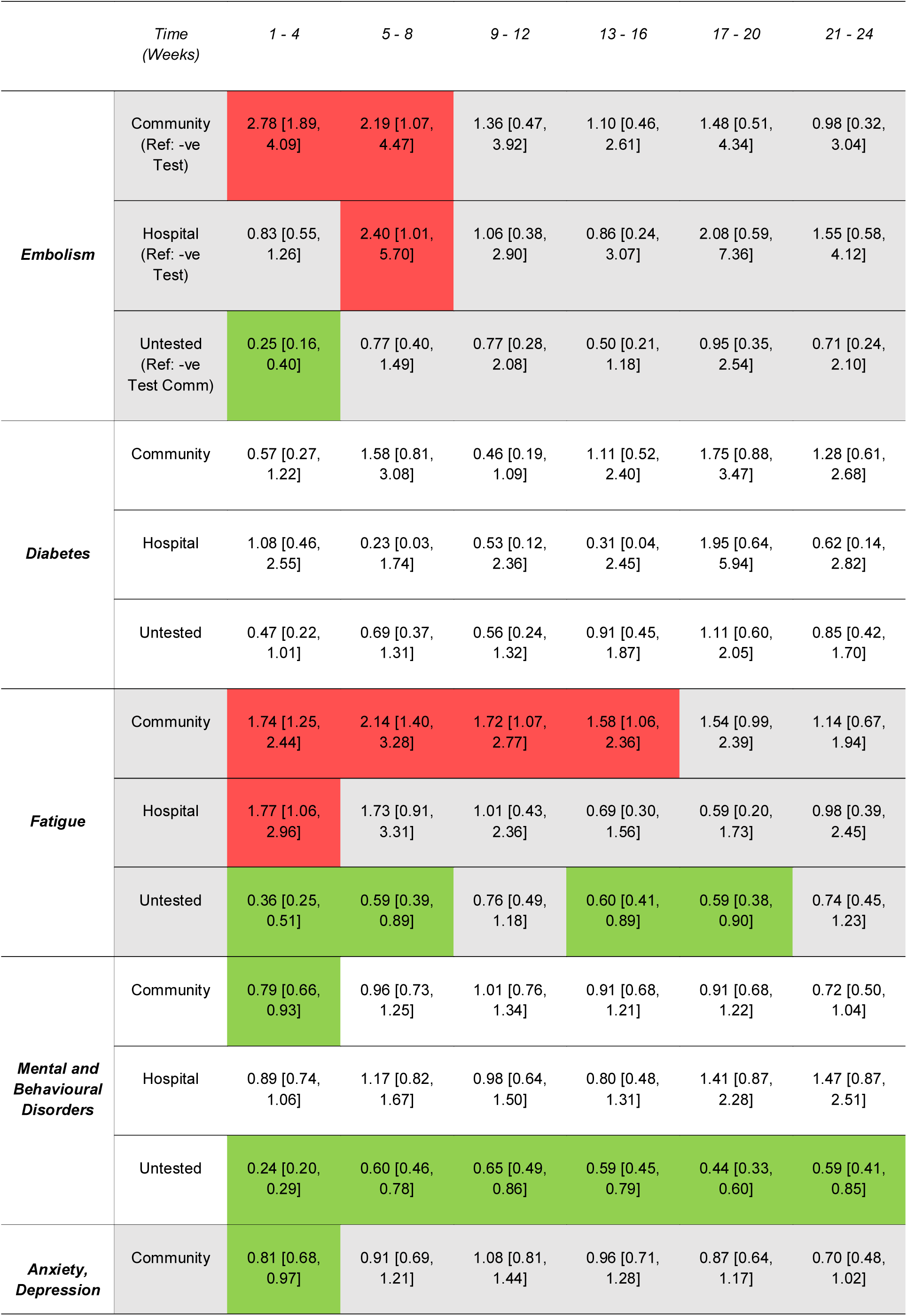
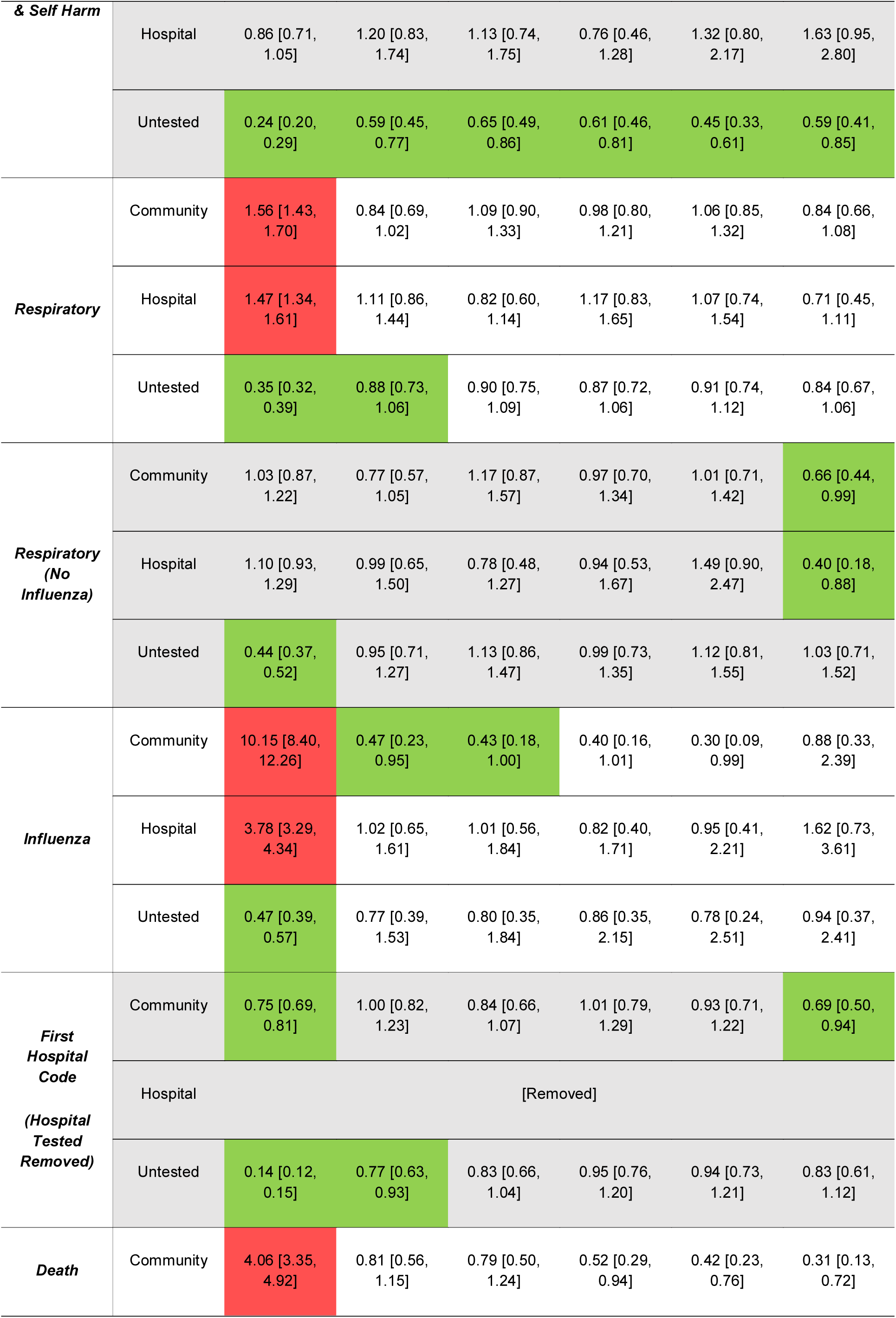
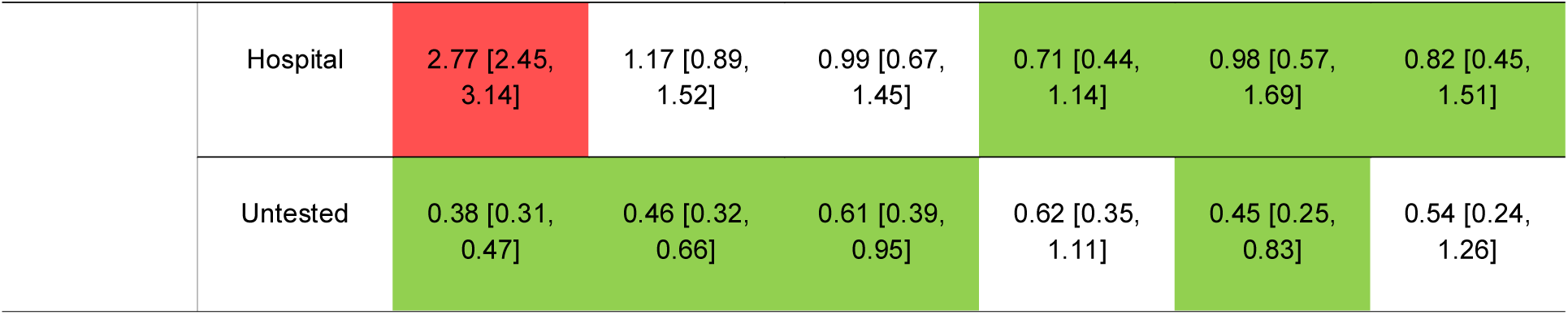
The risk ratios [95% confidence intervals] from life table analysis are presented and show the relative risk of a first event each month following a positive covid test or from the index date (Untested). Reference groups are stated in the first row. Sick notes and post viral syndrome removed due to insufficient data. **[Key: Green: =<0.99, Red >= 1.01 & 95%CI Does not cross 1.00 threshold].**

#### Outcomes 1 - 4 - and 5 - 24-weeks Post Test

##### Combined Environments

The hazard ratios, confidence intervals and p-values for the occurrence of each outcome can be seen in figures 3-6. For both environments combined (figure 3), diagnosis of influenza (HR: 5.68, 5.08 to 6.43, p<0.001), fatigue (HR 1.77, 1.34 to 2.35, p<0.001), respiratory condition (HR: 1.53, 1.43 to 1.63, p<0.001), embolism (HR: 1.50, 1.15 to 1.97, p<0.003), requirement for a sick note (HR: 3.58, 95%CI:1.20 to 10.07, p = 0.022) and death (HR: 3.12, 95%CI:2.80 to 3.46, p<0.001) were higher in the first 4 weeks compared to their propensity matched controls. In the 5–24-week period, previously positive individuals had higher post-viral syndrome (HR: 4.57, 95%CI:1.77 to 11.08, p=0.002), embolism (HR: 1.51, 95%CI:1.13 to 2.02, p=0.005) and fatigue (HR: 1.47, 1.24 to 1.75, p<0.001). In addition, when compared to negative controls, survivors of COVID-19 were less likely to be diagnosed with anxiety, depression, or other mental or developmental condition codes.

**Figure 3.**
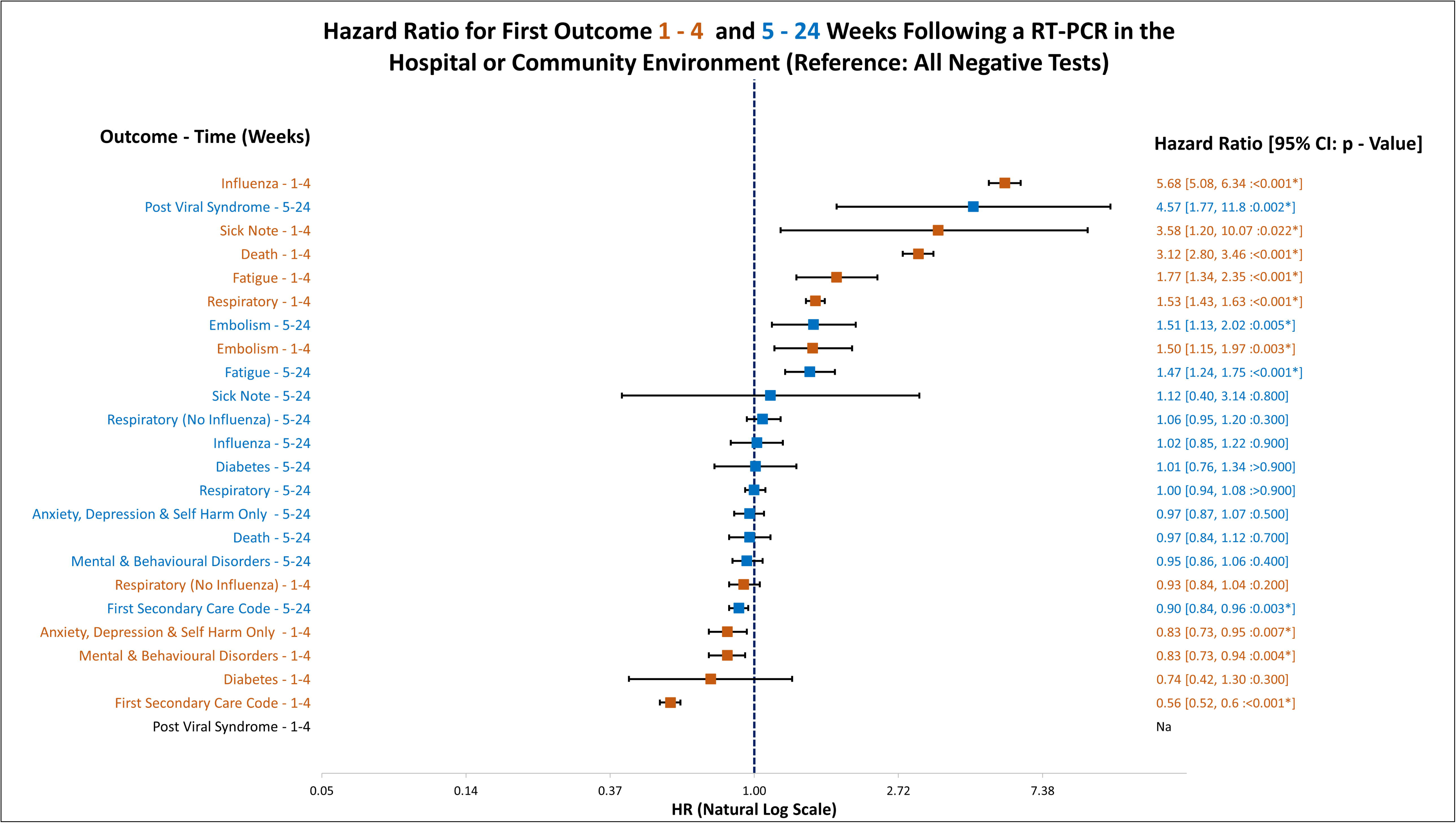
Displays the hazard ratio for the altered chance of an outcome if an individual had a positive COVID-19 test in either the hospital or community environment. The reference group is any negative COVID-19 test result in either environment.

#### Separate Test Environments

Hazard ratios for community and hospital tested individuals can be seen in figures 4 and 5. The use of the influenza codes were much higher in those diagnosed in the community (HR: 10.31, 95%CI: 8.53 to 12.50, p<0.001) compared to the hospital diagnosed (HR: 3.92, 95%CI: 3.40 to 4.51, p<0.001) in the first 4 weeks. Sick notes were more likely to be given following a positive test in hospital (HR: 3.04, 95%CI: 0.88 to 10.50, p=0.079) and community sites (HR: 6.43, 95%CI: 0.58 to 70.90, p=0.130). Fatigue occurred with a similar frequency 1-4 weeks following tests at either location Hospital (HR: 1.81, 95%CI:1.08 to 3.02, p = 0.024) and community (HR: 1.76, 95%CI: 1.26 to 2.46, <0.001). Embolism codes occurred more frequently within 1-4 weeks in the community (HR: 2.83, 95%CI: 1.91 to 4.21, p<0.001) vs Hospital (HR: 0.88, 95%CI: 0.55 to 1.27, p = 0.440).

**Figure 4.**
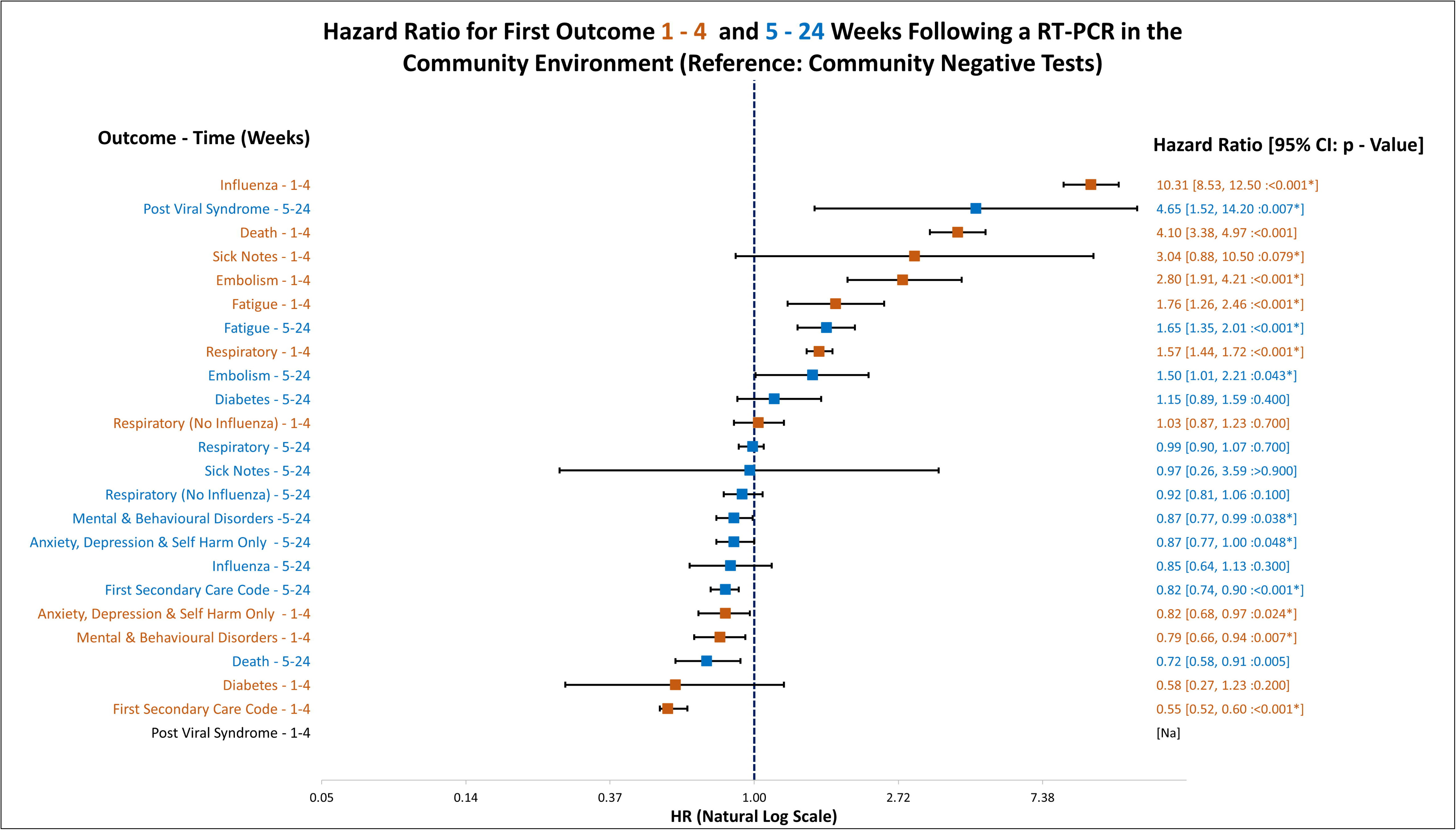
Displays the hazard ratio for the altered chance of an outcome if an individual had a positive COVID-19 test in the community environment only. The reference group is negative COVID-19 tests in the community environment only.

**Figure 5.**
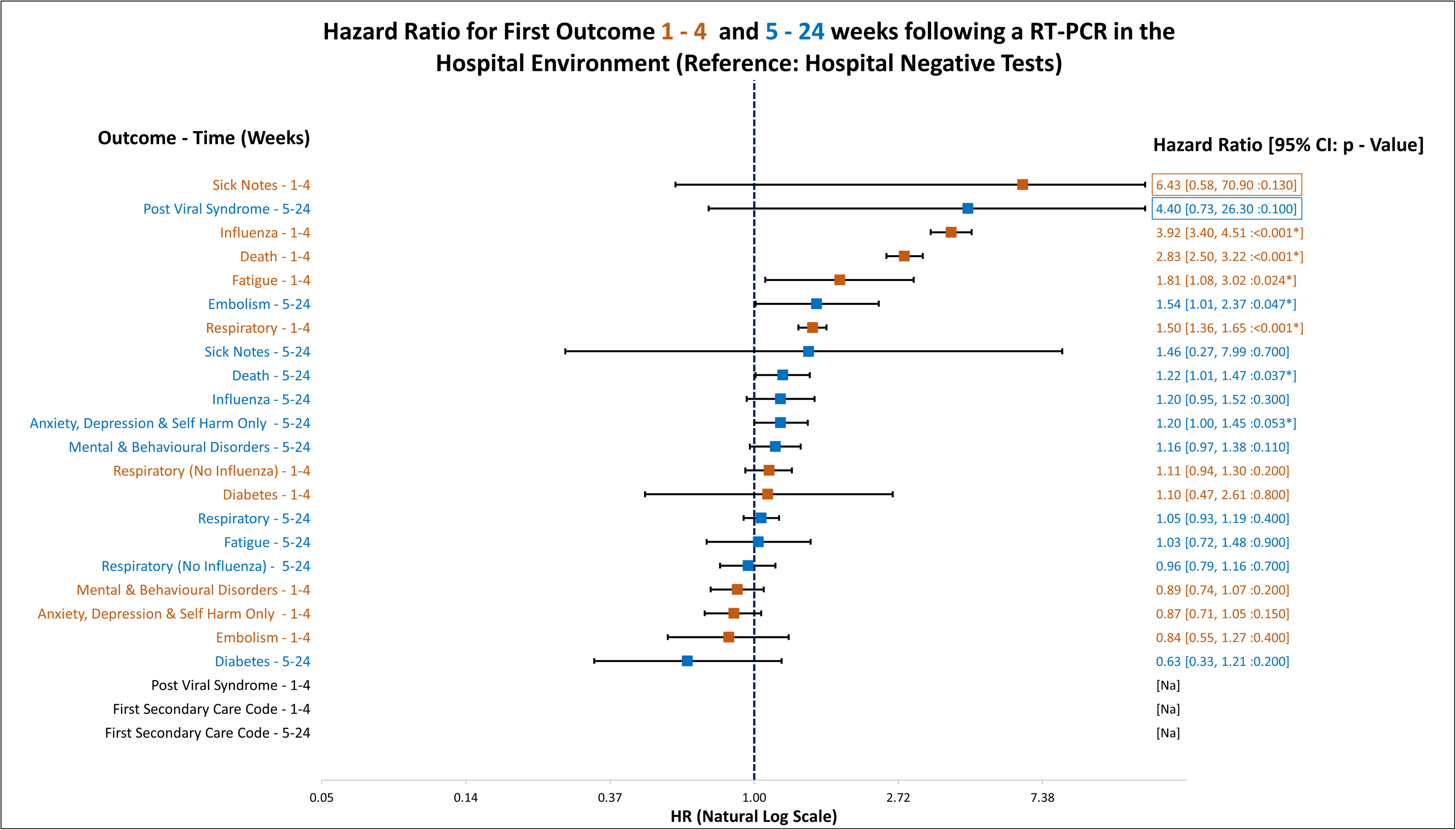
Displays the hazard ratio for the altered chance of an outcome if an individual had a positive COVID-19 test in the hospital environment. The reference group is negative COVID-19 tests in the hospital environment only. Hazard ratios in boxes exceed the x-axis, and the positive value has been capped at a value of 15.

Individuals were more at risk of post-viral syndrome following a test in the community (HR:4.65, 95%CI: 1.52 to 14.20, p<0.007) and hospital (HR: 4.40, 95%CI: 0.73 to 26.30, p<0.100). Regardless of the environment, individuals who had tested positive were more likely to have an embolism code 5-24 weeks following a test (HR: 1.50, 95%CI:1.01 to 2.21, p = 0.043) (community) and (HR:1.54, 95%CI 1.01 to 2.37, p = 0.047) (hospital). Survivors of COVID tested in the community appeared to be at a lower risk from anxiety, depression and self–harm (HR: 0.87, 95%CI: 0.77 to 1.00, p = 0.048), however those tested in the hospital were at an increased risk (HR: 1.20, 95%CI: 1.00 to 1.45, p = 0.053).

### Untested Controls

Hazard ratios for the untested controls comparison to test negative individuals can be seen in figure 6. The people who did not have any record of a COVID test between 28/02/2020 and 26/08/2021 were at a lower risk of all outcomes.

**Figure 6.**
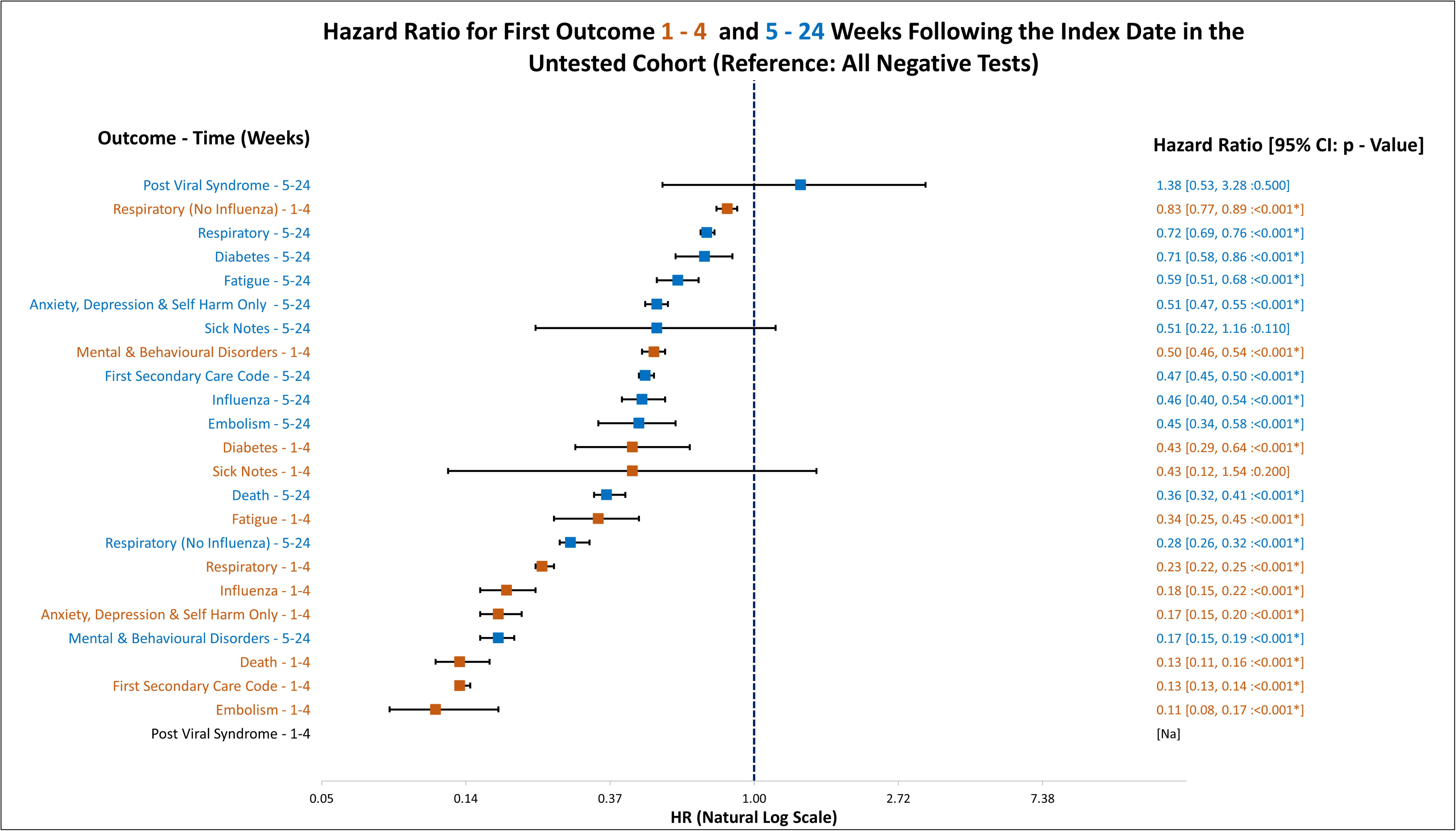
Displays the hazard ratio for the altered chance of an outcome if an individual has not had a COVID-19 test. This comparison was used to examine possible bias in the tested population compared to the untested.

### Fatigue

Fatigue was higher for the entire follow-up period for those who had tested positive. Community rates were comparable to those in hospital. Those who had been in hospital (but tested negative) had higher levels of fatigue than people in the community who were untested (see figure 7).

**Figure 7:**
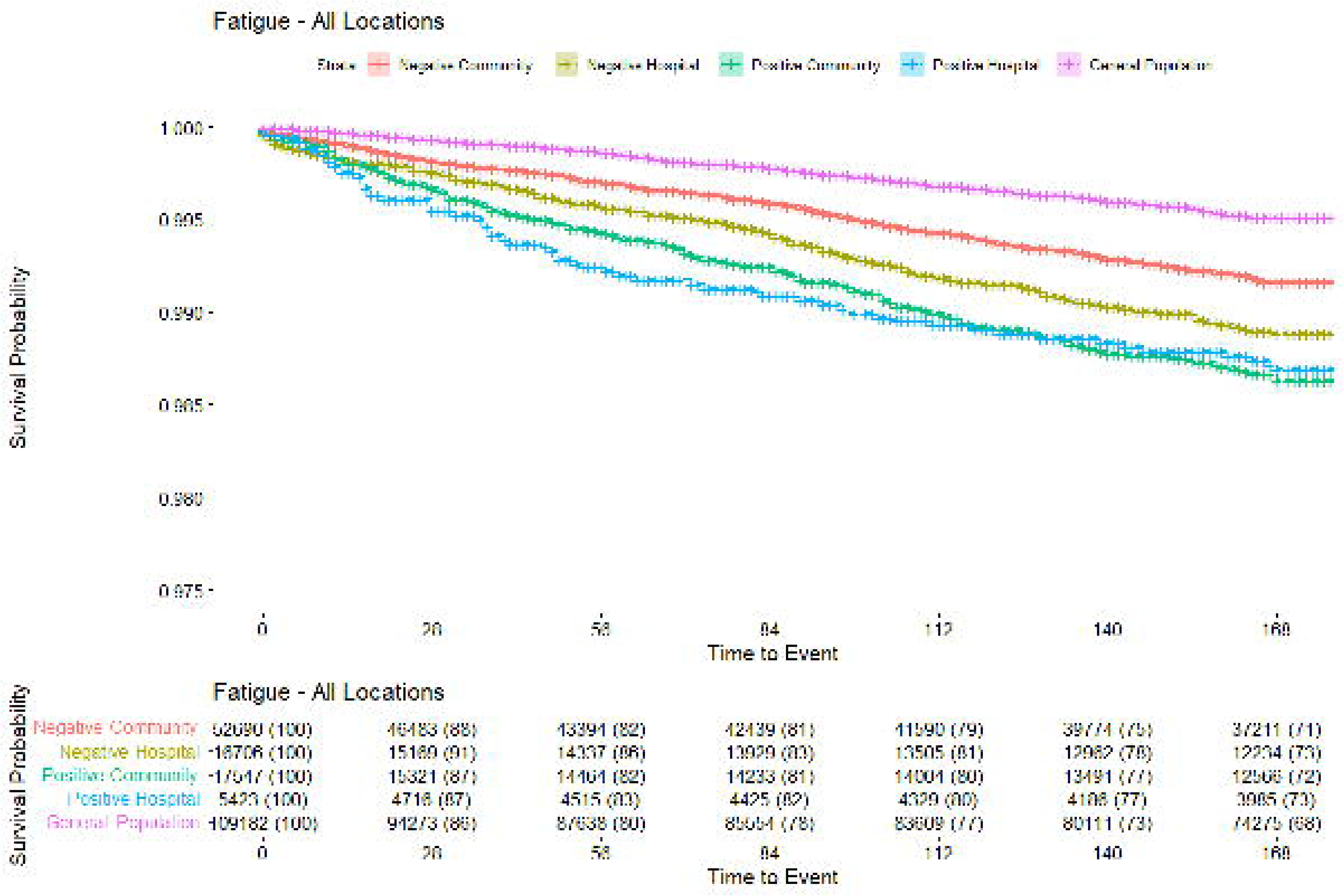
Time to first fatigue code by COVID diagnosis.

### Embolism

Across all groups, the overall volume of events is very low. The risk of embolism (figure 8) is much higher in those who were in hospital. The rates are equivalent in those testing either positive or negative. There appears to be a greater volume of events within the first 3 weeks for those who were hospital negative vs hospital positive. There seems to be an equivalent number of events up to around 140 days where there begin to be marginally more in the hospital negative group. Positive cases in the community also seem to have a greater volume of events compared to community negative. Beyond 3 weeks both community groups appear to have a similar frequency of events shown by a similar gradient.

**Figure 8:**
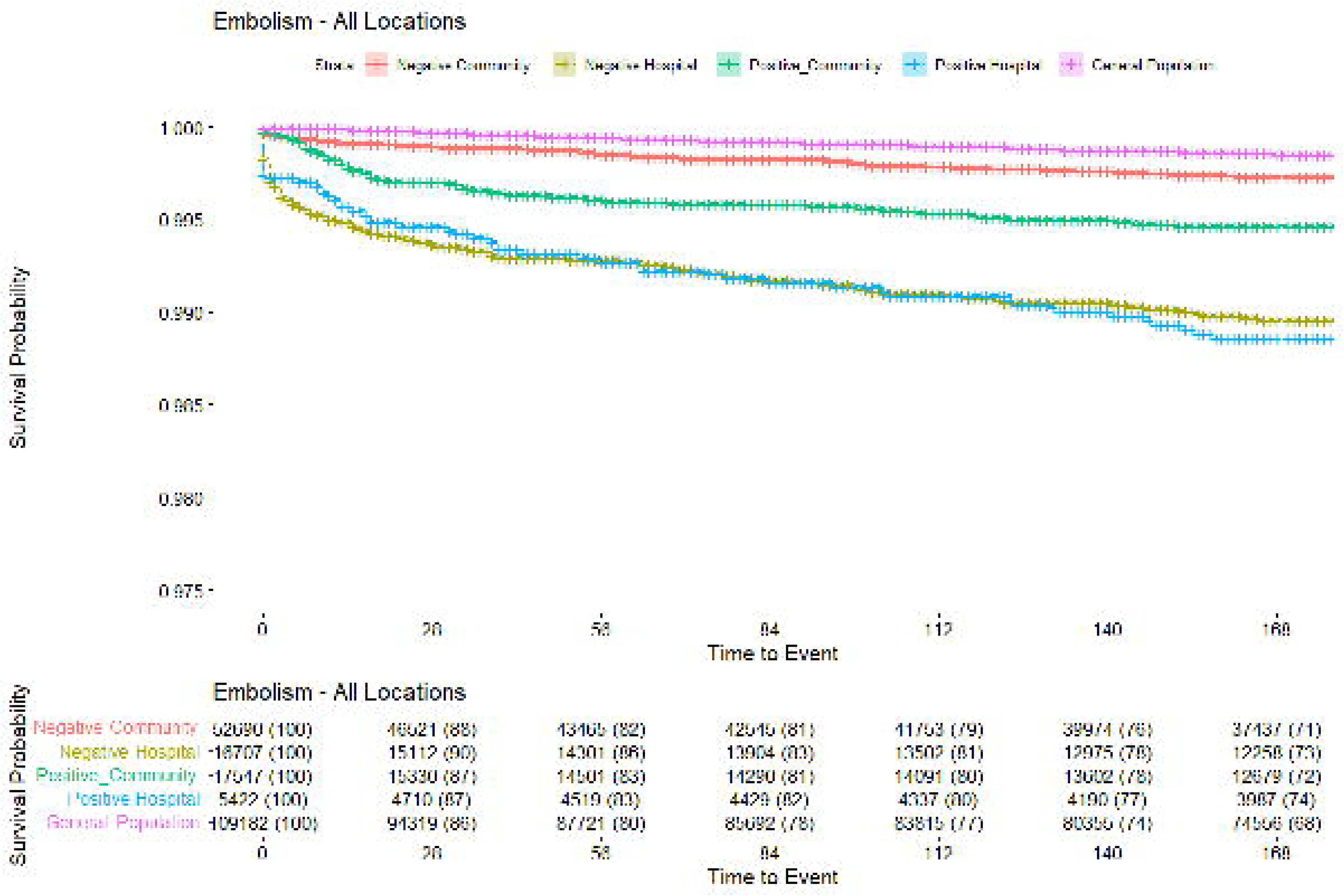
Time to First Embolism Code by COVID-19 diagnosis.

### Life Table Analysis

Table 2 shows individuals who tested positive in the community have an increased risk or increased trend of a first embolism and first fatigue code occurring through almost the entire follow up period. Community positive individuals are only less at risk of an embolism code 21-24 weeks following a test.

## Discussion

### Principle findings

This study examines the health care use 1-4 and 5-24 weeks following COVID-19 using propensity matched control methods to emulate a trial and suggest causal inference. It compares individuals who test positive for SARS-CoV-2 with controls who are propensity matched to account for deprivation, comorbidities, numbers in the households, number of previous SARS-CoV-2 tests (i.e., propensity to test positive), gender, age, and local authority area. These findings relate to testing prior to the identification of the Omicron variant and therefore includes all variants except Omicron. The cohorts were stratified by individuals testing in the community or hospital and their matches also needed to have been tested in the same stratification. Experiencing COVID-19, even if not accompanied by hospital admission, was associated with an increased risk of fatigue, post-viral illness, and a higher risk of embolism in the community cohort (e.g., code for Venous thromboembolism). The risk of death was greater for COVID positive individuals in the first four weeks, but no excess mortality risk was seen after that. Overall, positive individuals were less likely to receive codes for anxiety, depression, or self-harm. However, after 4 weeks positive individuals tested in hospital had an increased risk.

### Strengths and Limitations

This study has several strengths, it utilises an entire country’s (Wales) primary and secondary care data and so gives a national perspective of outcomes of COVID-19, making the work generalisable as it is a total population cohort. The use of propensity matching has the advantage of adjusting for many variables and so adjusting for propensity to contact COVID and account for a rich set of covariates which are associated with infection risk, so giving robust findings of association outcomes with surviving COVID. In addition, we were able to match and so control for differences between those tested in the community compared to when they attend a hospital. However, the matching did reduce the sample size of COVID patients from 249,390 to 98,600 which means a loss of 60% of COVID cases who did not have a match, this would result in a loss of precision for detecting rare events. Some limitations of this study include, the study only investigates the first occurrence and does not reflect total burden or duration of an existing problem for example, this study showed higher levels of fatigue in those with COVID, but it did not show how long this fatigue lasts for as the analysis gives a time to first mention of a fatigue diagnosis. This study examines engagement with healthcare and so can reflect use and burden to the health care system due to COVID specifically. However, it cannot capture the unmet need of people who have morbidity associated with COVID but do not seek healthcare for their illness. In addition, this study could not identify diagnosis which may not have a clear coding that can be captured using the GP or hospital system, such as memory loss, or brain fog which have been found to be associated with COVID (16).

### Comparison with other studies

The finding that those who survive COVID show higher rates of cardiovascular disease is in agreement with other published findings, such as findings that several cardiovascular disorders are higher in veterans data in the US (17) and higher rates of venous thromboembolism (6) using CPRD data in the UK. However, the finding that there is no overall increase in diagnosis of mental health problems was at odds with literature from the US veterans study(16) and a study using the US TriNetX (3) dataset which both find higher rates of psychiatric morbidity and mental health diagnosis after COVID (18). The difference in findings may be due to differences in the variables used to the propensity scores to match with test negative patients, or real differences in risk of mental health conditions associated with the health care system (US compared to UK) and with population included e.g. US veterans cohort vs Wales population cohort.

### Implications and future research

The absolute numbers of contacting their health care professional with long term effects of COVID are low and there was no increased need for sick notes compared to a matched comparison group after 4 weeks. Therefore, the findings are reassuring that post-COVID adverse consequences do occur but the overall number of people seeking health care for this are low. It must be noted though that some adverse events such as embolism are serious and so clinicians should be aware of higher rates for a prolonged period in those who have had COVID. In addition, more work is needed to examine the burden to patients who are not seeking health care.

### Conclusions

This used a national cohort of people with COVID-19, showed risk of fatigue, post-viral syndrome and embolism is higher compared to a propensity matched comparison group and this was for those hospitalised and non-hospitalised and lasted for at least 24 weeks. Absolute numbers of those attending health care settings for complications of COVID are low (e.g. 1% of those surviving COVID) but some of the burden may be undiagnosed due to sufferers not presenting to a health care setting.

## Data Availability

All data produced in the present study are available upon reasonable request to the authors

## Acknowledgements

This study is part of the National Centre for Population Health and Wellbeing, which is funded by Health Care Research Wales. This study makes use of anonymised data held in the Secure Anonymised Information Linkage (SAIL) Databank (9,19,20). We would like to acknowledge all the data providers who make anonymised data available for research.

This work was supported by Health Data Research UK, which receives its funding from HDR UK Ltd (HDR-9006) funded by the UK Medical Research Council, Engineering and Physical Sciences Research Council, Economic and Social Research Council, Department of Health and Social Care (England), Chief Scientist Office of the Scottish Government Health and Social Care Directorates, Health and Social Care Research and Development Division (Welsh Government), Public Health Agency (Northern Ireland), British Heart Foundation (BHF) and the Wellcome Trust.

The responsibility for the interpretation of the information supplied is the authors’ alone.

## Appendix

**Appendix A:** Individual SARS-CoV-2 testing sites included under each testing location.

|  | ***Test Location*** |  |
| --- | --- | --- |
| **Test Site** | **Community** | **Hospital** |
|  | Care Home | AE |
|  | Community Unit | Hospital |
|  | Covid Unit | ICU |
|  | CTU | Outpatient |
|  | GP | Hospice |
|  | HC Staff |  |
|  | Police |  |
|  | Prison |  |
|  | Self-Home Test |  |

**Appendix B:** Location origins of the codes used to define the outcomes in the study. Additional notes also provided.

| Outcome | Codes |  | Notes |
| --- | --- | --- | --- |
|  | Primary Care | Secondary Care |  |
| Diabetes | ✓ | ✗ | Only Primary Care |
| Embolism | ✓ | ✓ |  |
| Fatigue | ✓ | ✗ | Only Primary Care |
| Influenza (Only) | ✗ | ✓ | Only Secondary Care |
| Mental and Behavioural Disorders | ✓ | ✓ | Anxiety, Depression, OCD, Schizophrenia, ADHD, Bipolar, Autism, Self-Harm |
| Anxiety, Depression and Self Harm Only | ✓ | ✓ |  |
| Post Viral Syndrome | ✓ | ✗ | Only Primary Care |
| Respiratory (All) | ✓ | ✓ | General Respiratory, Mastoid, Upper and Lower Respiratory Infection, Asthma |
| Respiratory (Influenza Removed) | ✓ | ✓ |  |
| Sick Notes | ✓ | ✗ | Only Primary Care |

**Appendix C:**
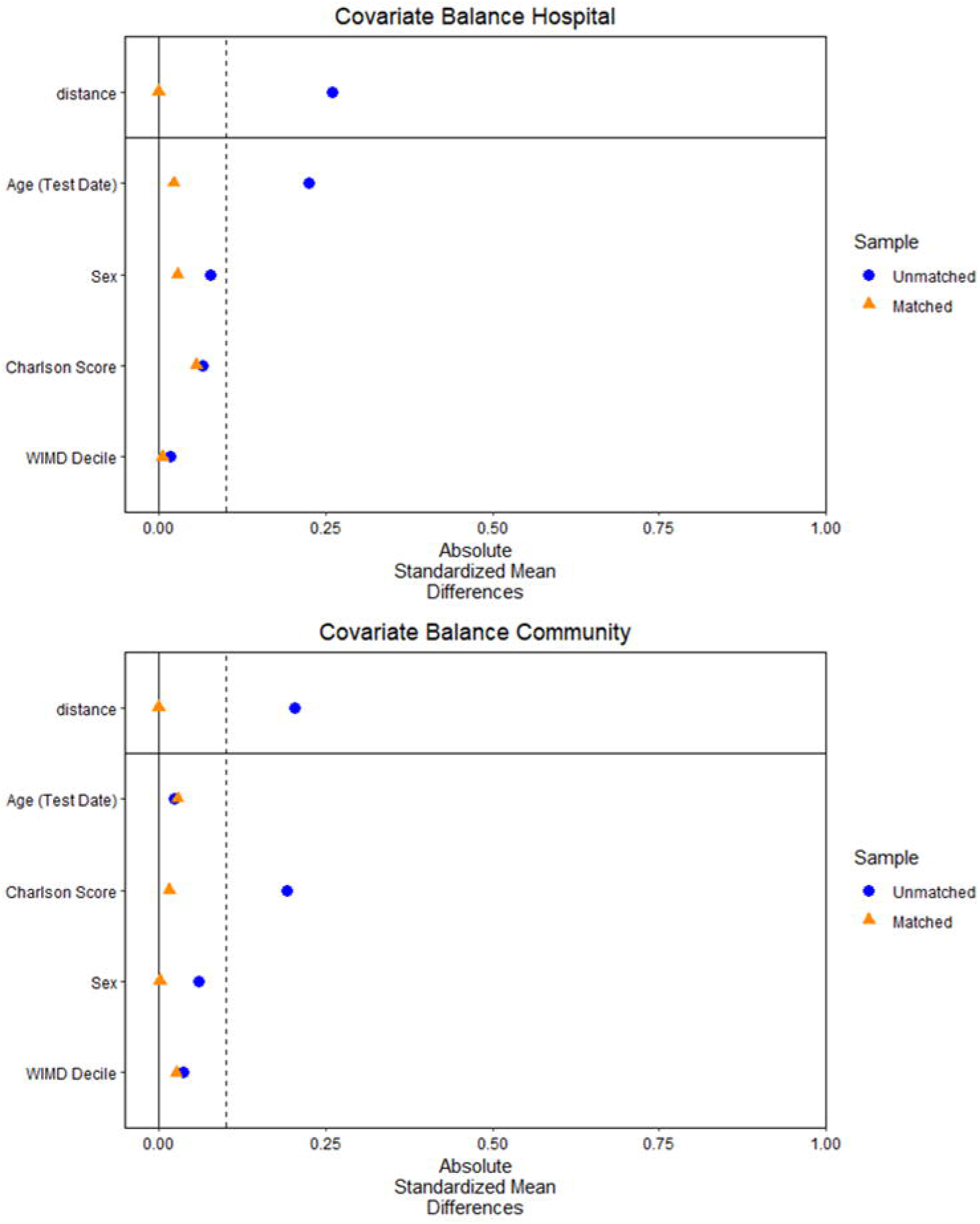
Shows ‘Love Plots’ for the main covariates before and after the propensity matching had taken place for community and hospital tested individuals.

**Appendix D:** Shows the raw values for figure 2 which show death, primary and secondary care codes per 100,000 individuals.

| Deaths/100,000 |  |  |  |  |  |  |
| --- | --- | --- | --- | --- | --- | --- |
| Time (Weeks) | 1-4 | 5-8 | 9-12 | 13-16 | 17-20 | 21-24 |
| Negative | 980.36 | 557.53 | 344.63 | 294.23 | 250.13 | 199.08 |
| Positive | 3019.77 | 552.76 | 303.06 | 180.42 | 156.34 | 106.11 |
| Untested | 128.23 | 140.55 | 127.75 | 107.39 | 91.89 | 77.29 |

**Appendix E:**
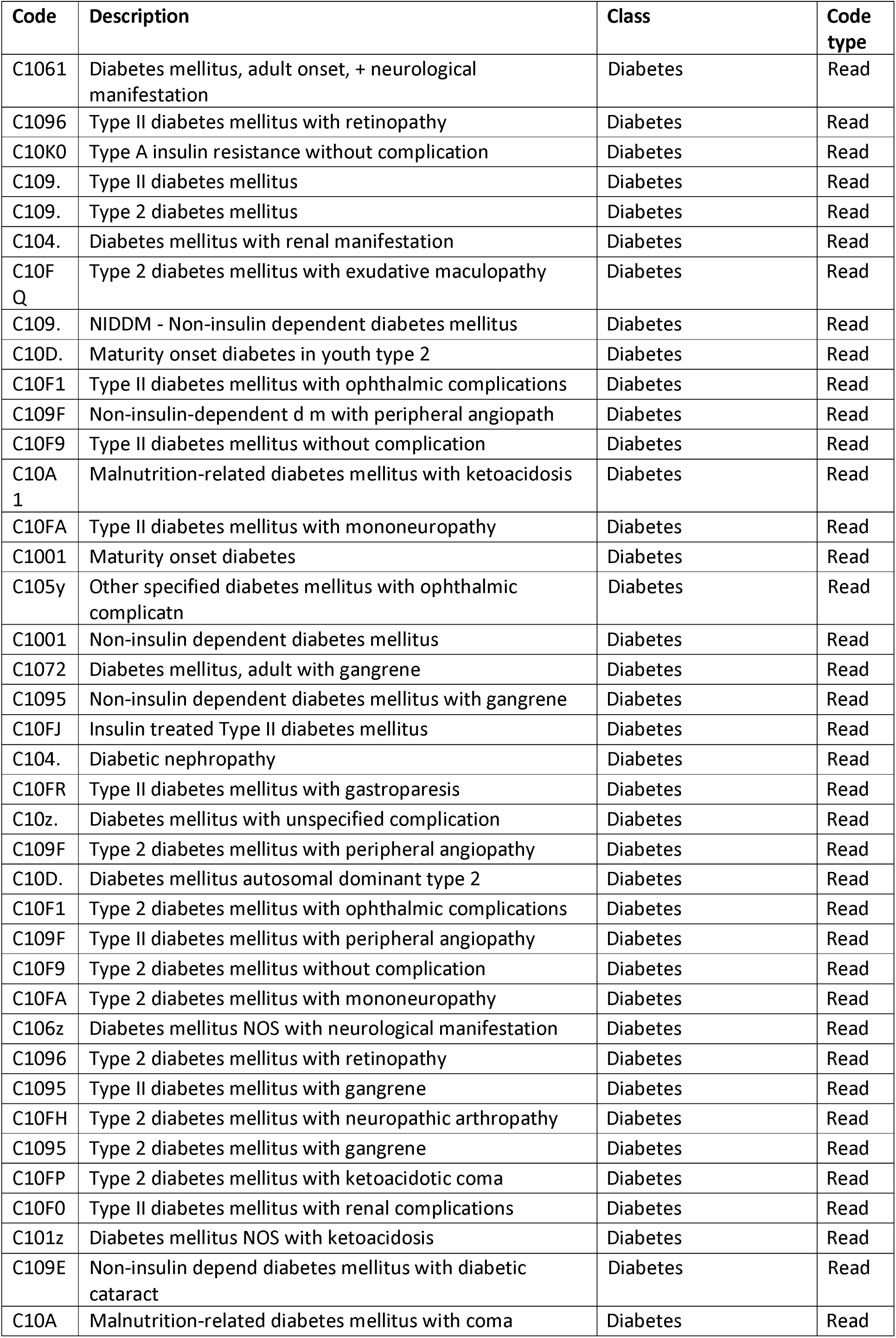
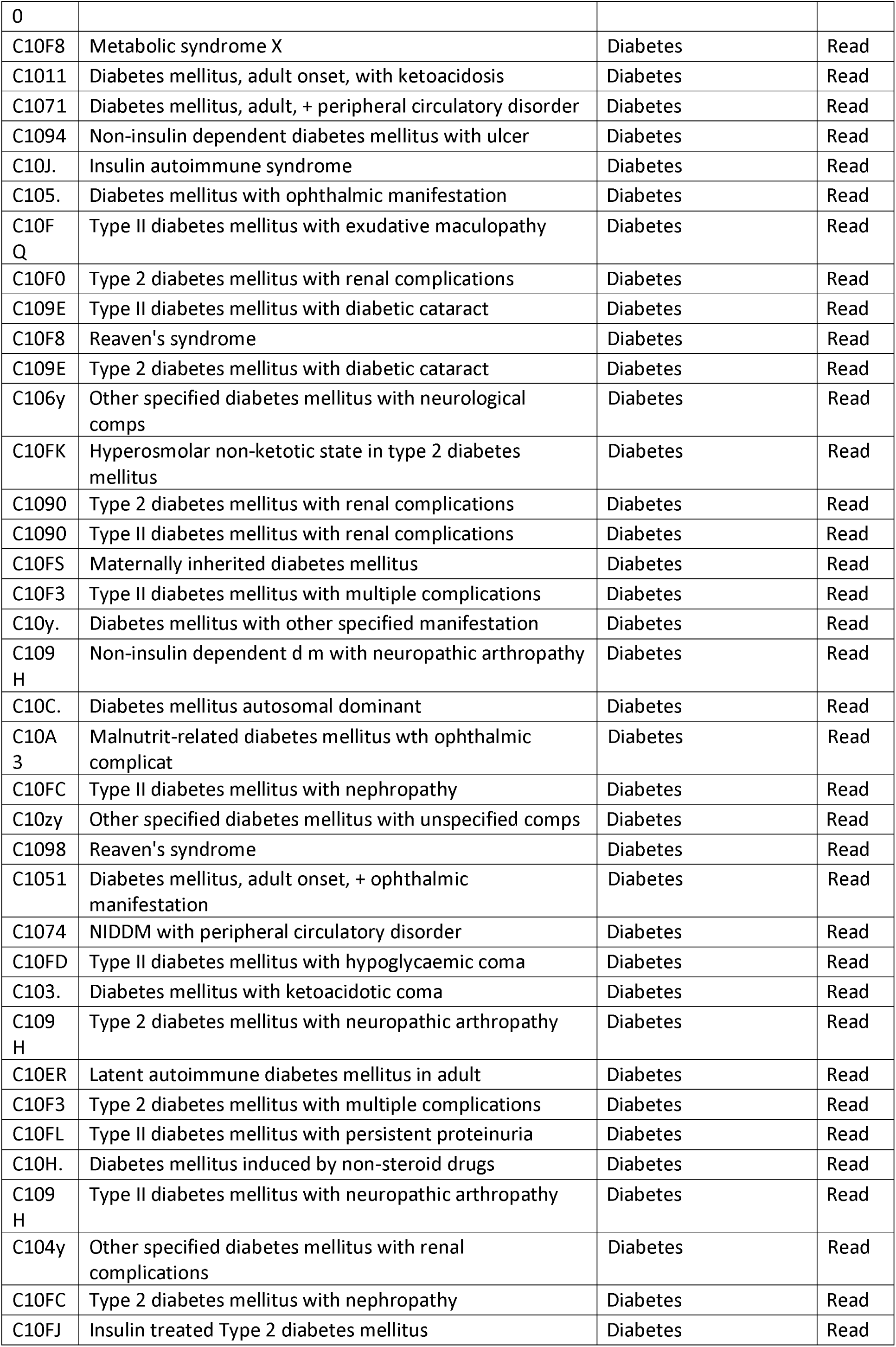
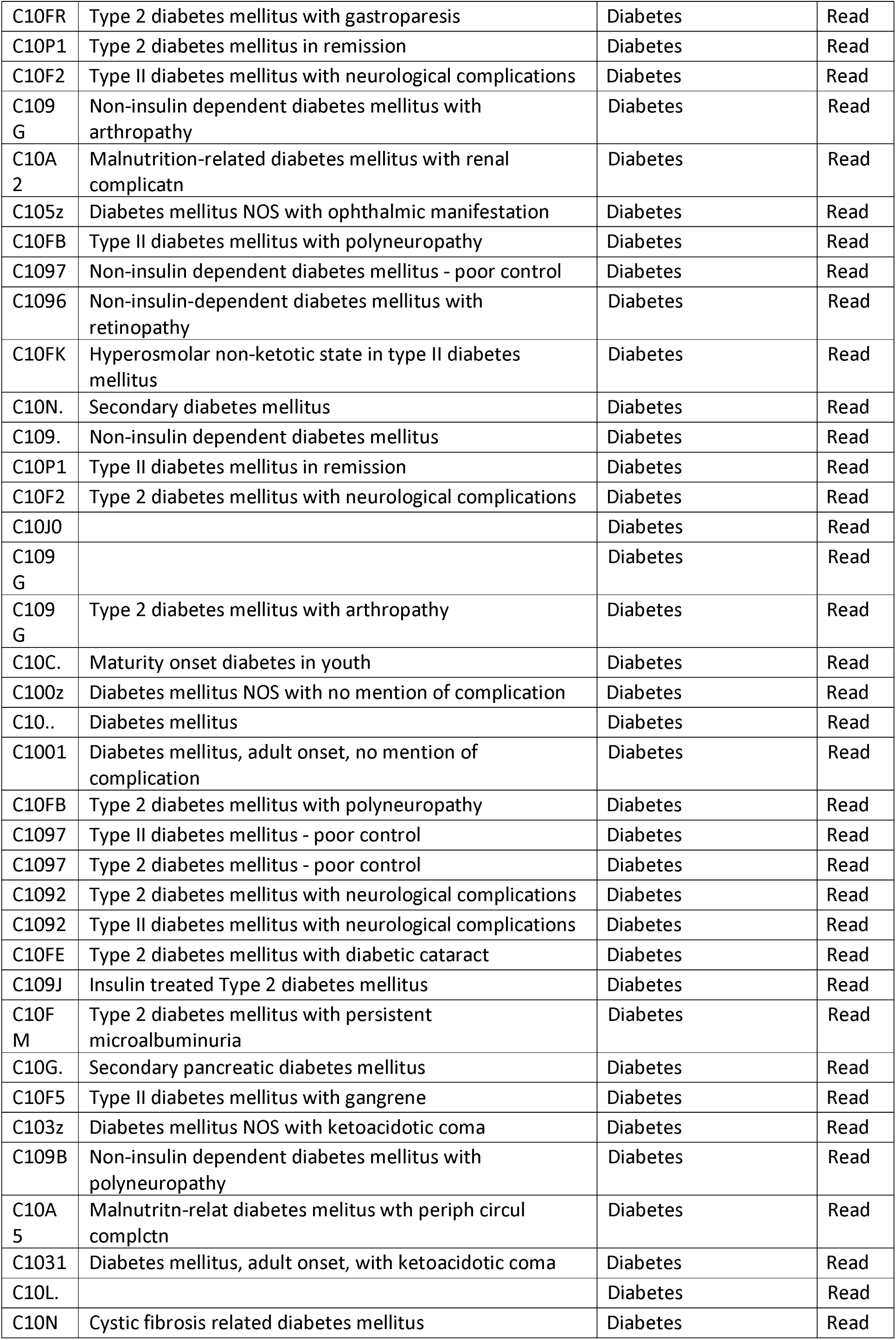
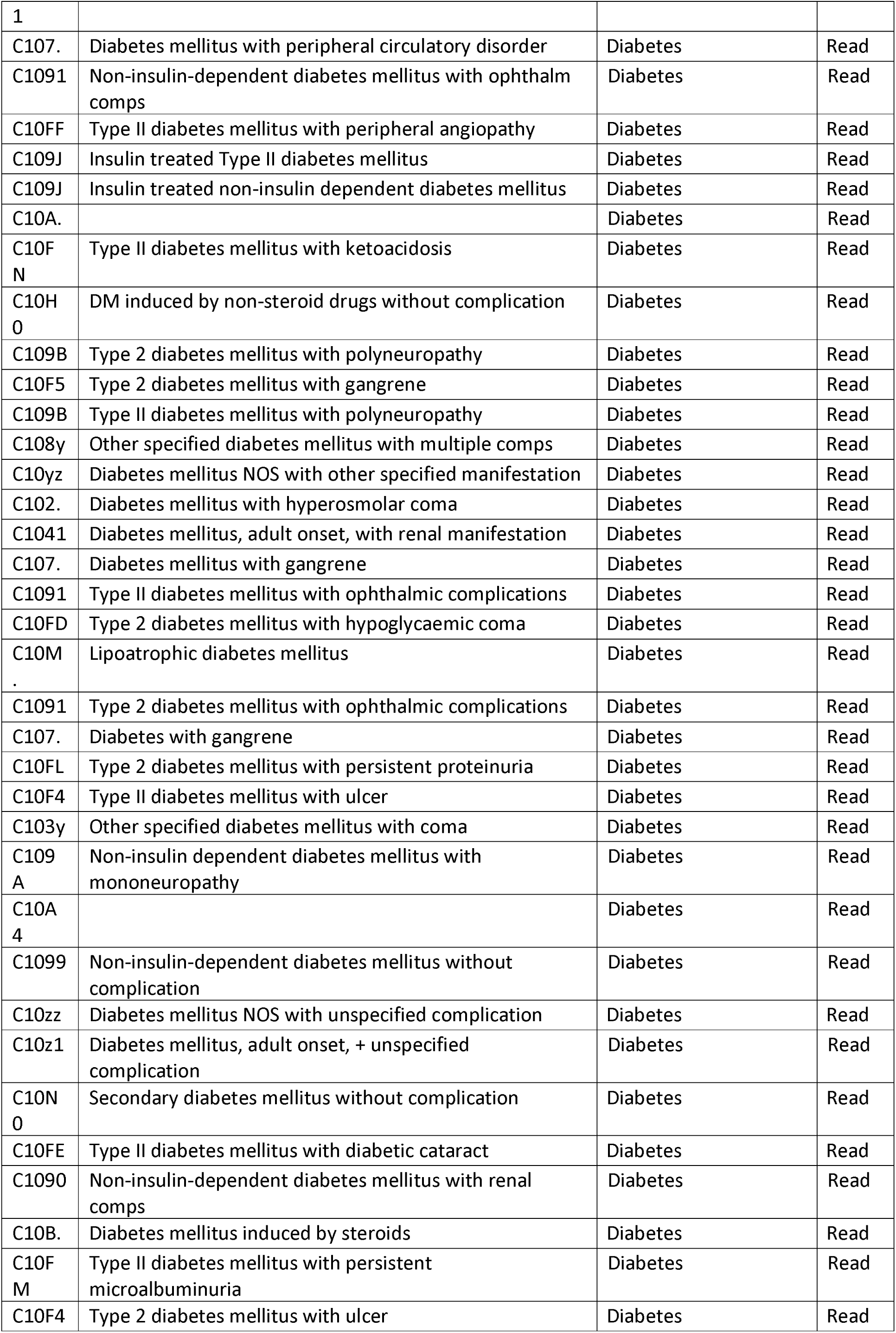
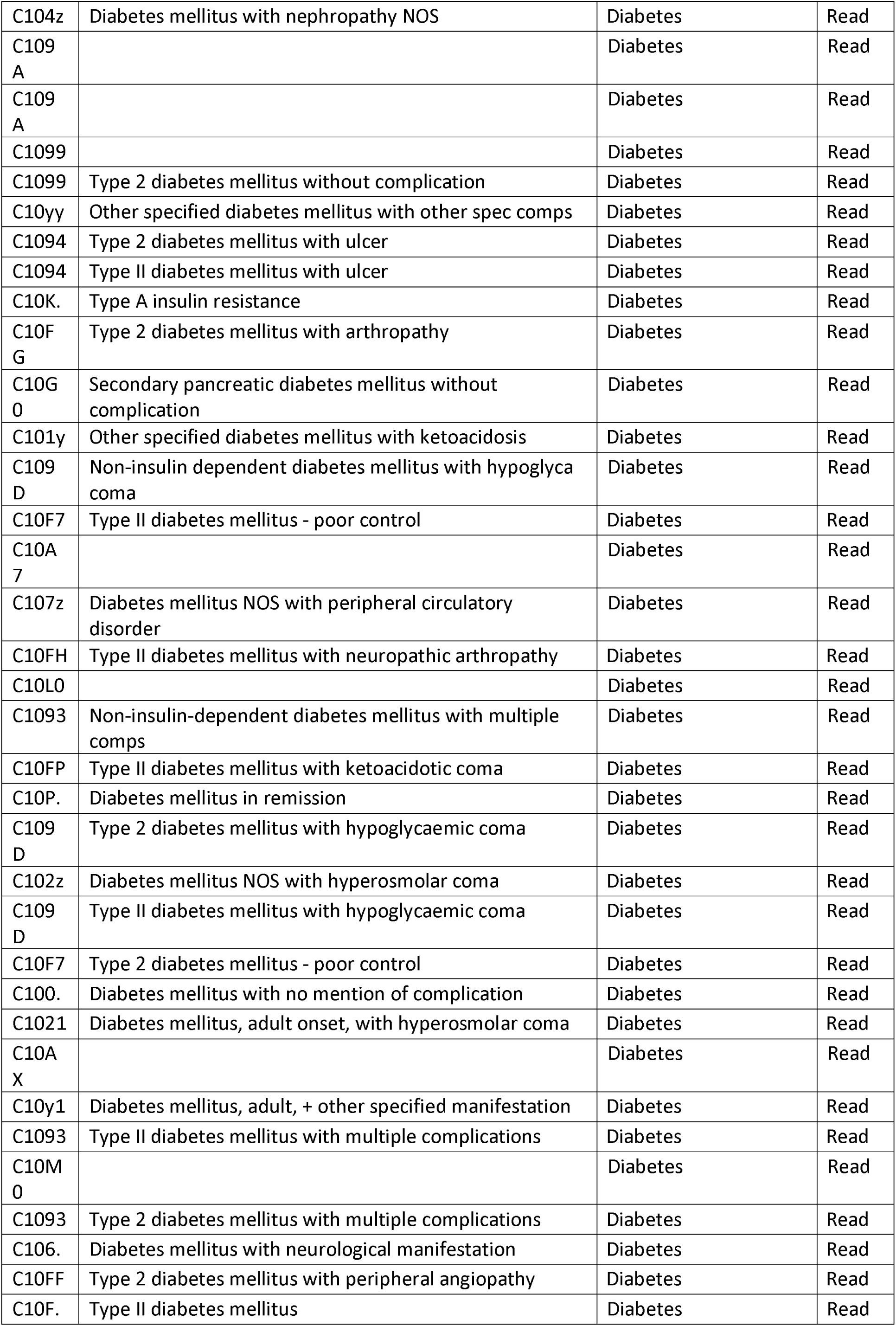
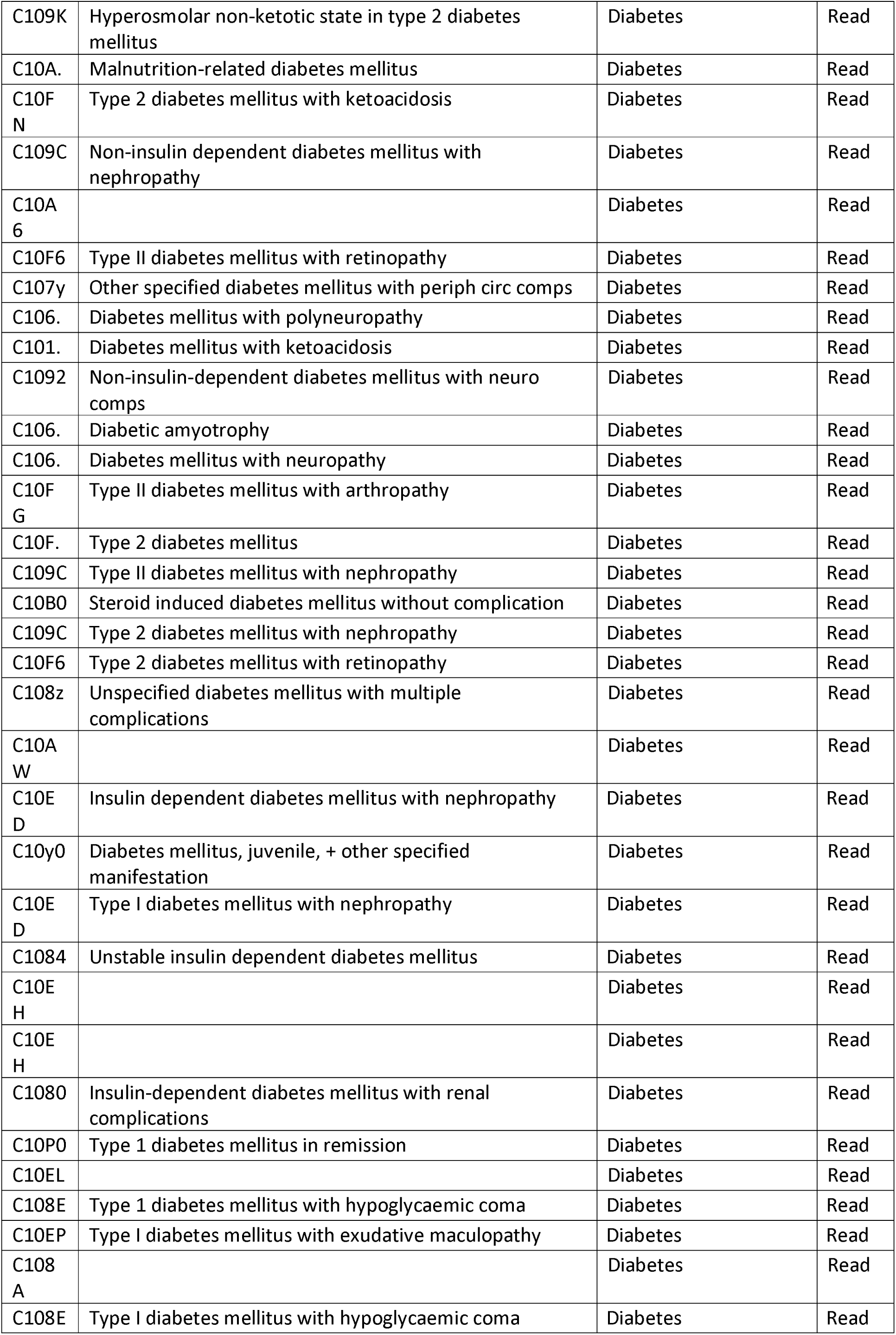
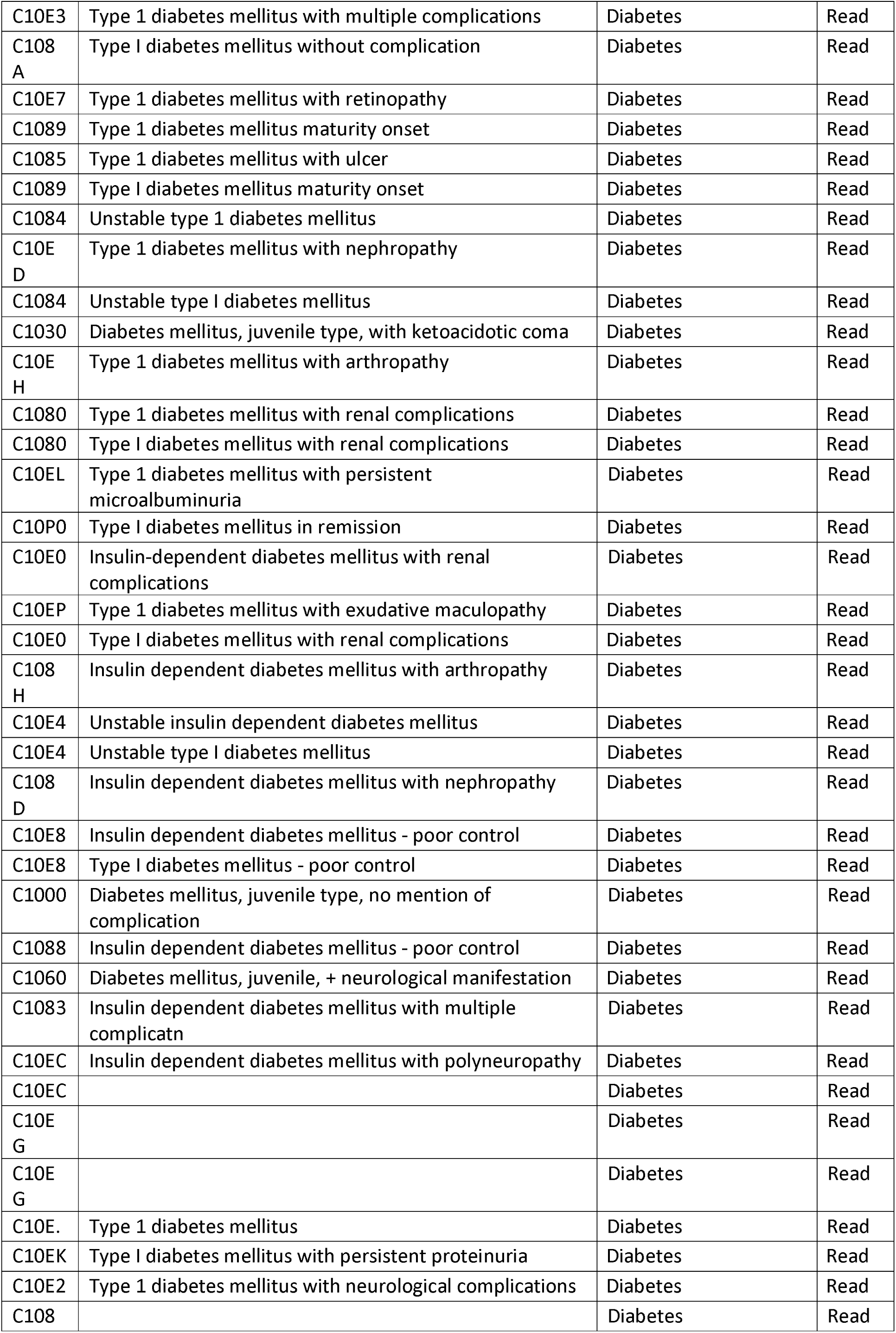
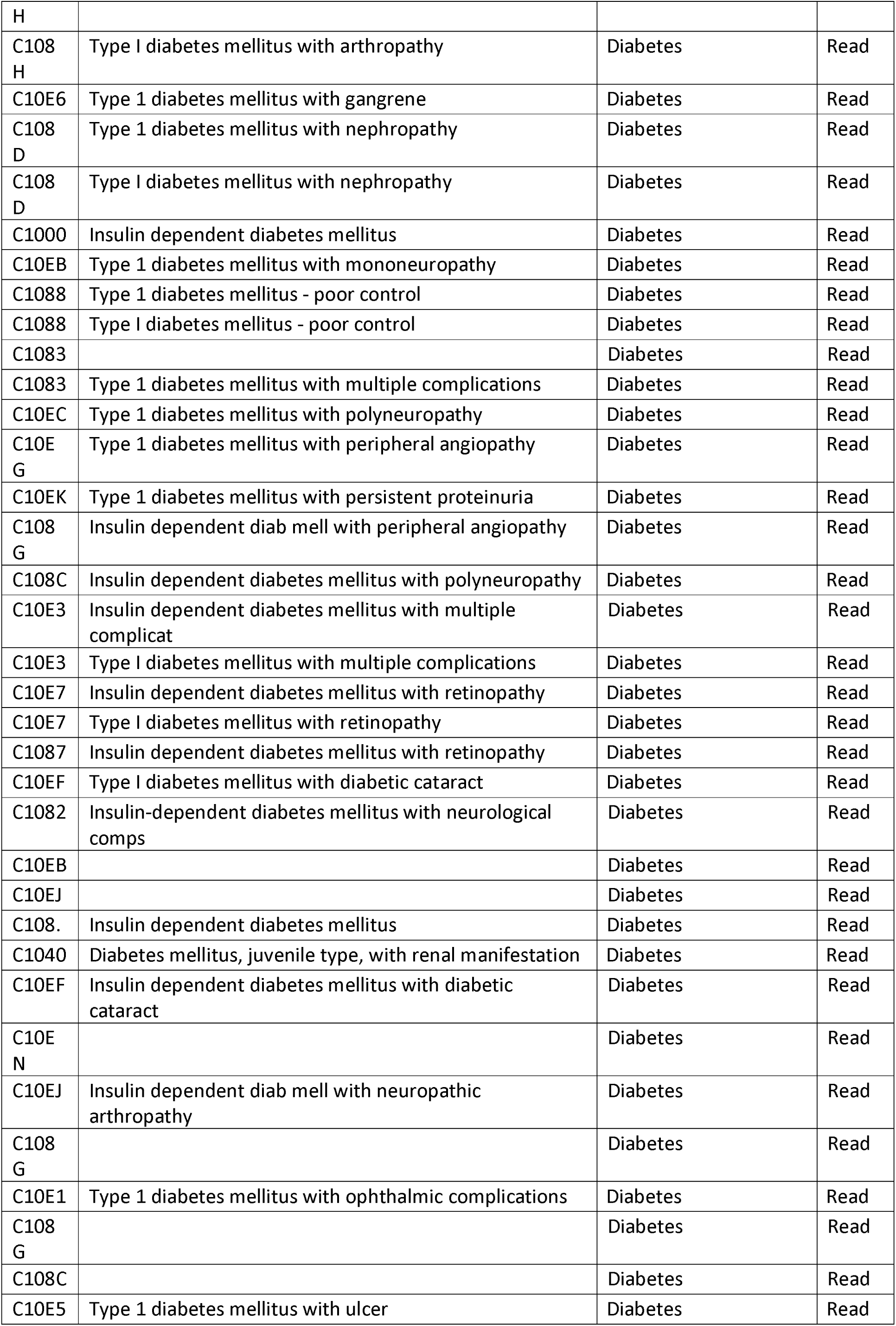
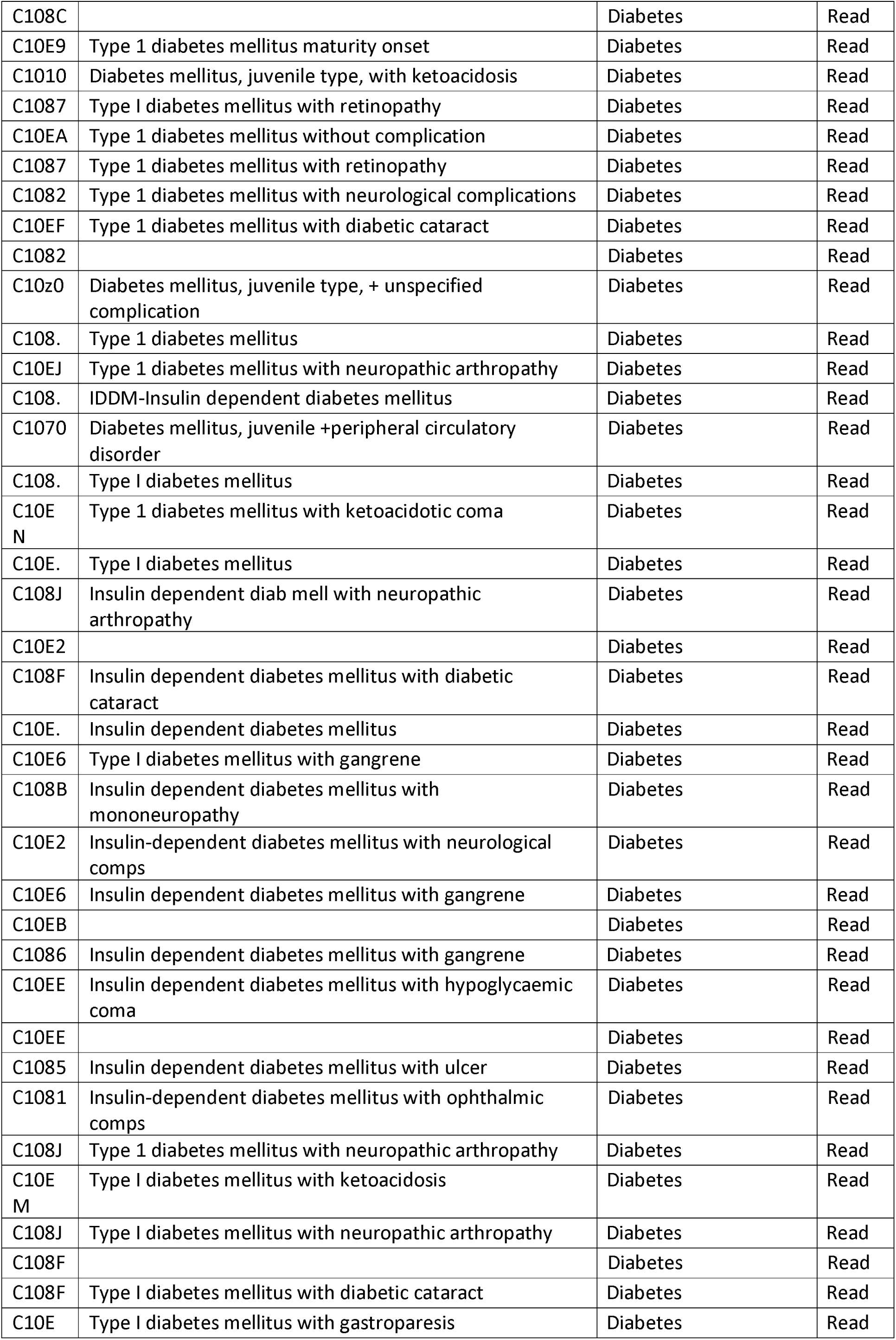
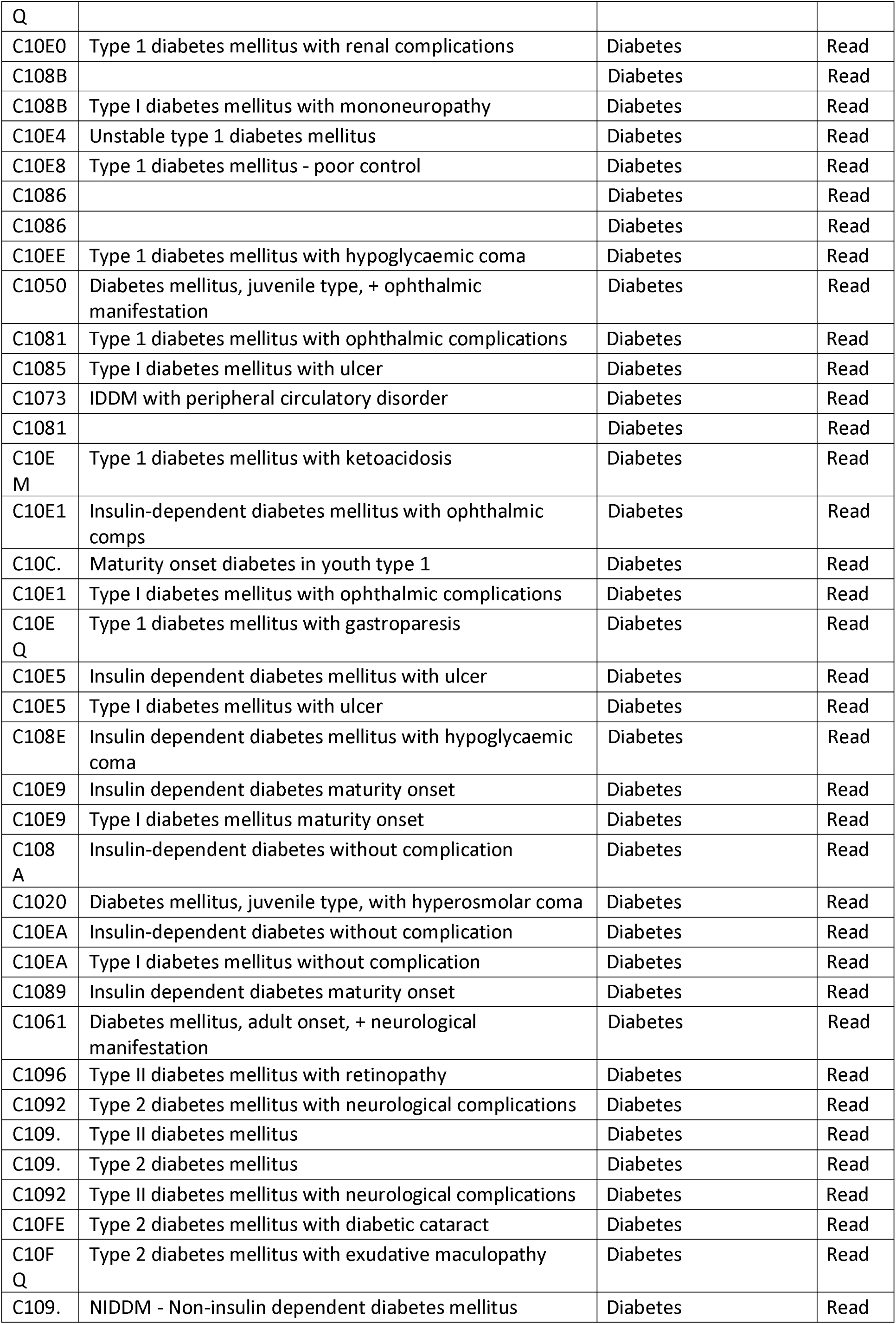
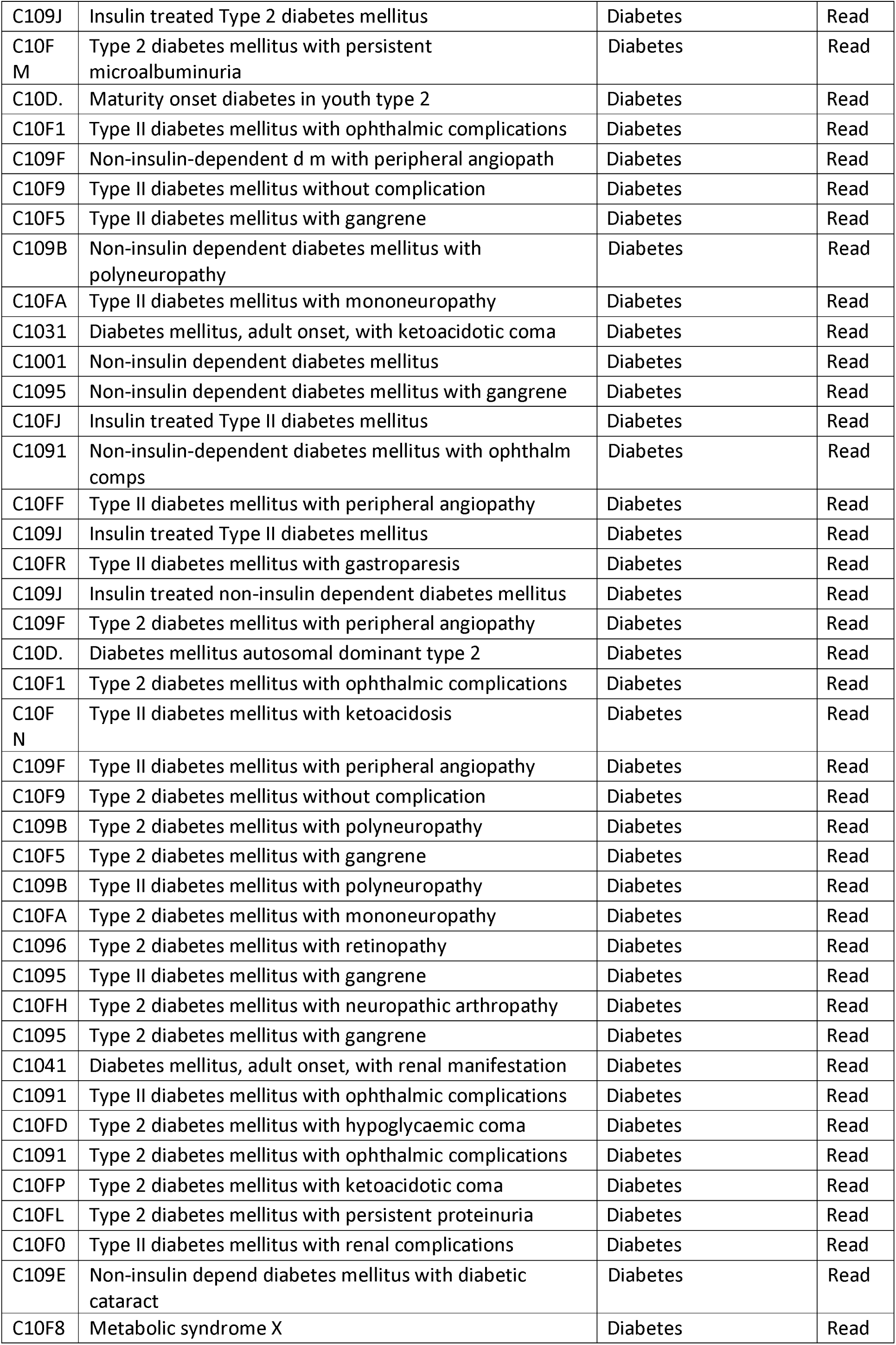
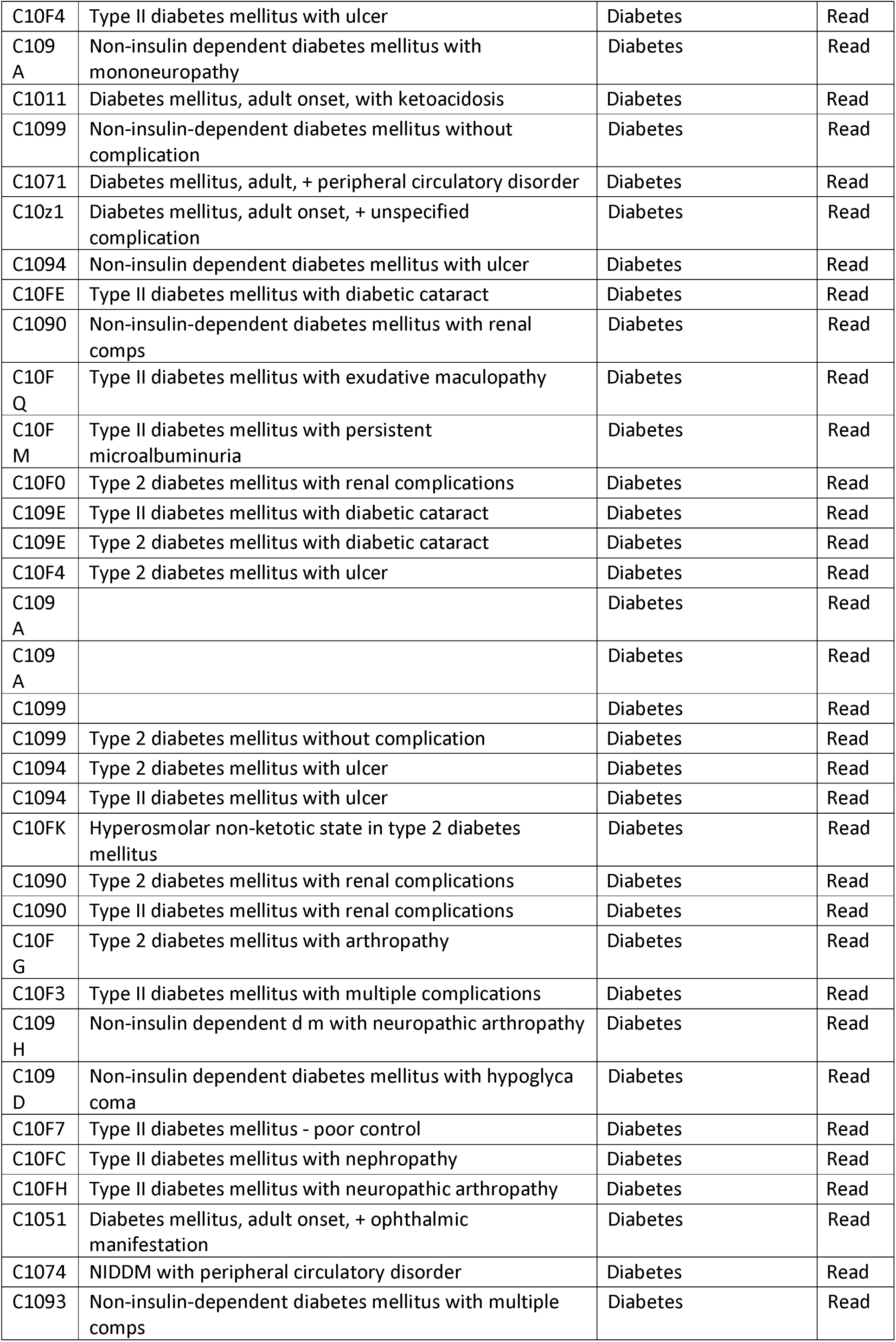
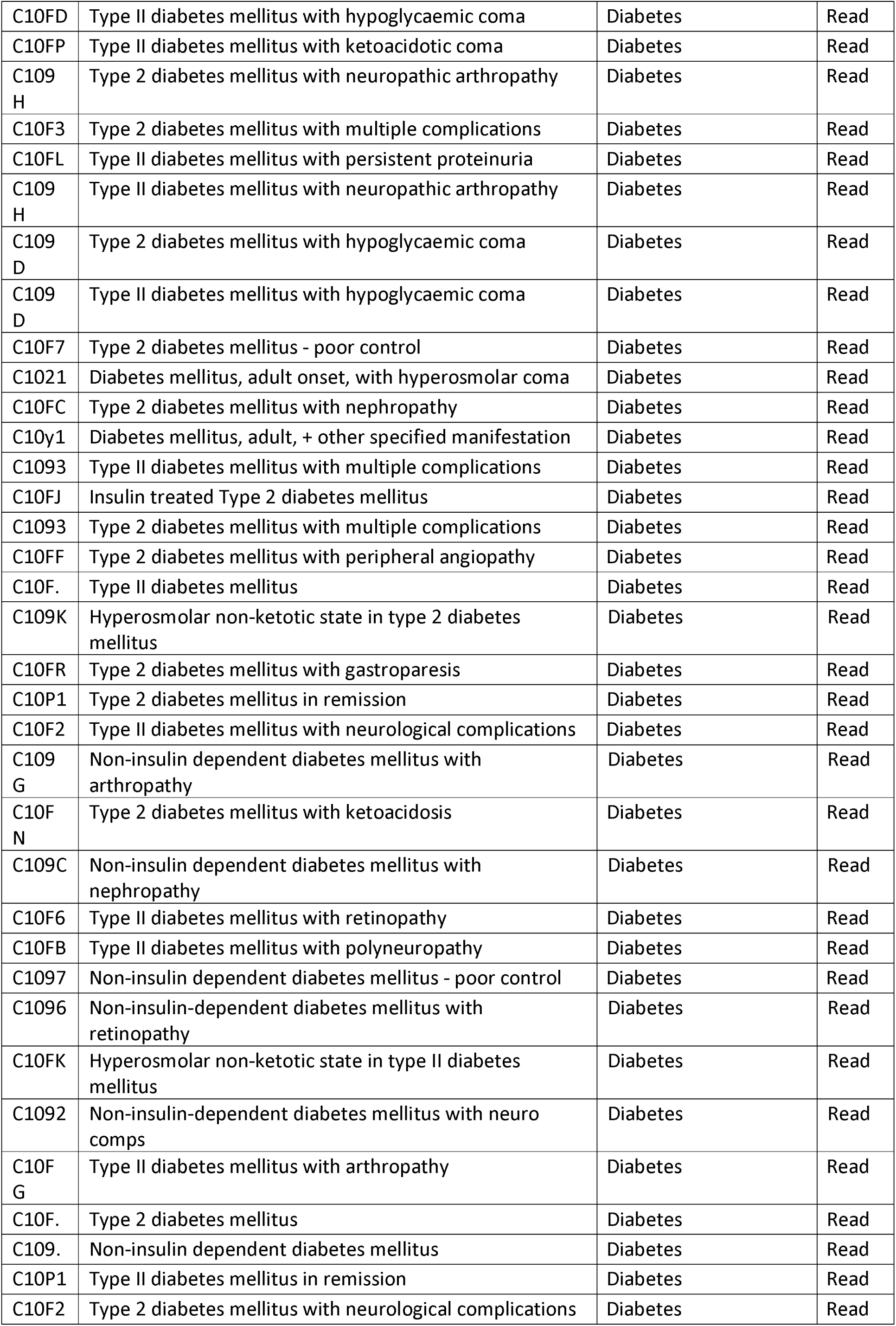
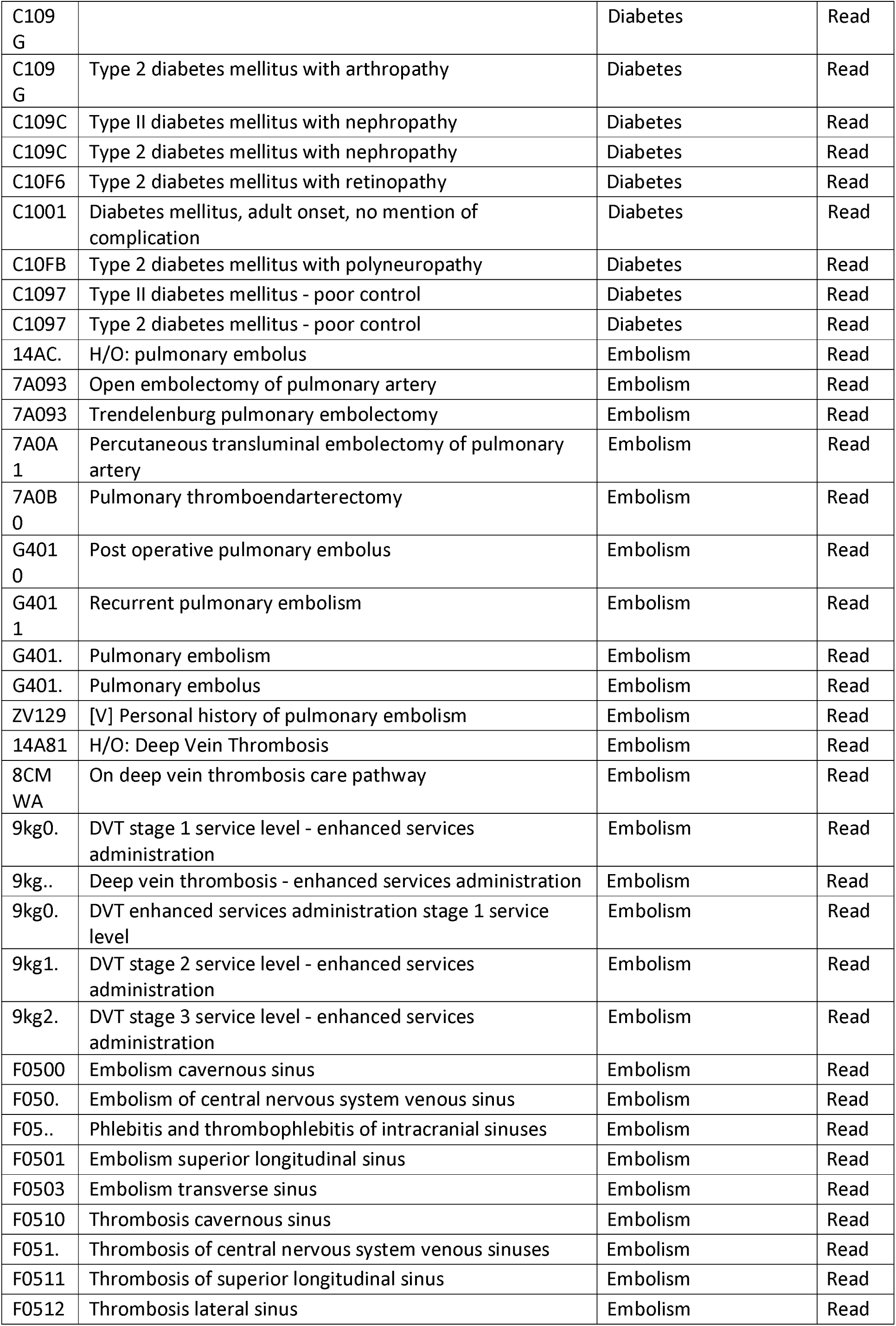
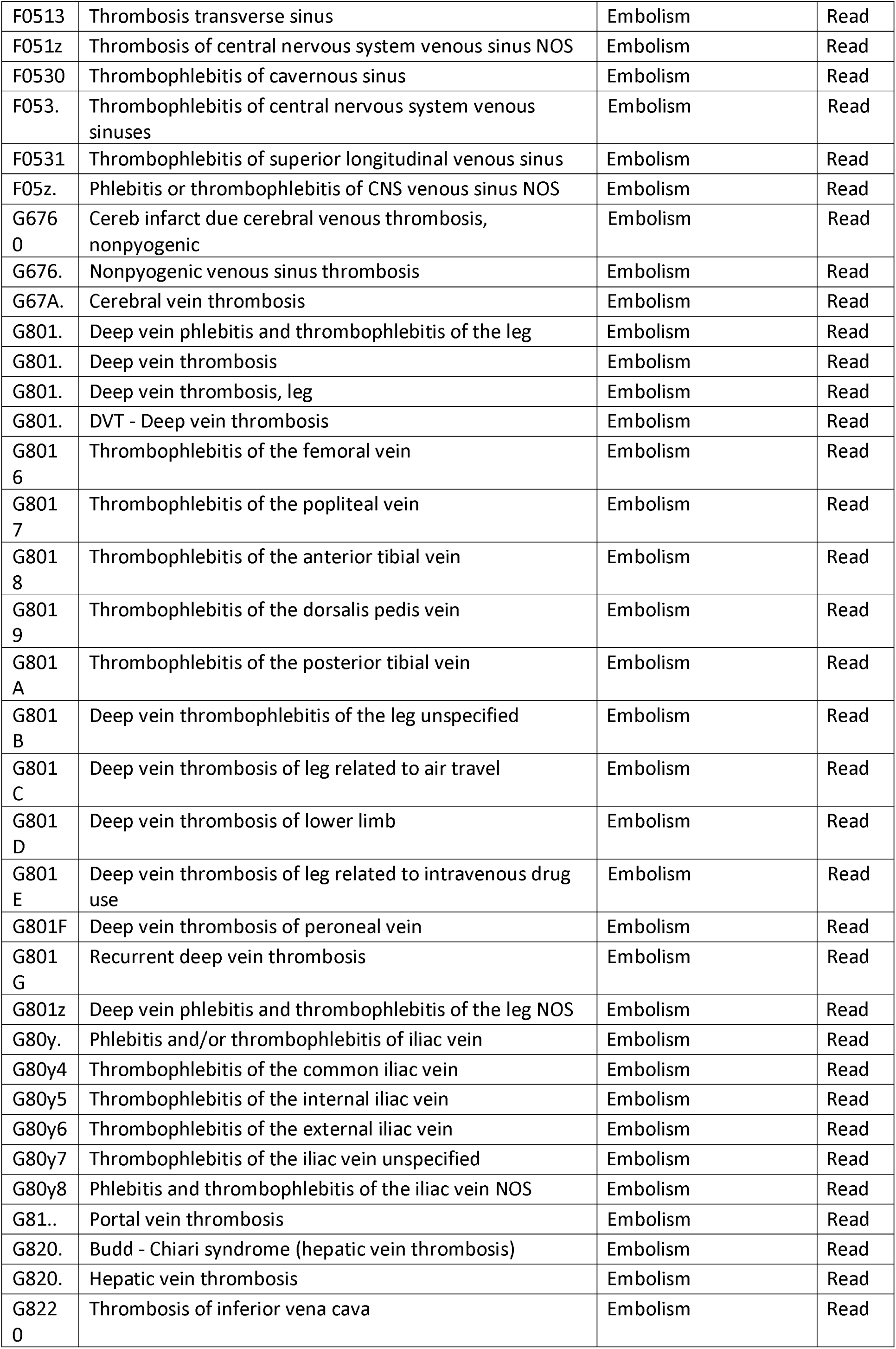
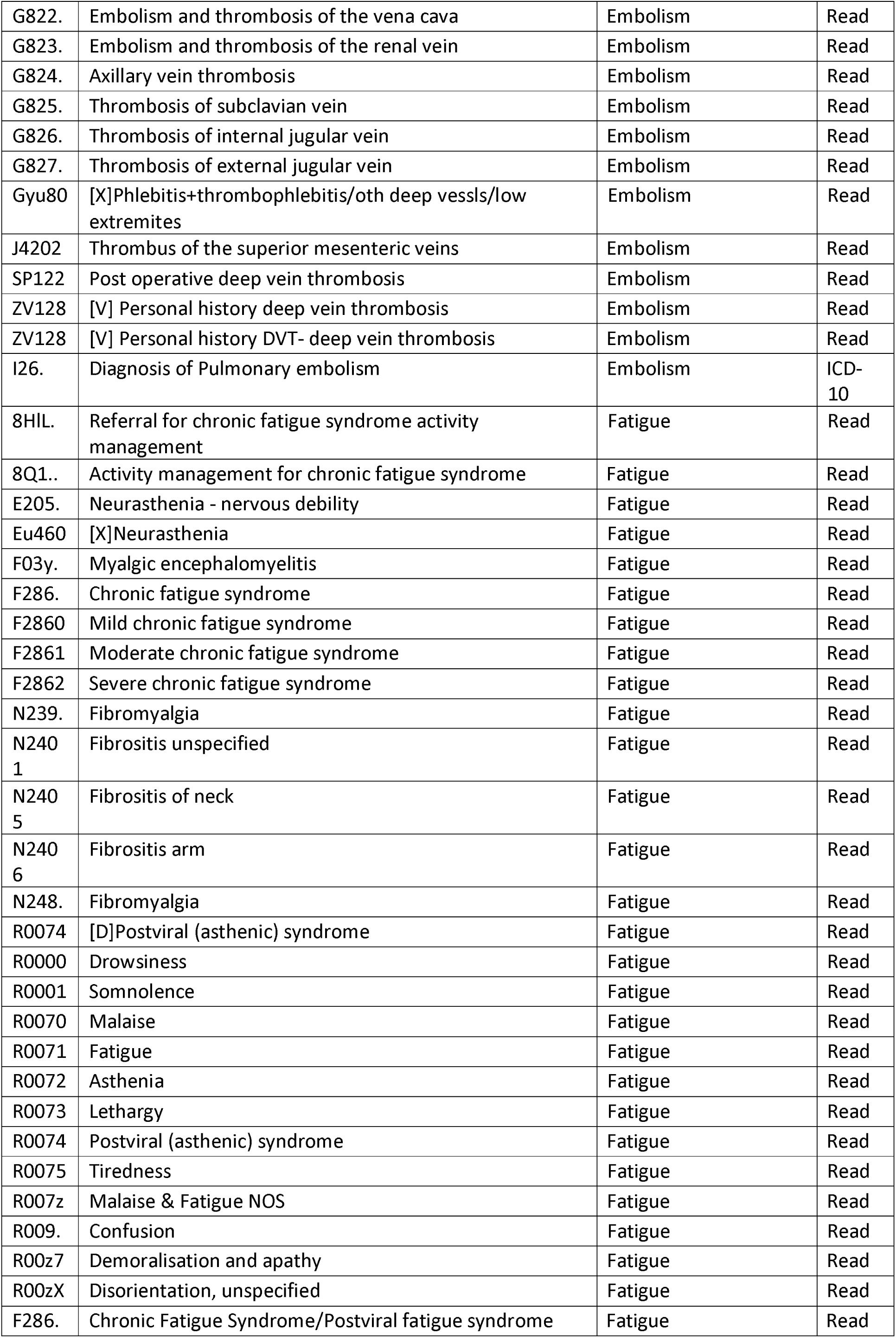
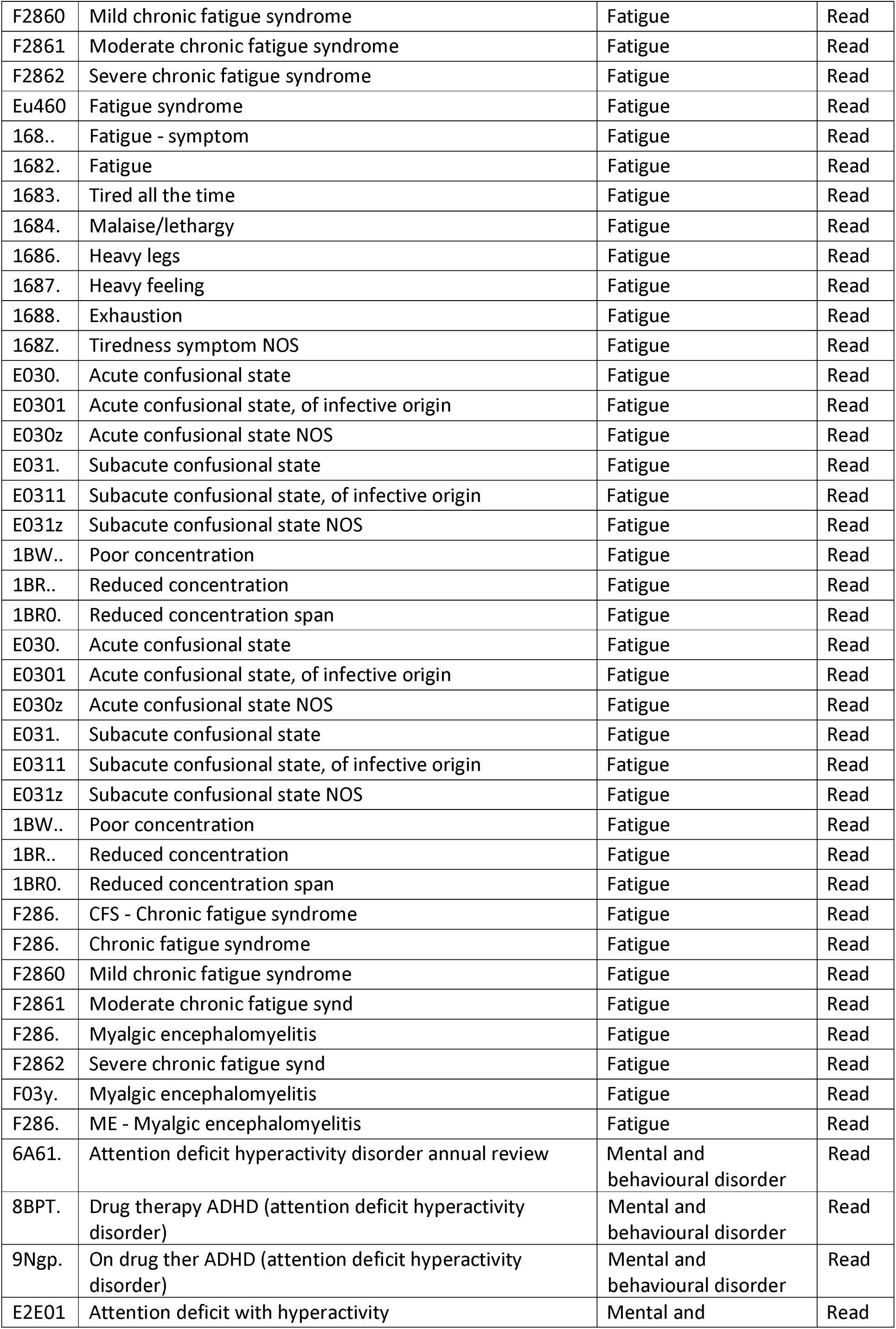
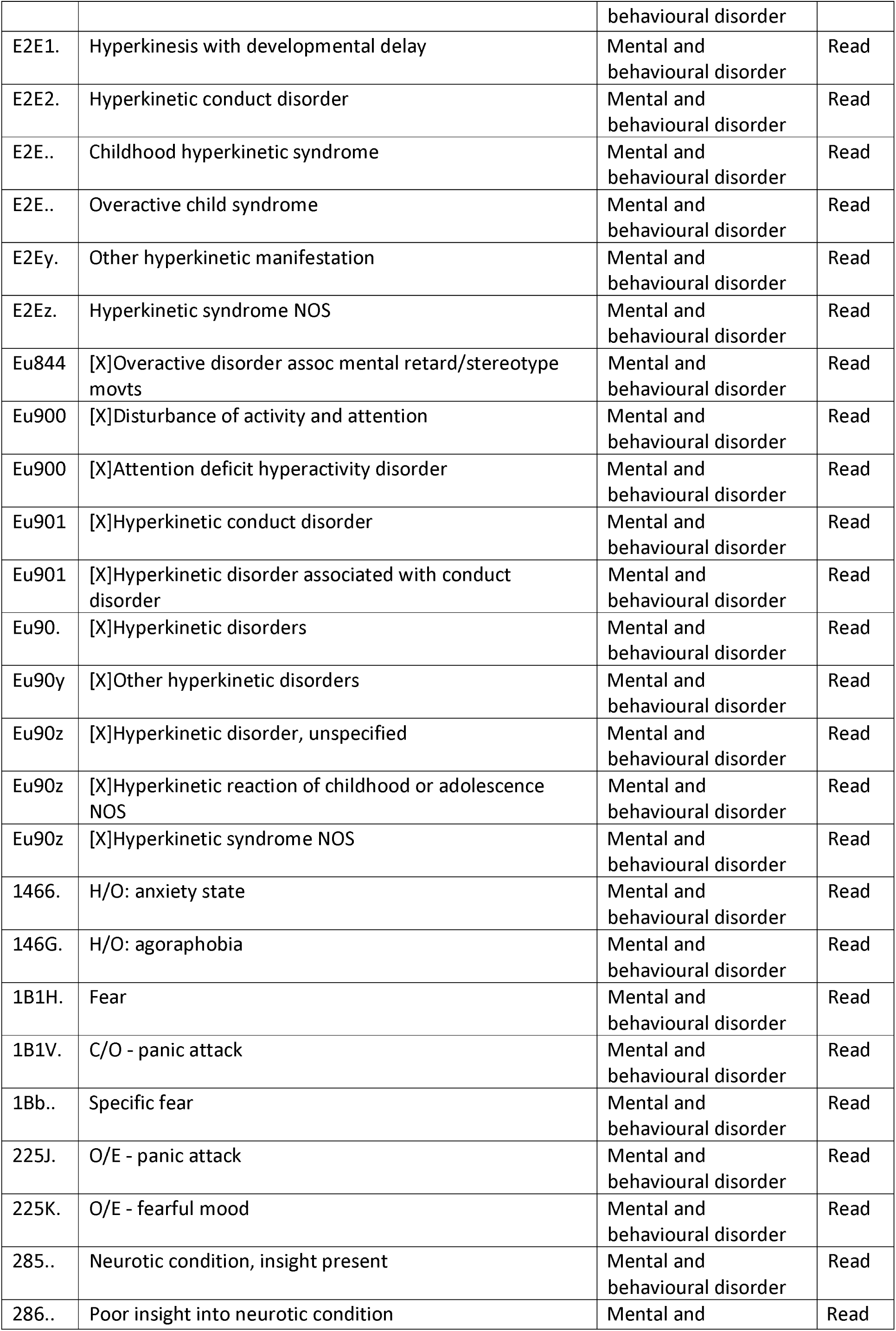
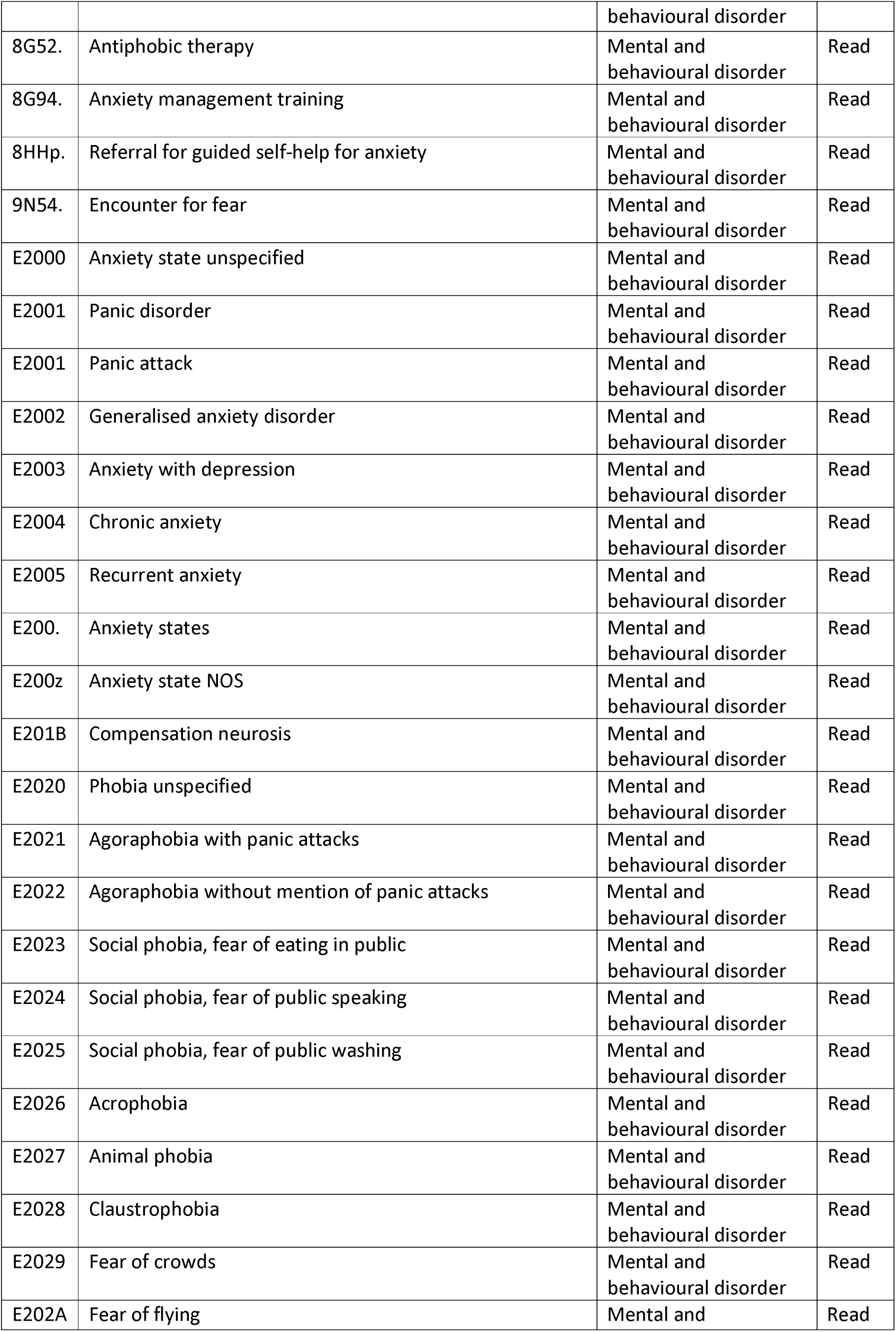
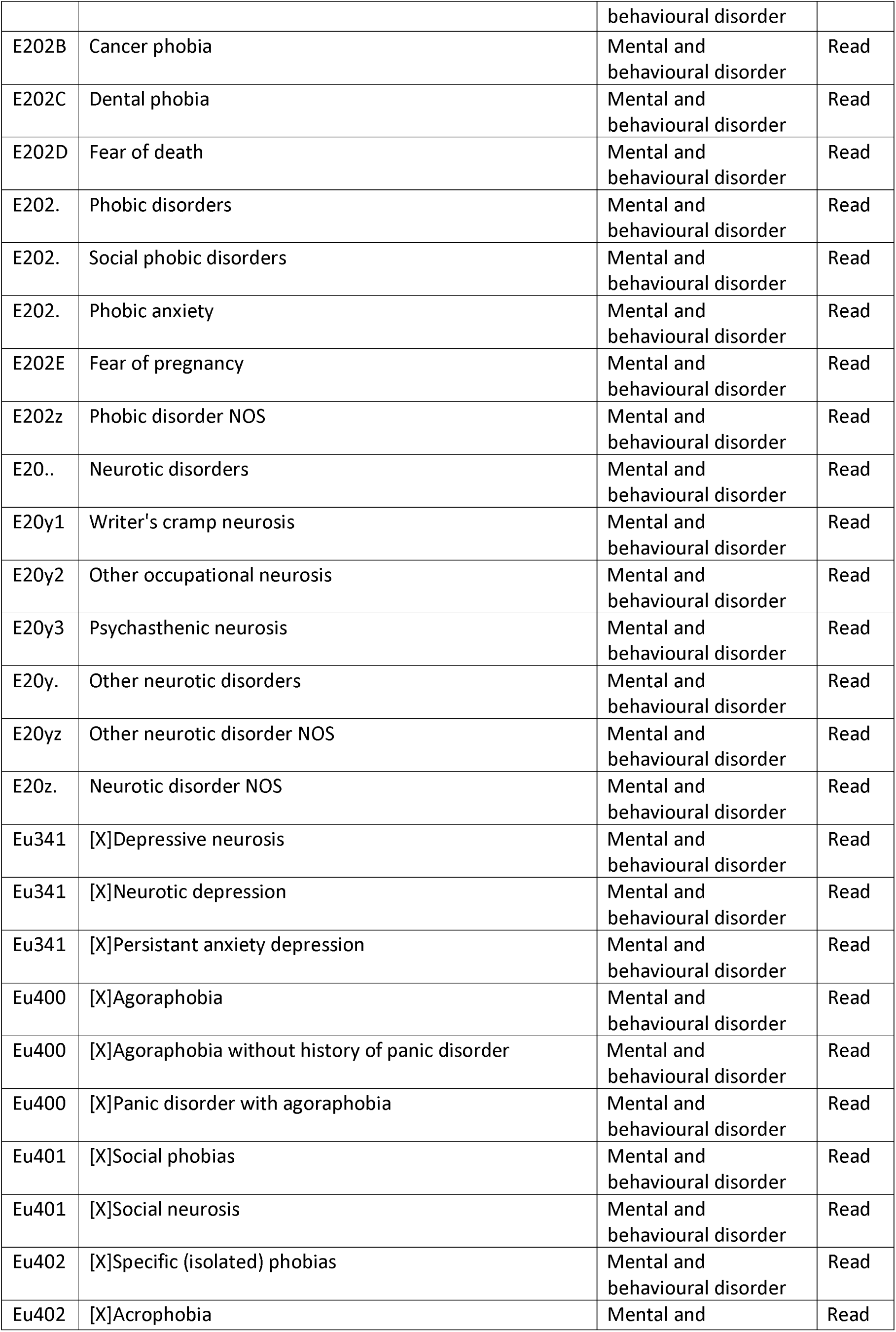
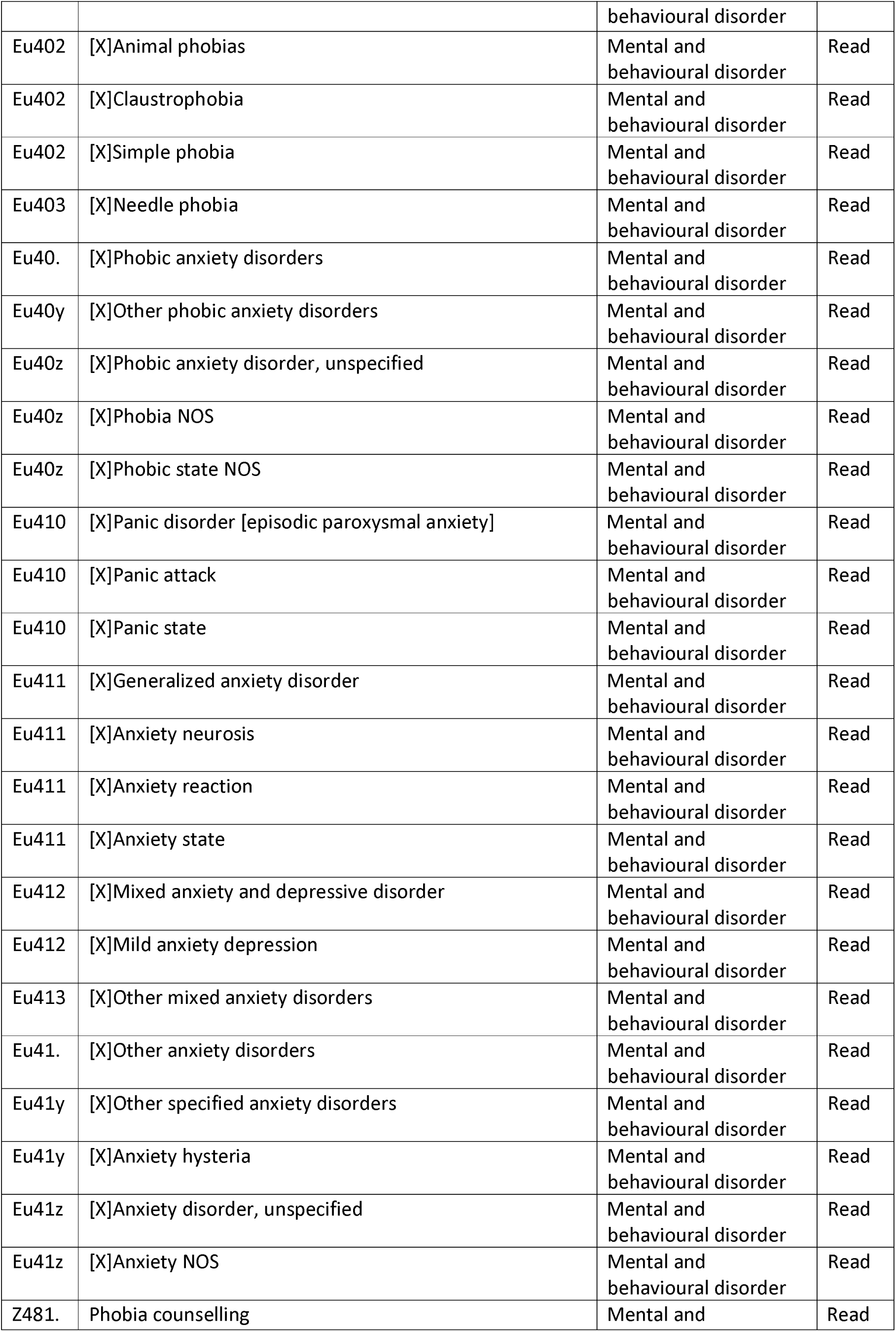
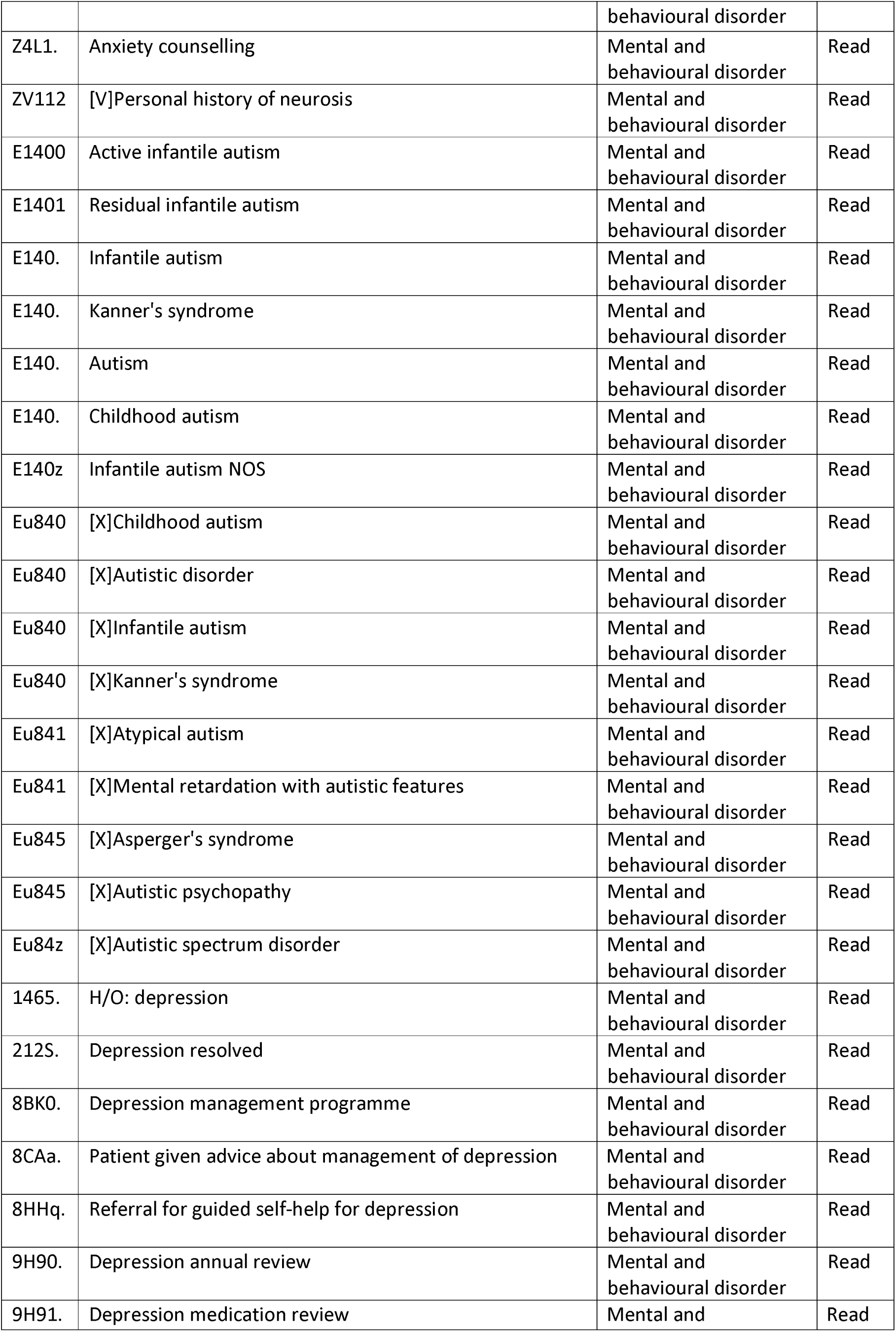
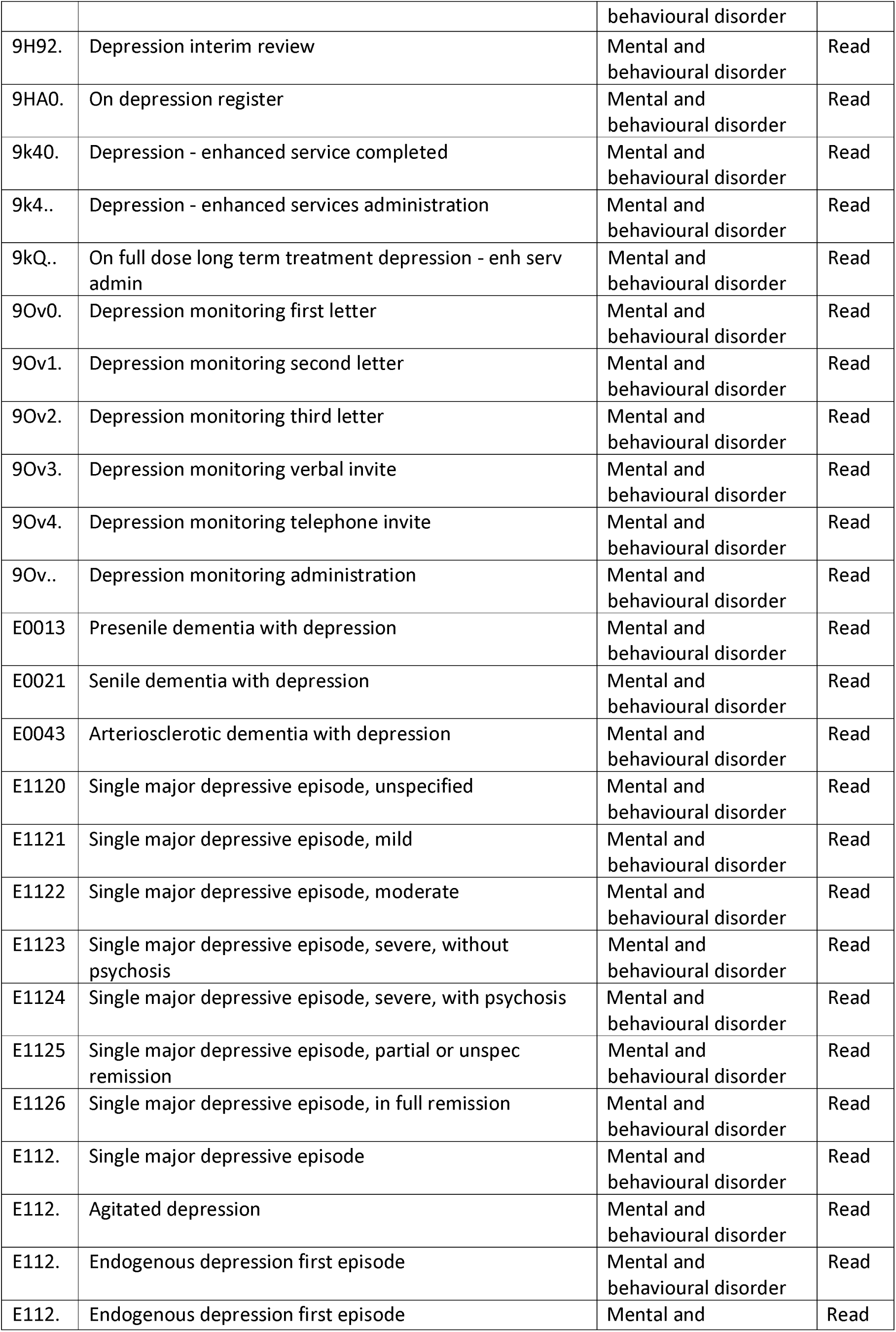
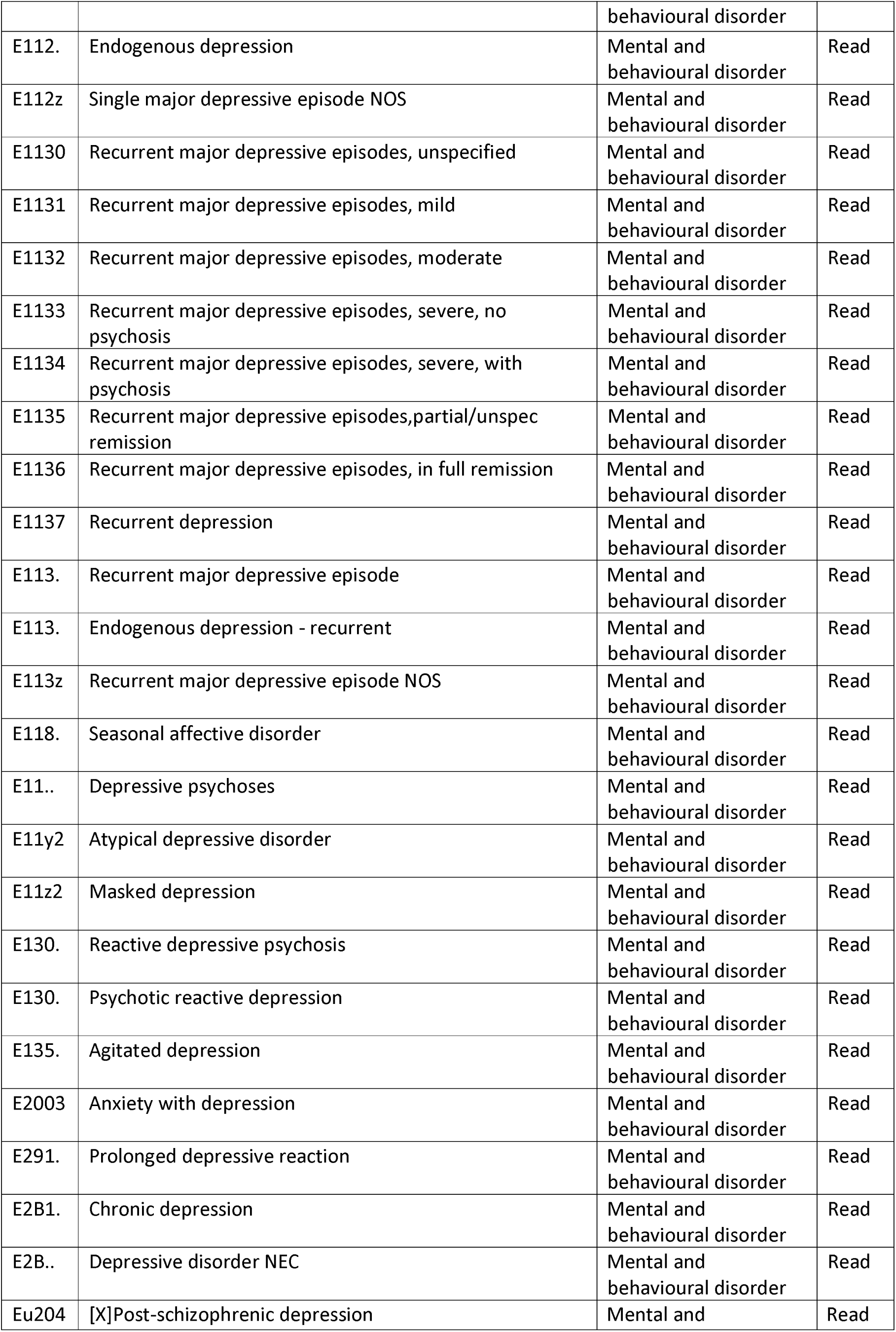
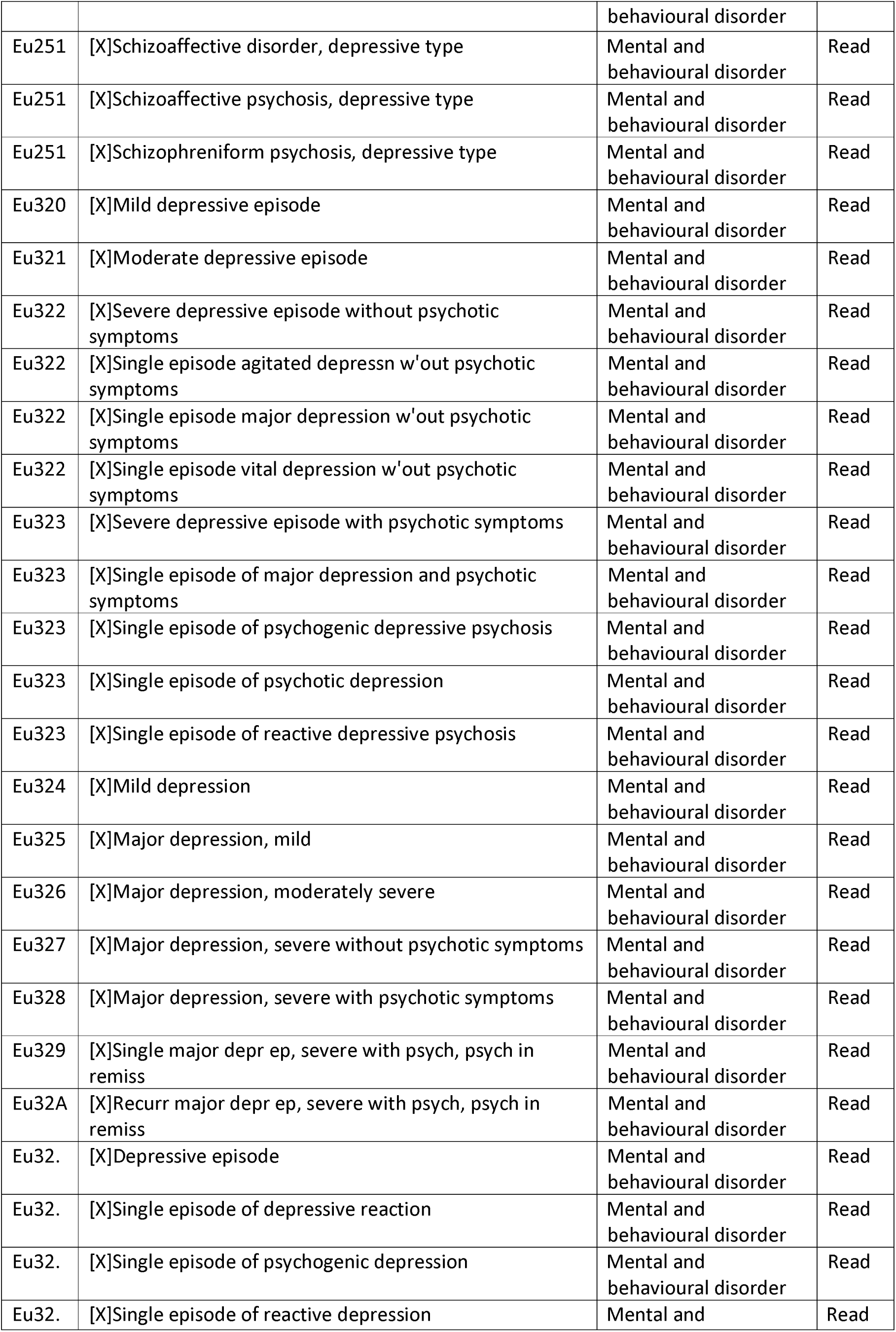
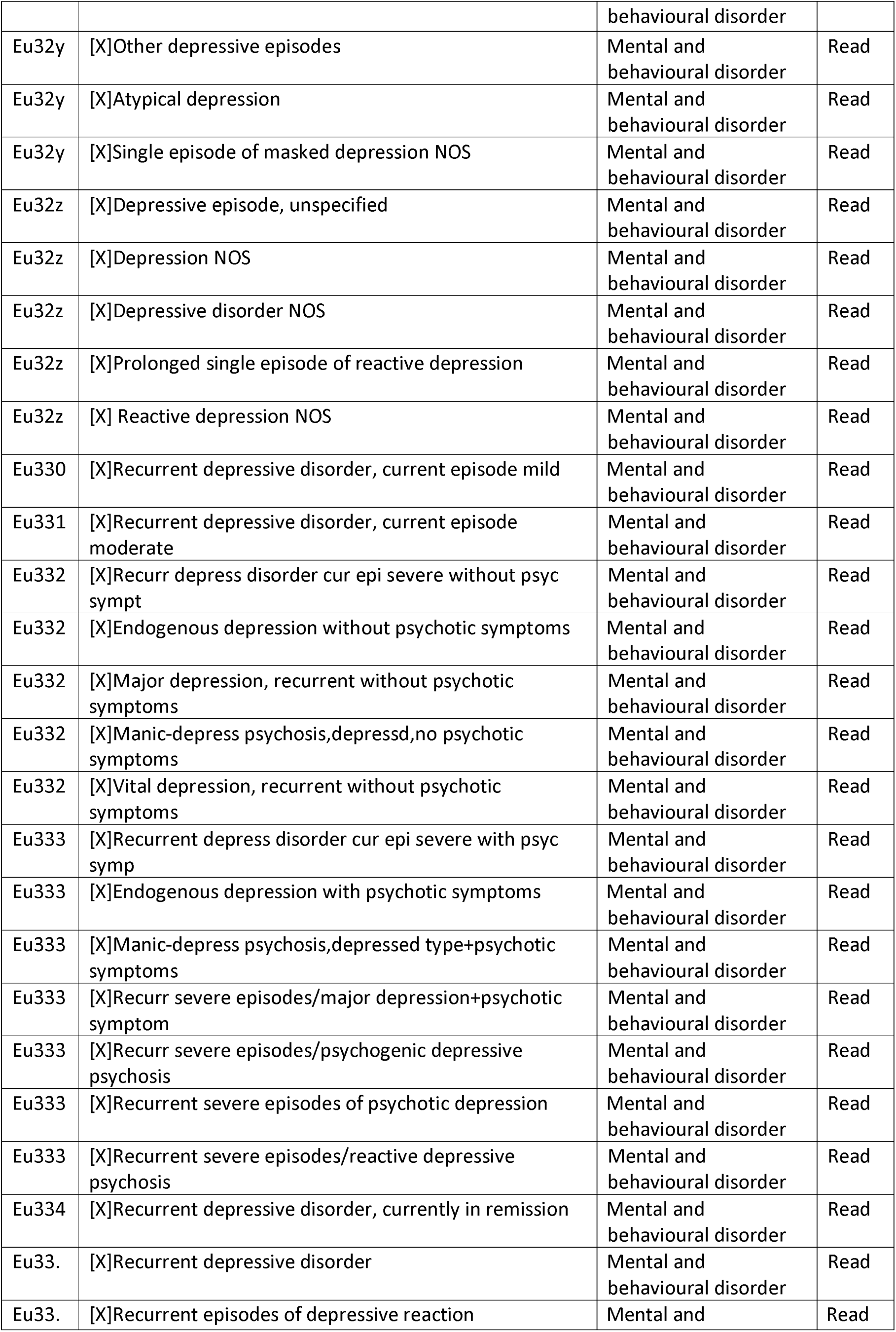
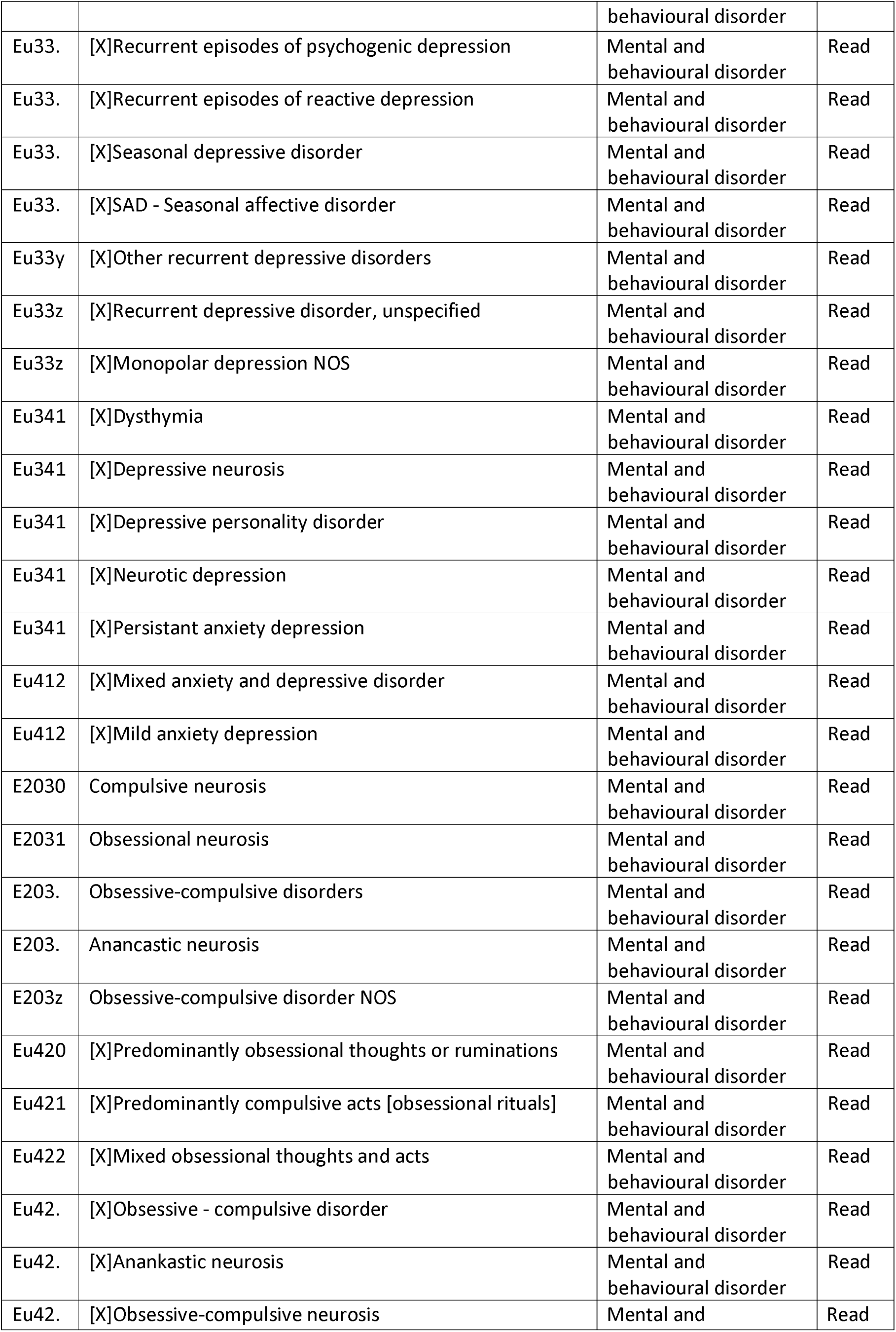
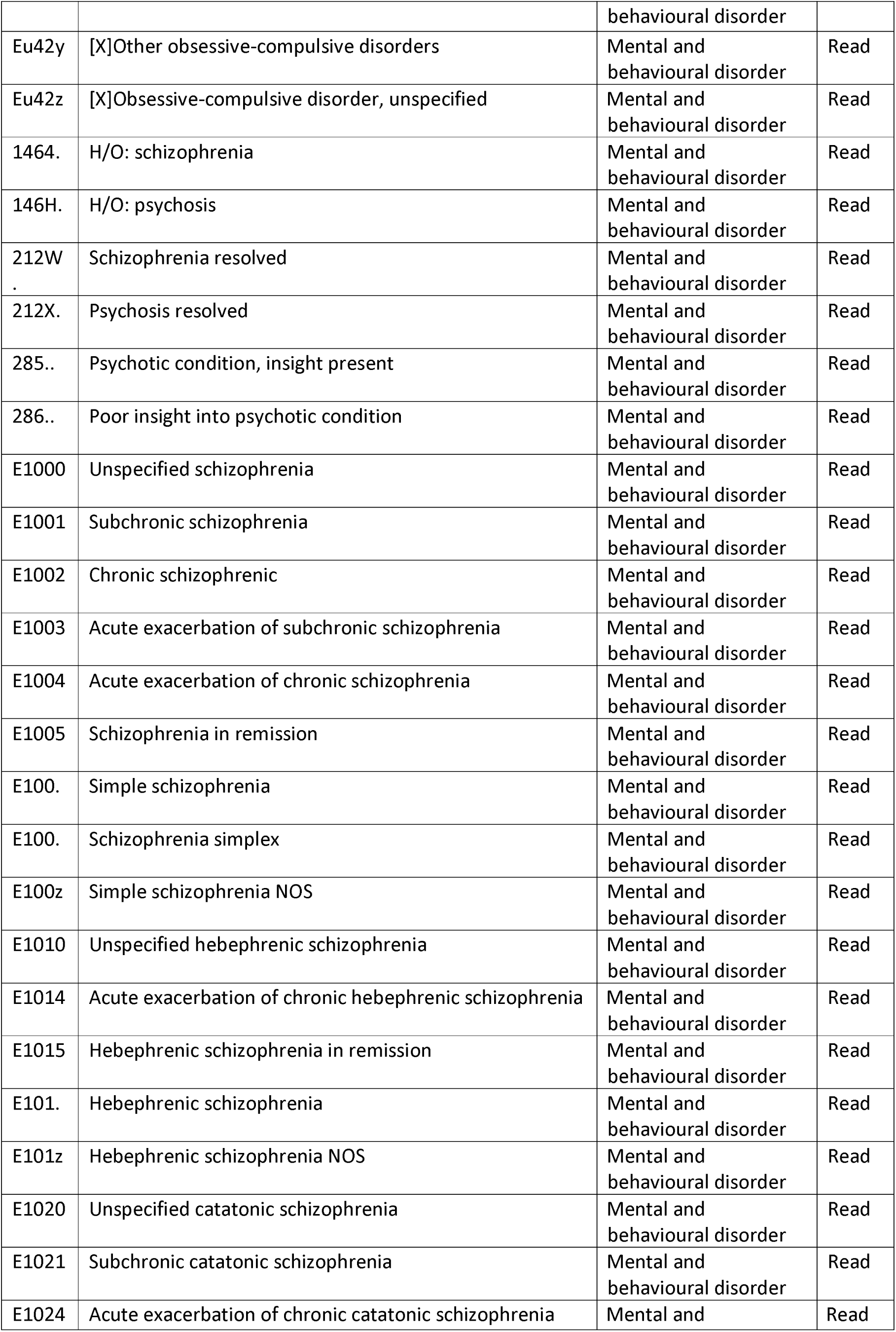
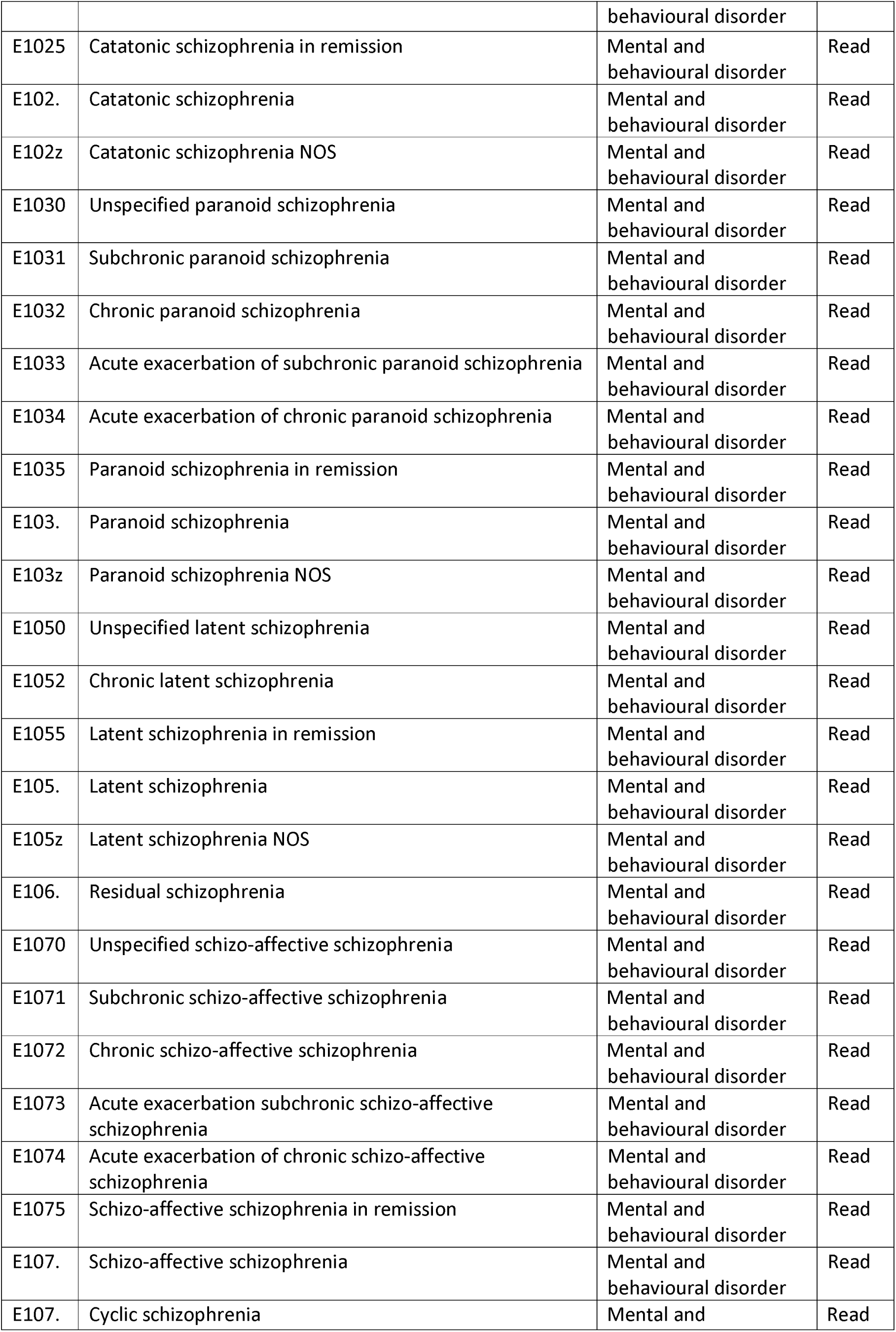
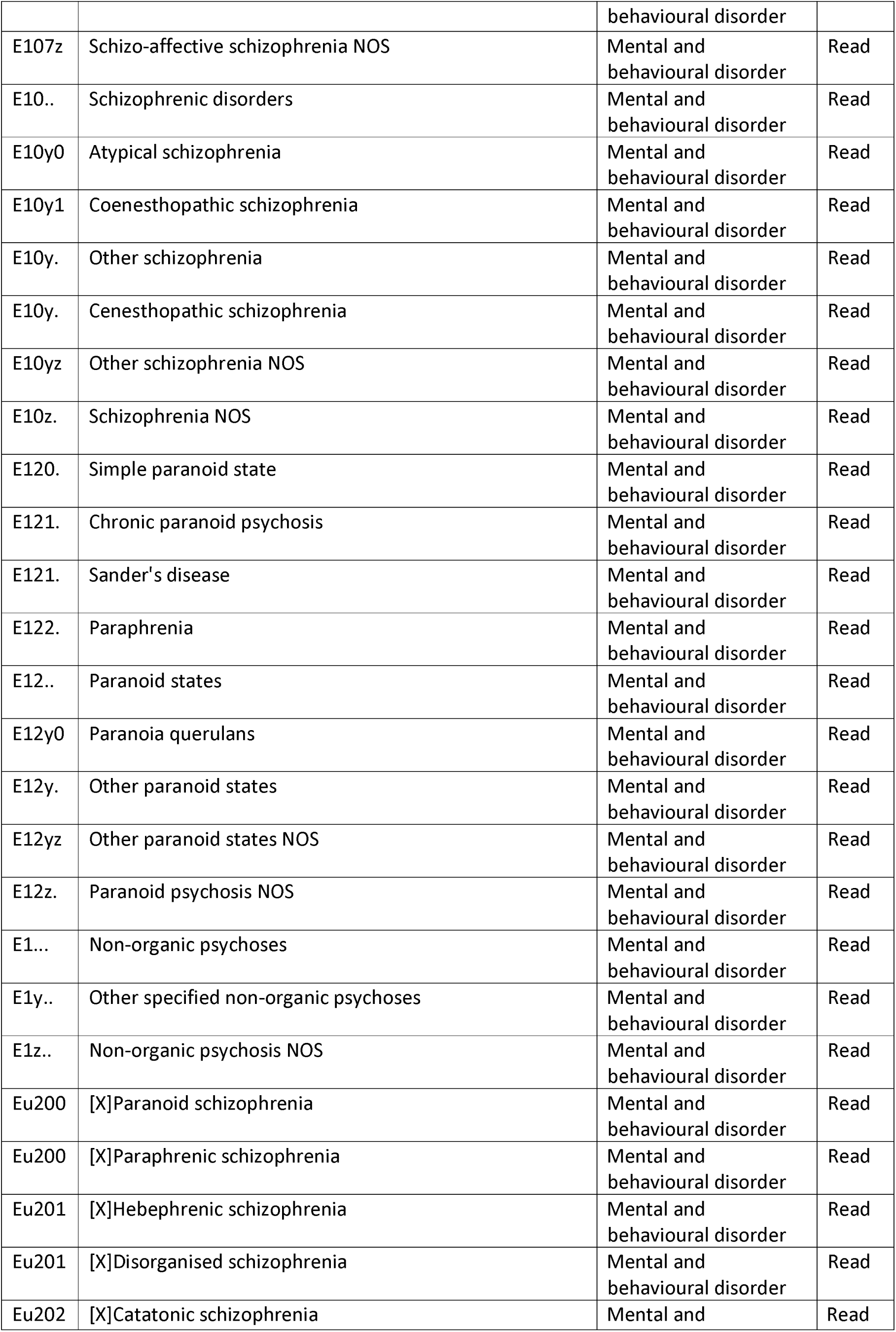
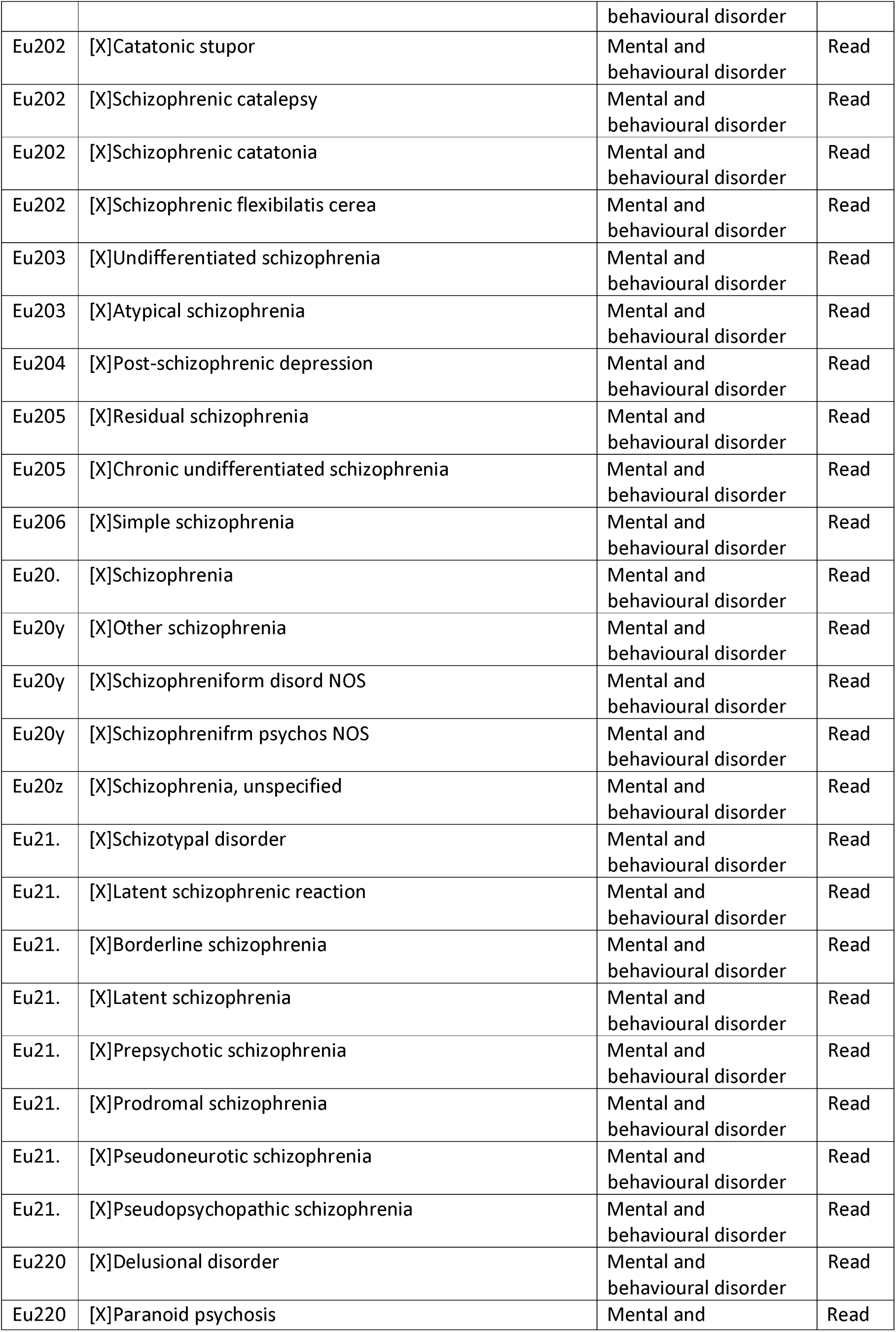
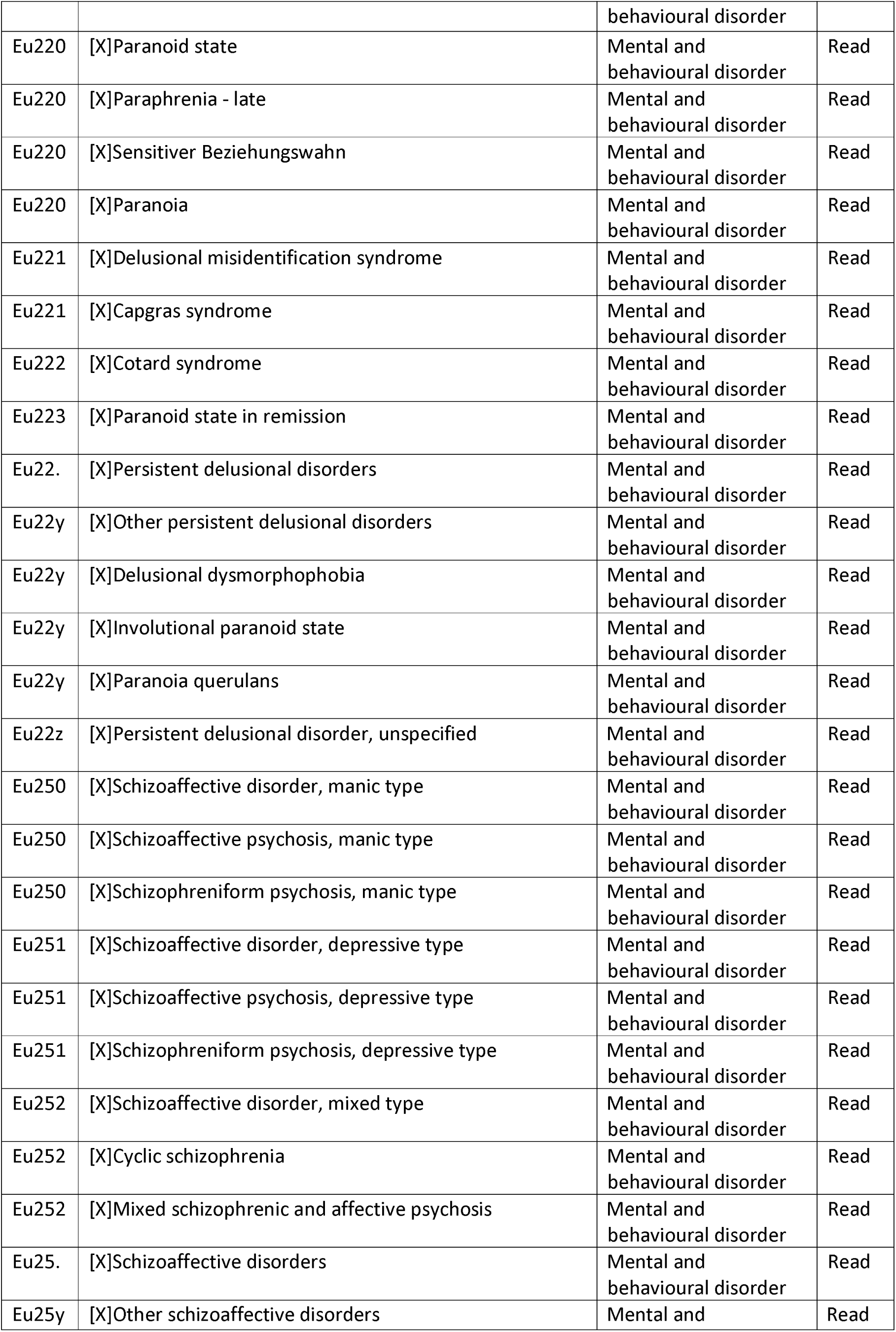
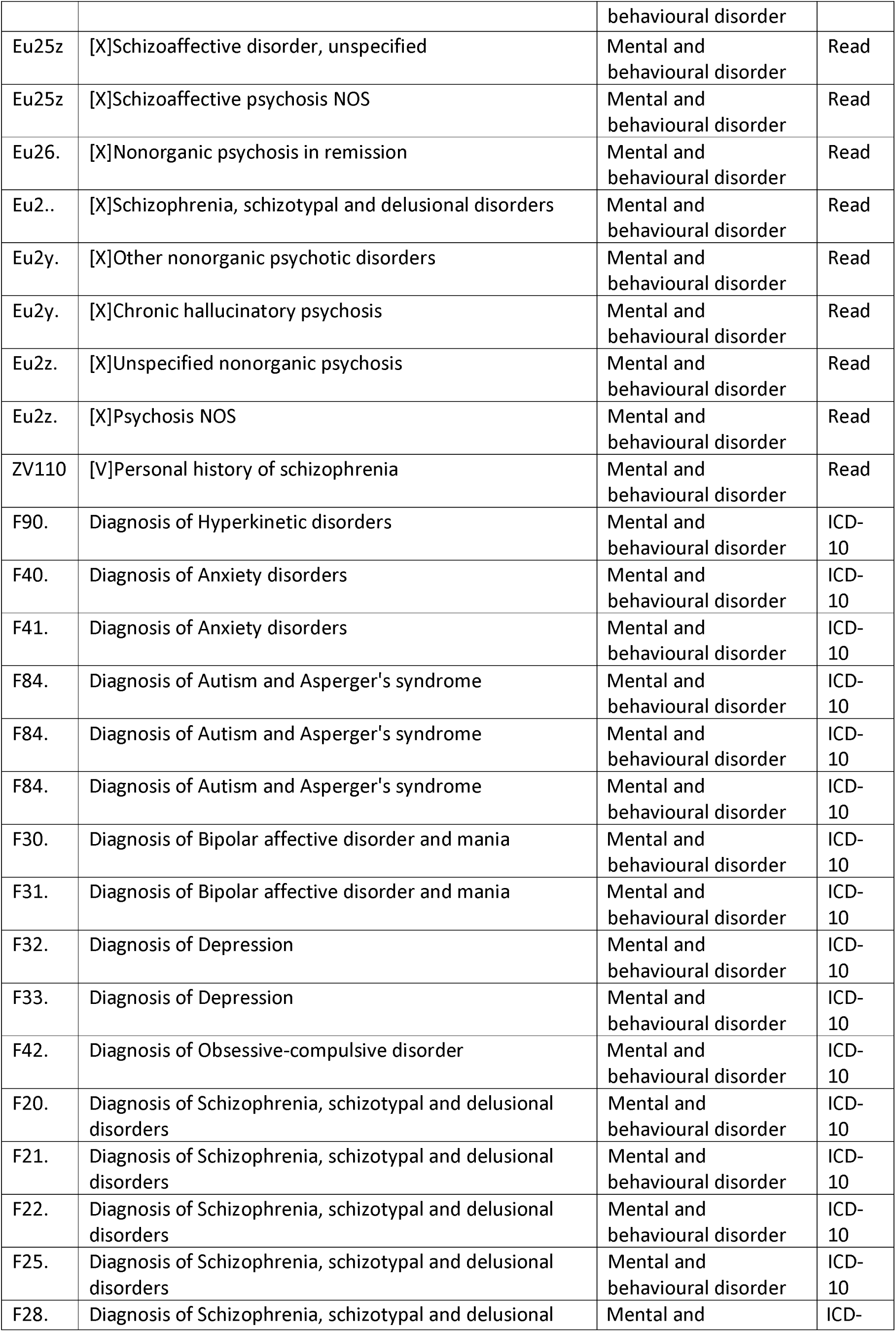
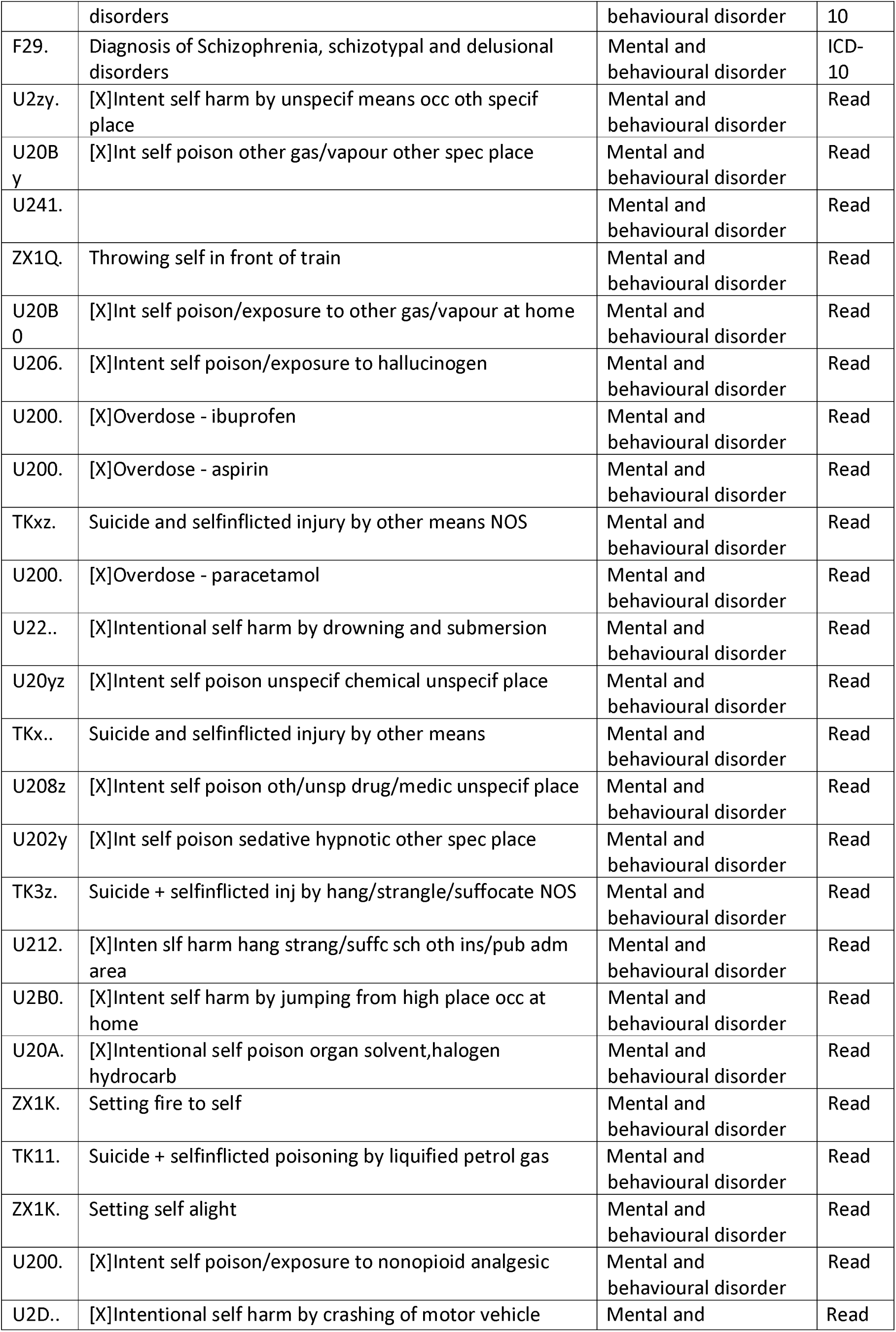
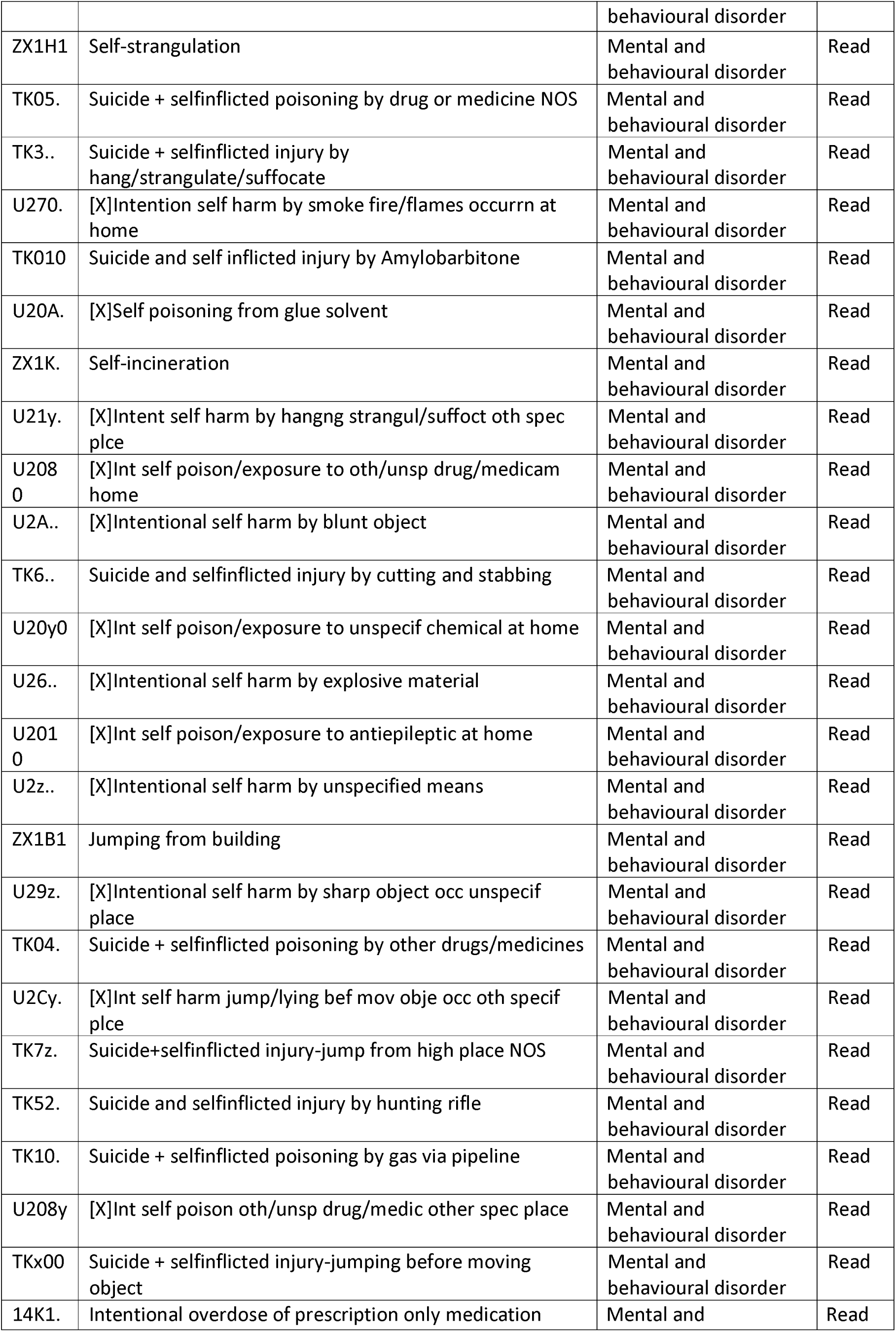
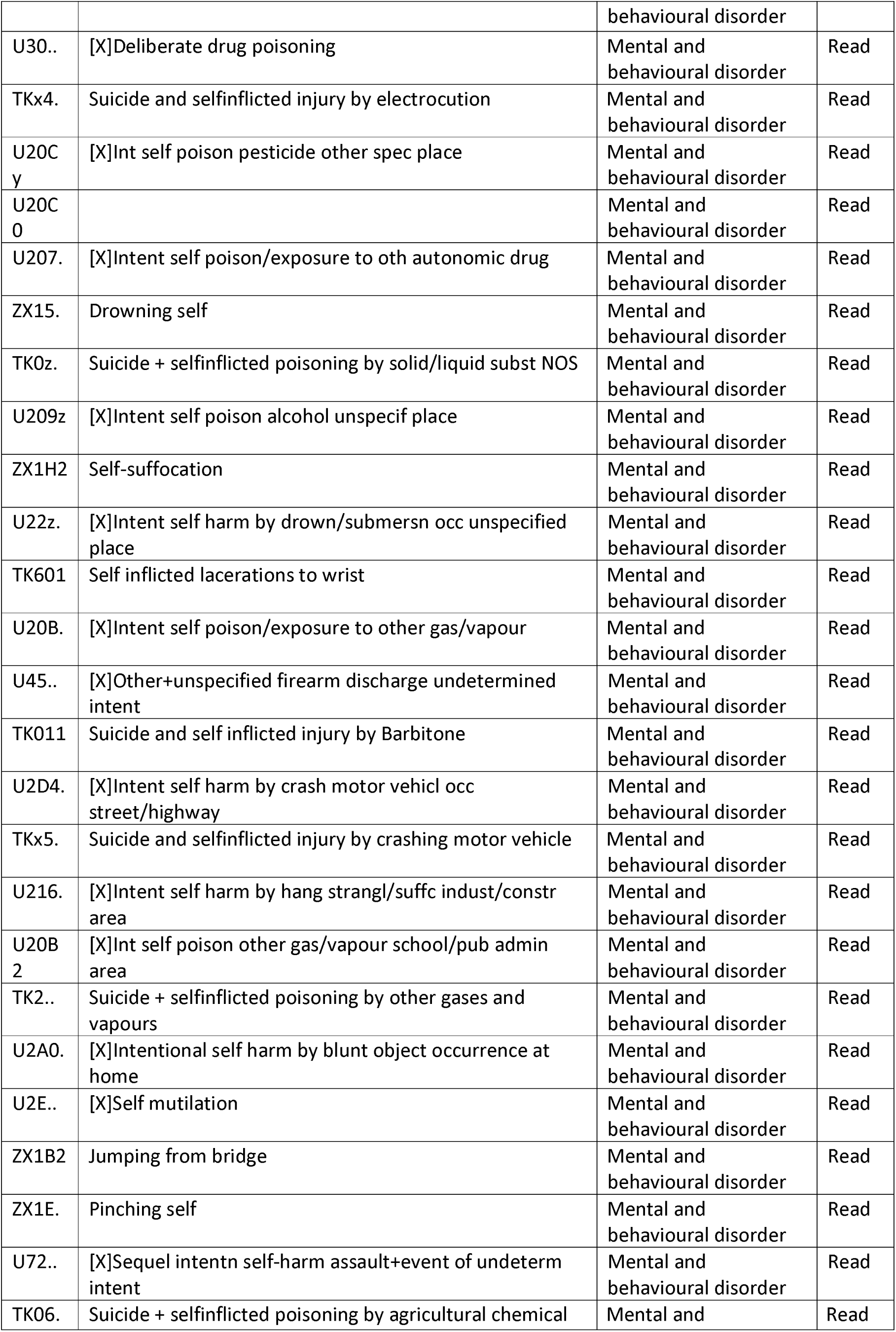
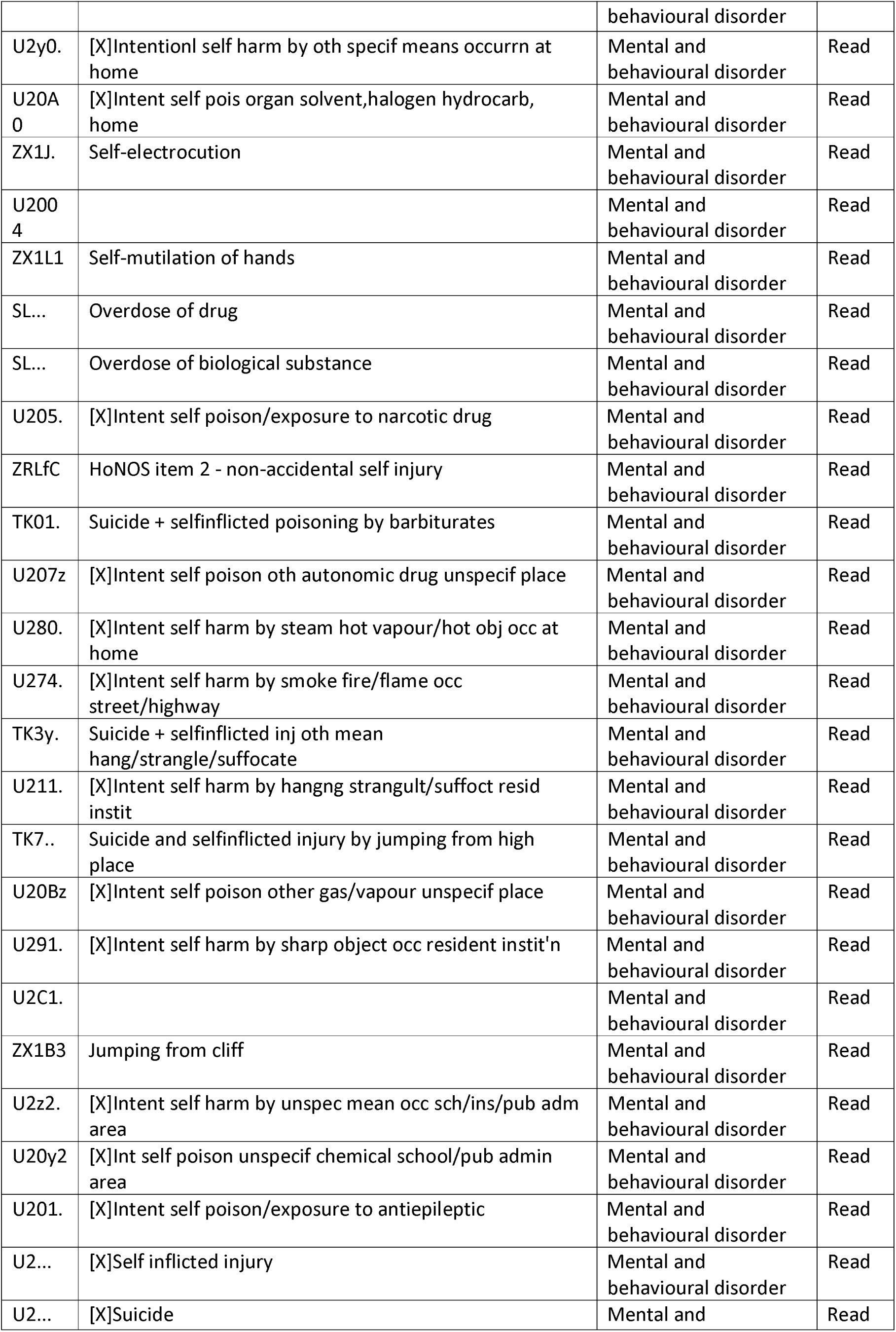
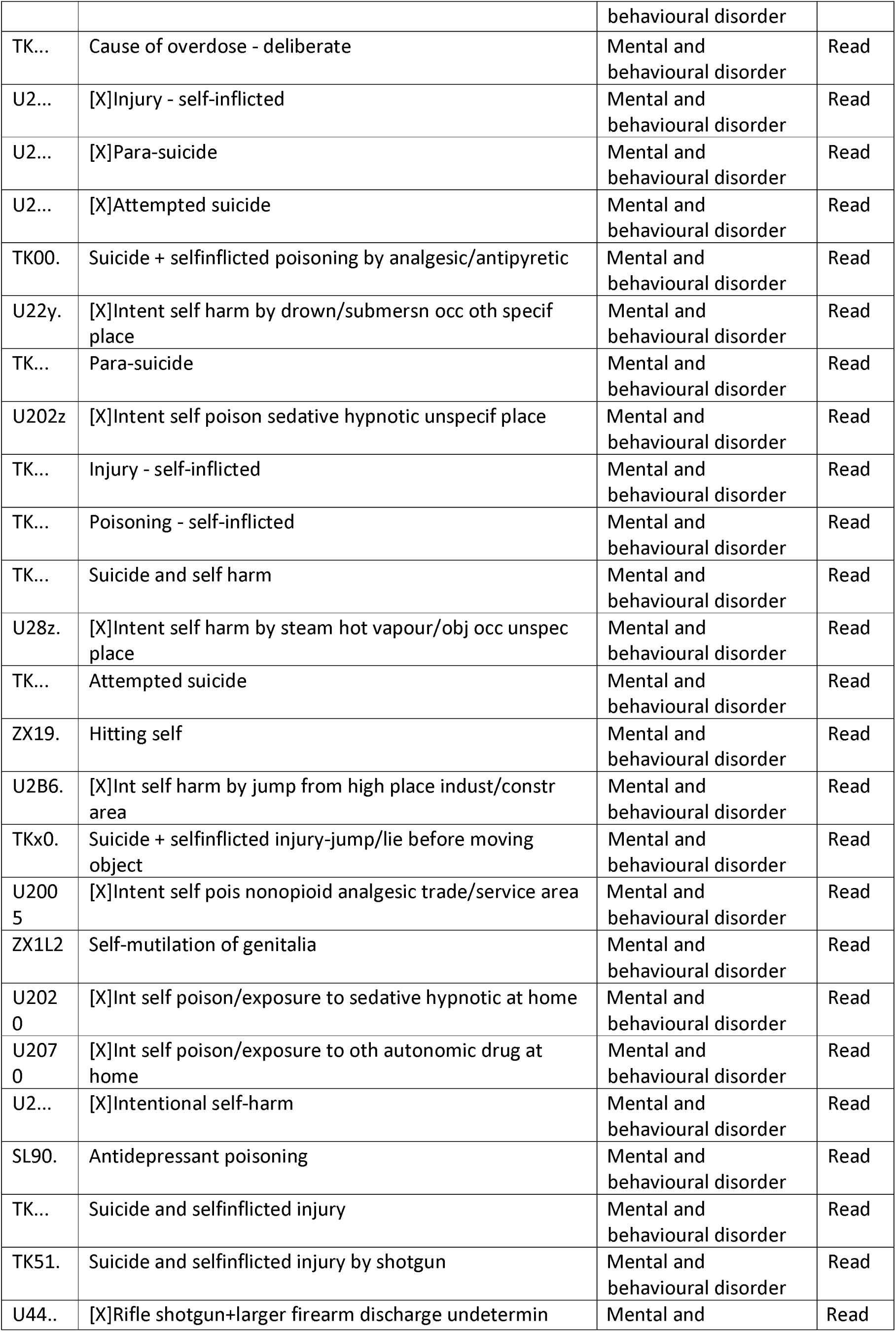
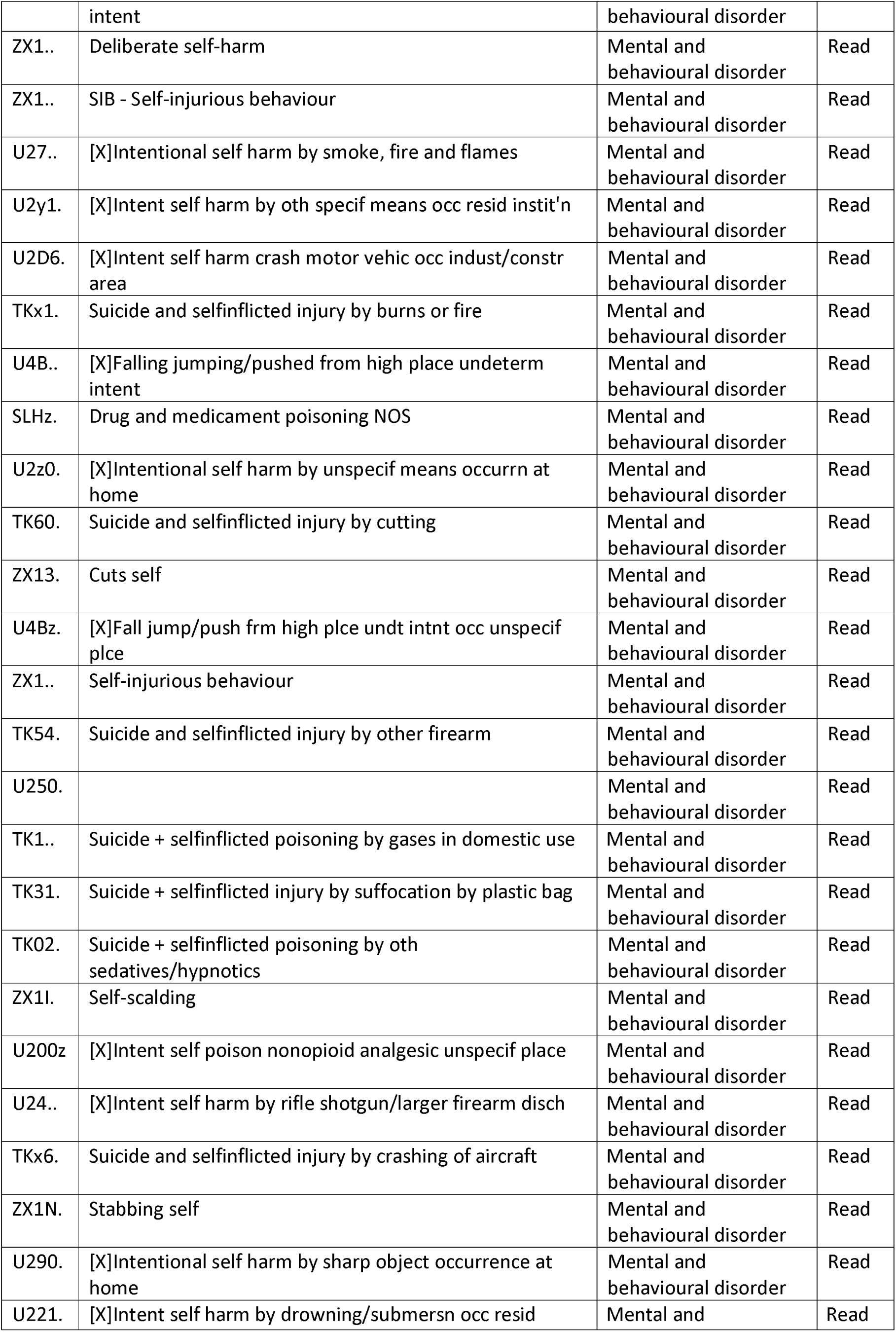
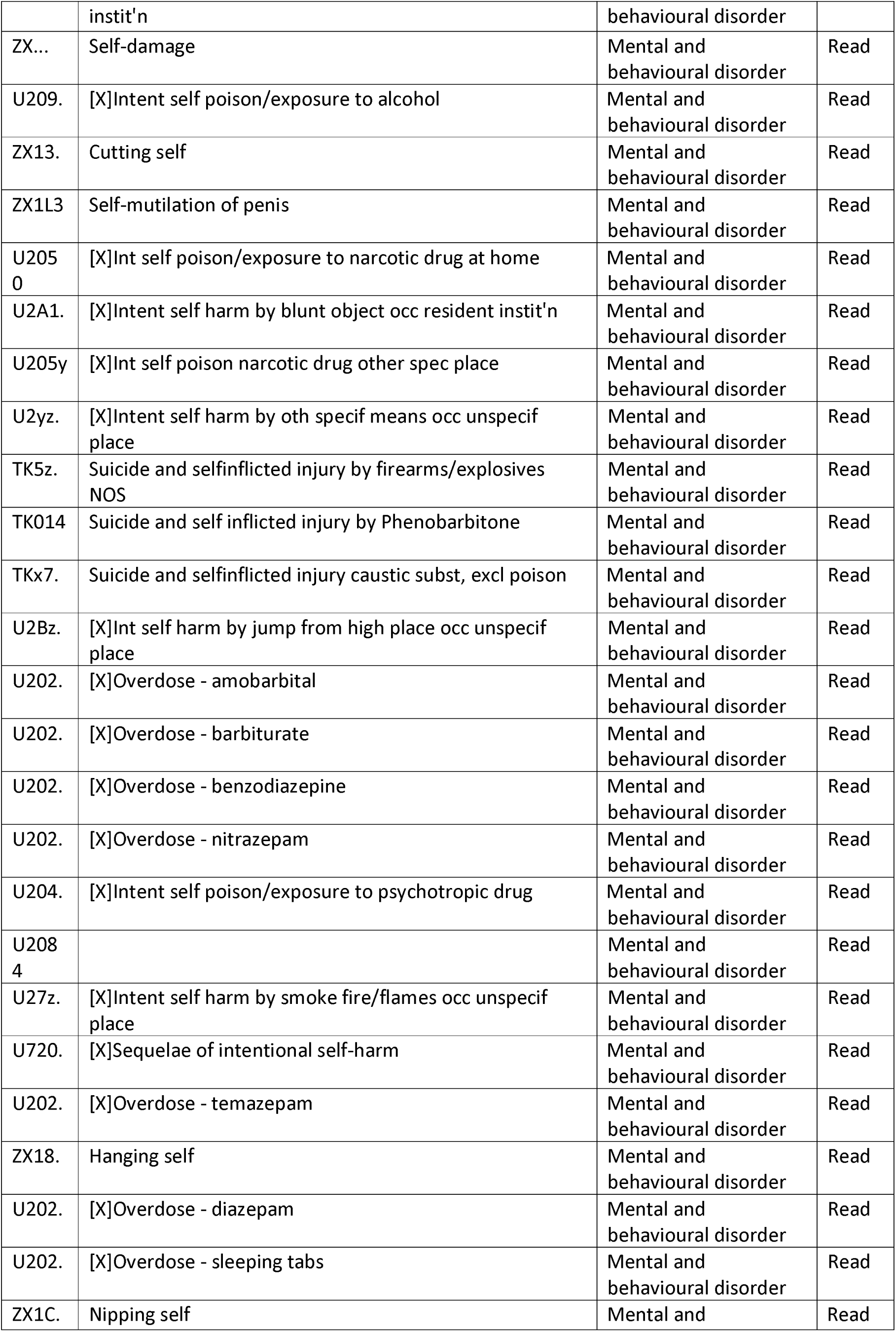
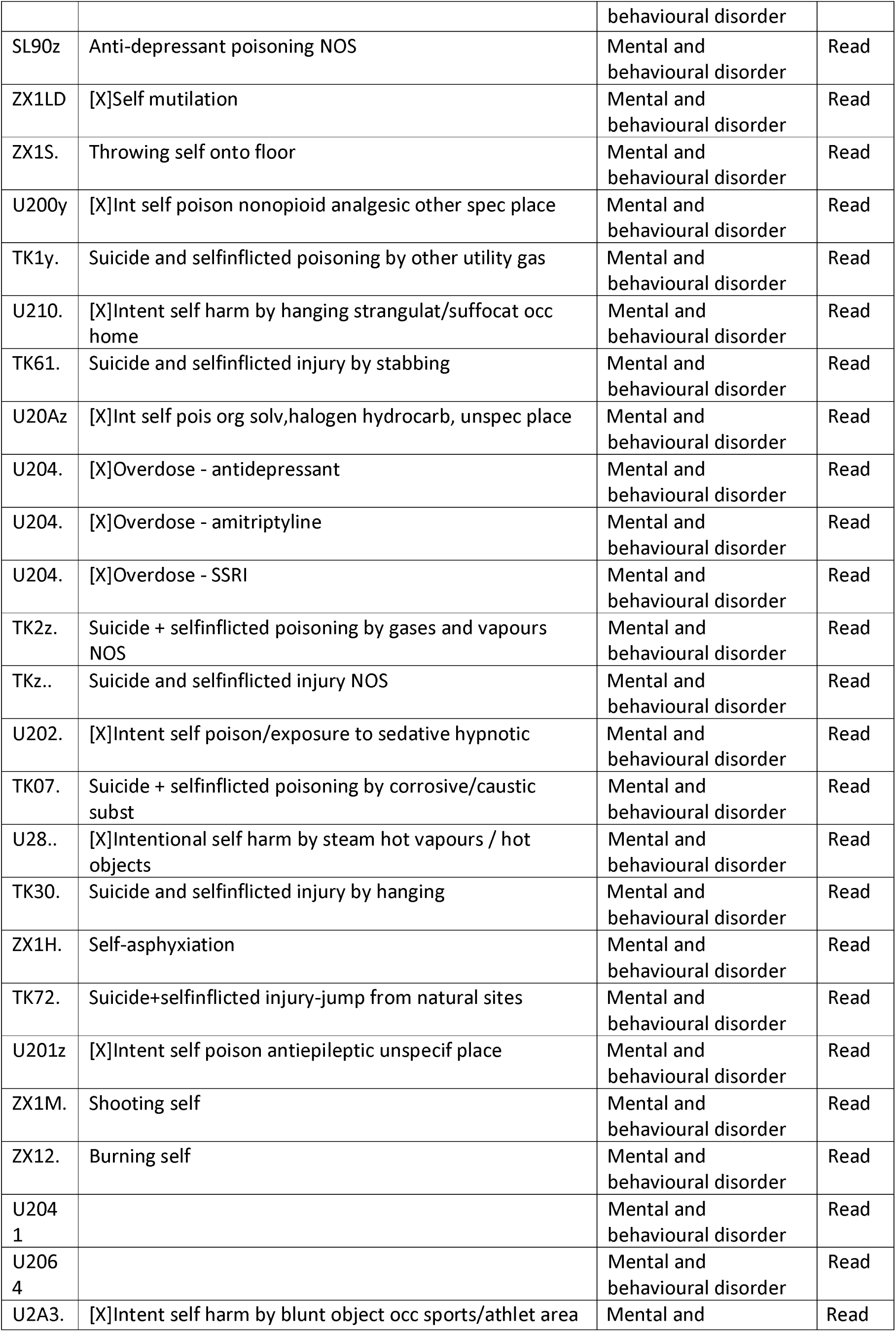
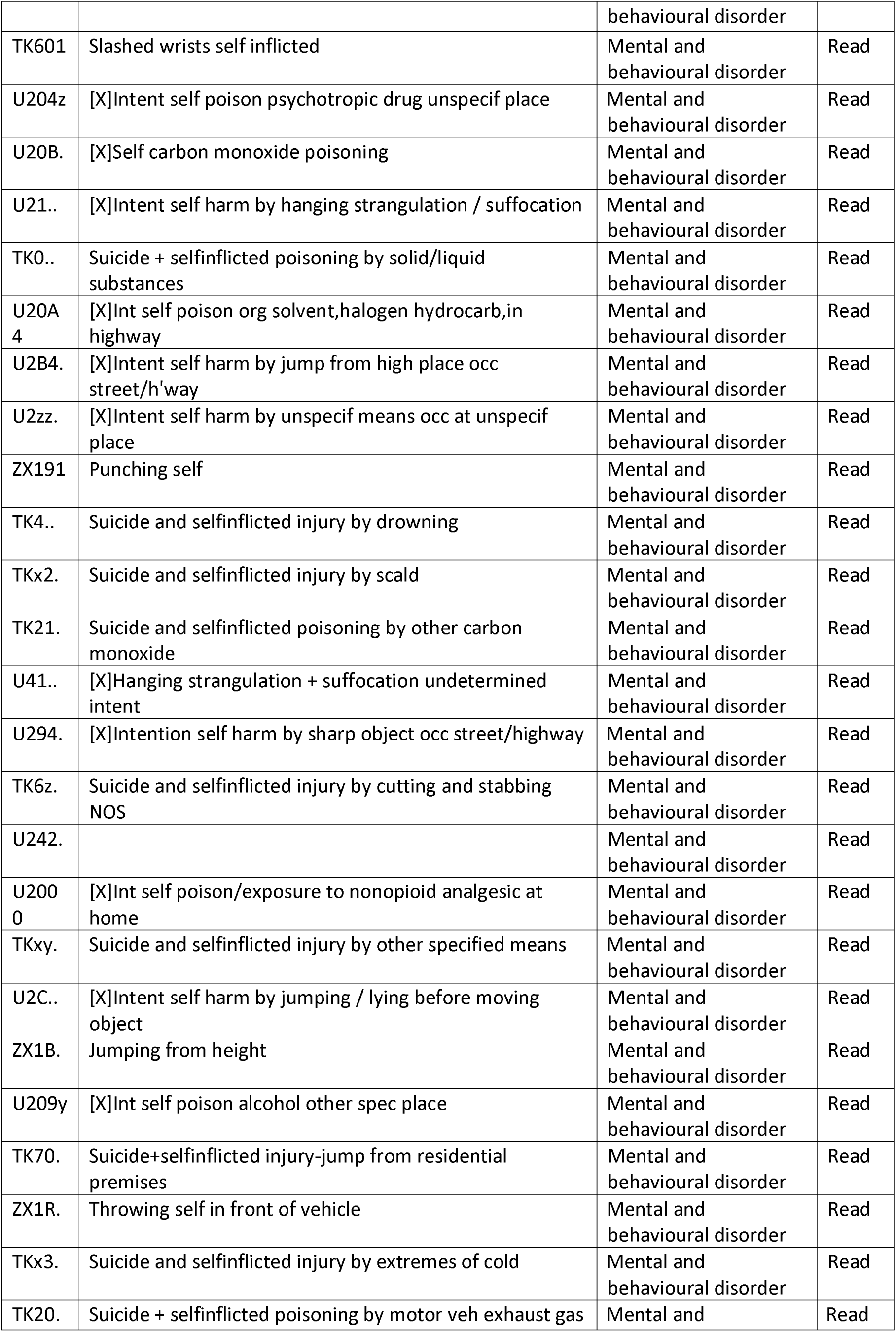
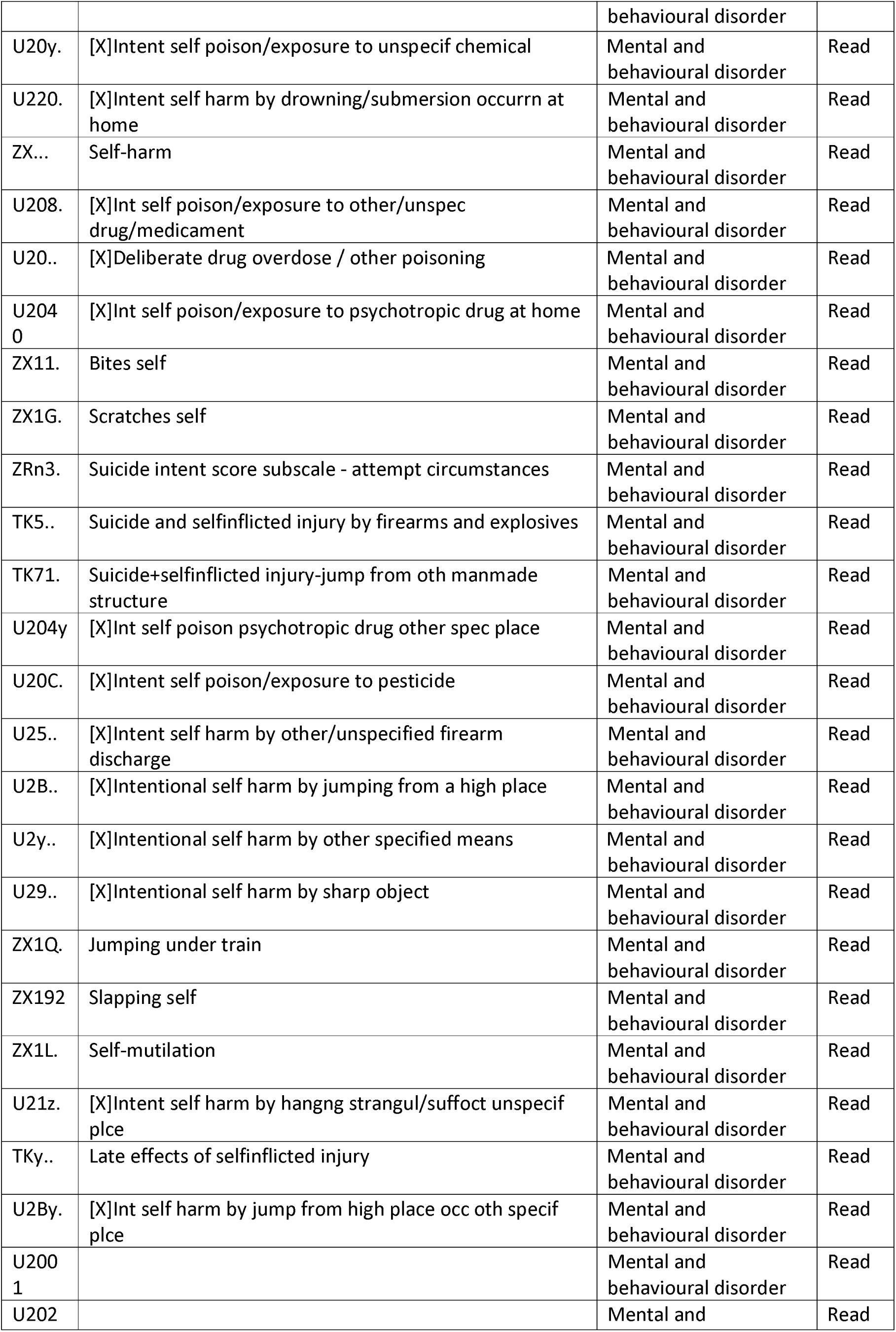
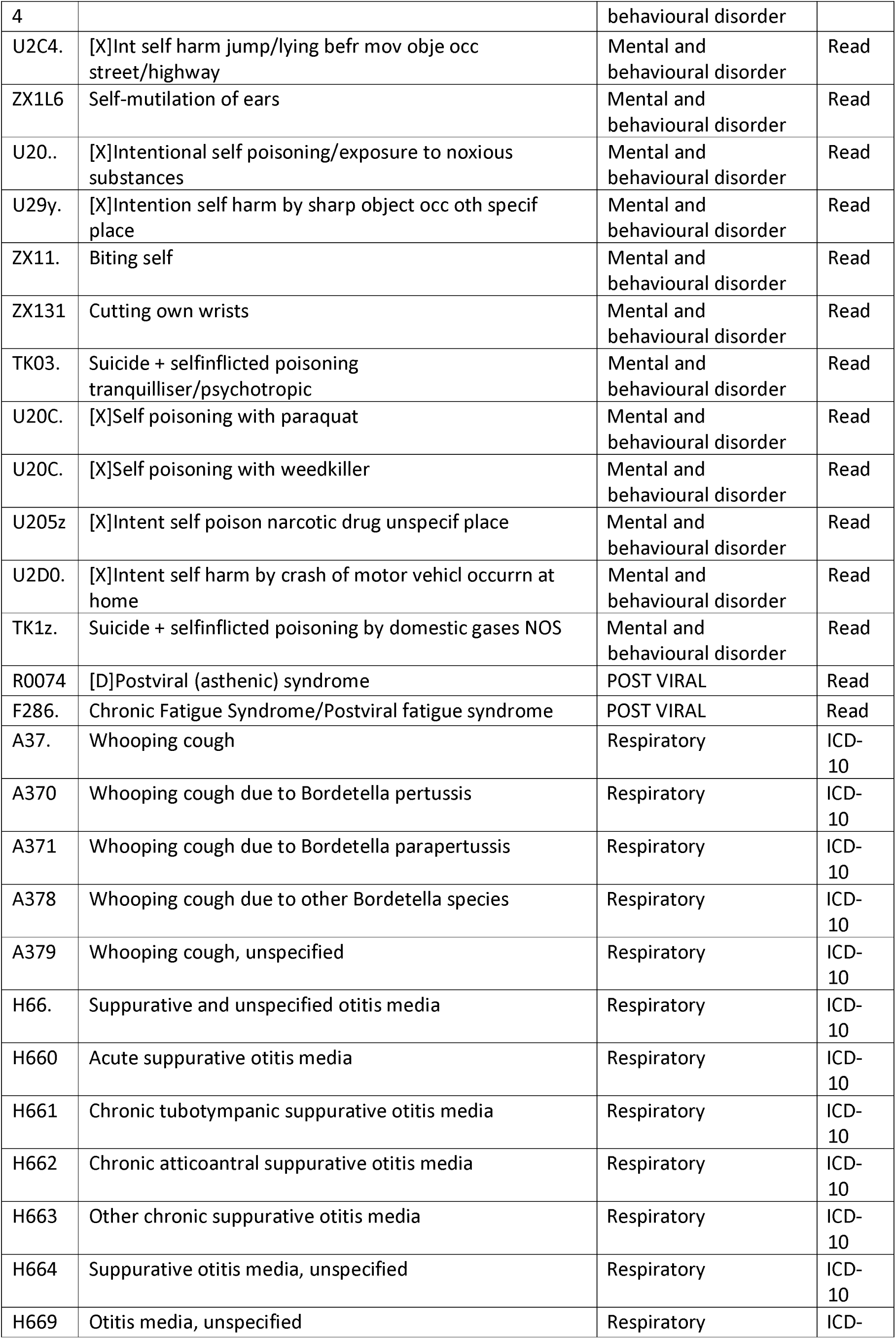
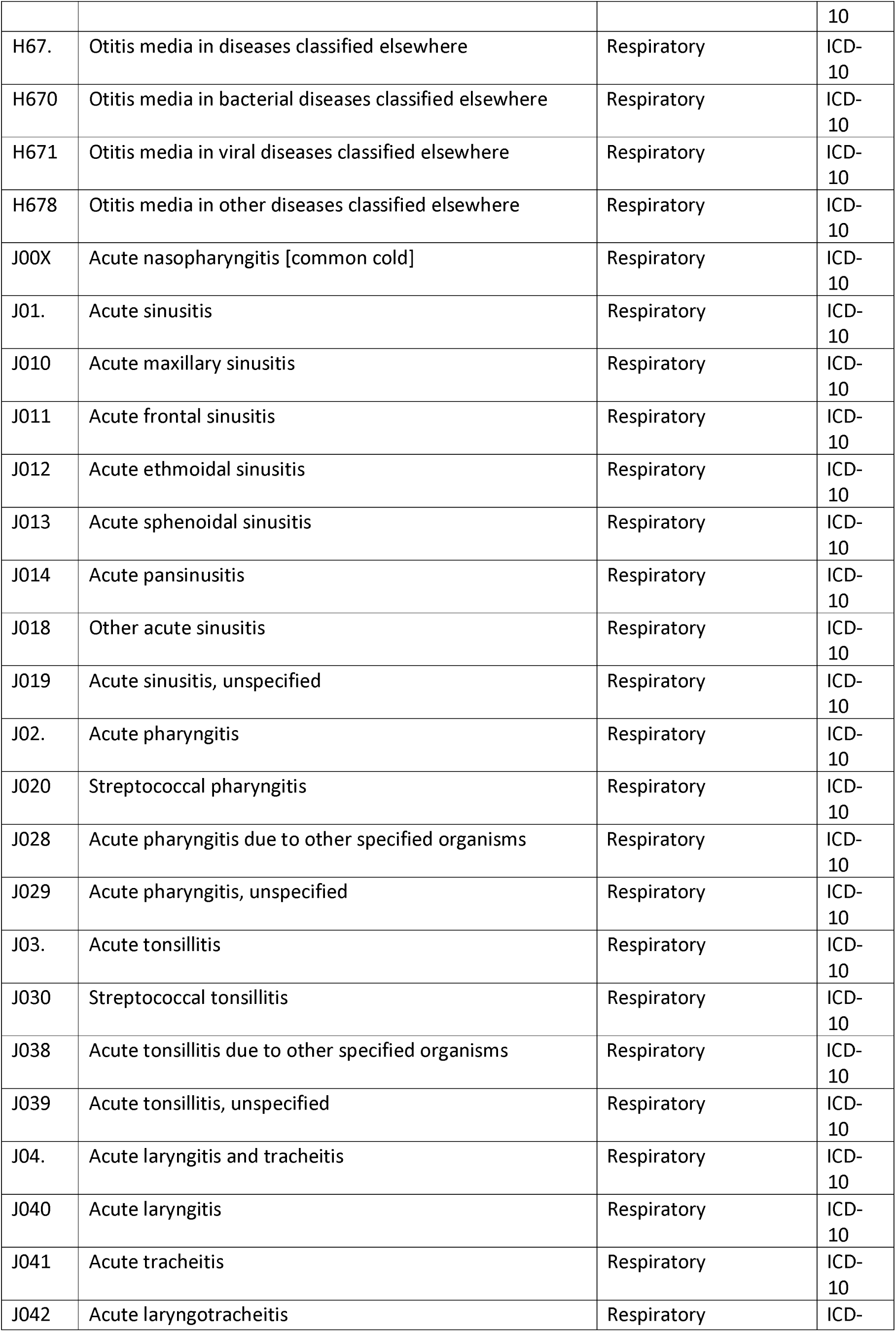
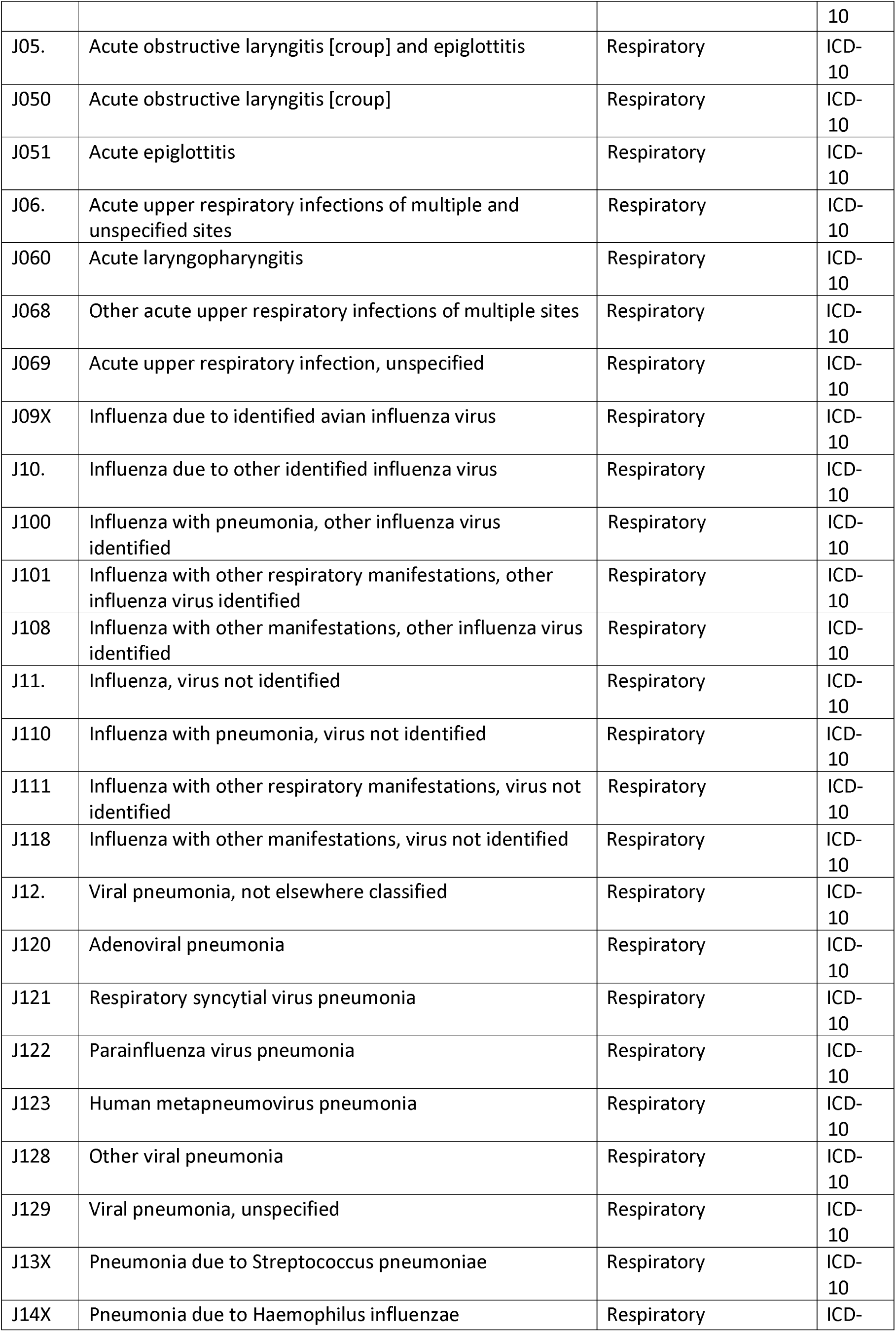
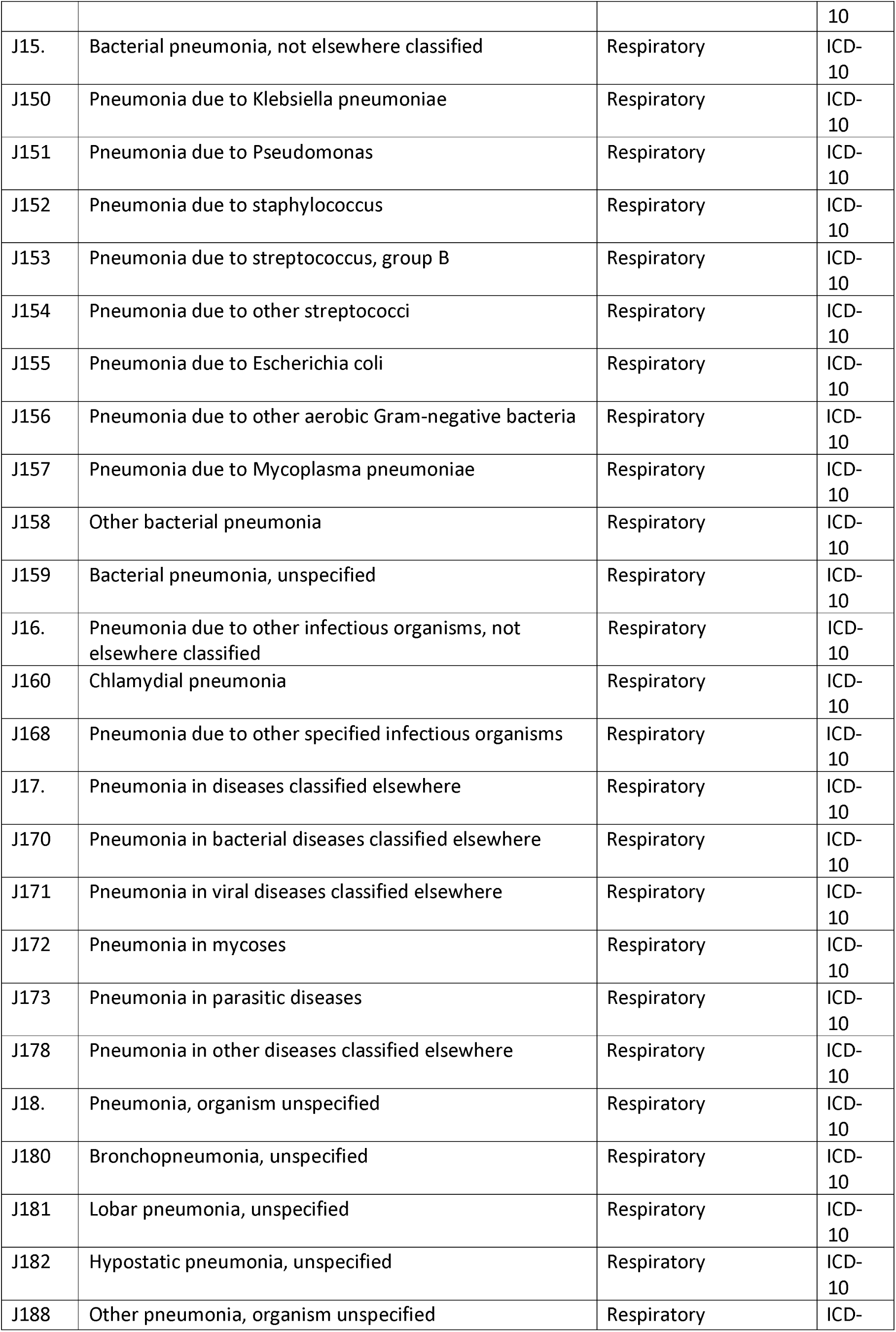
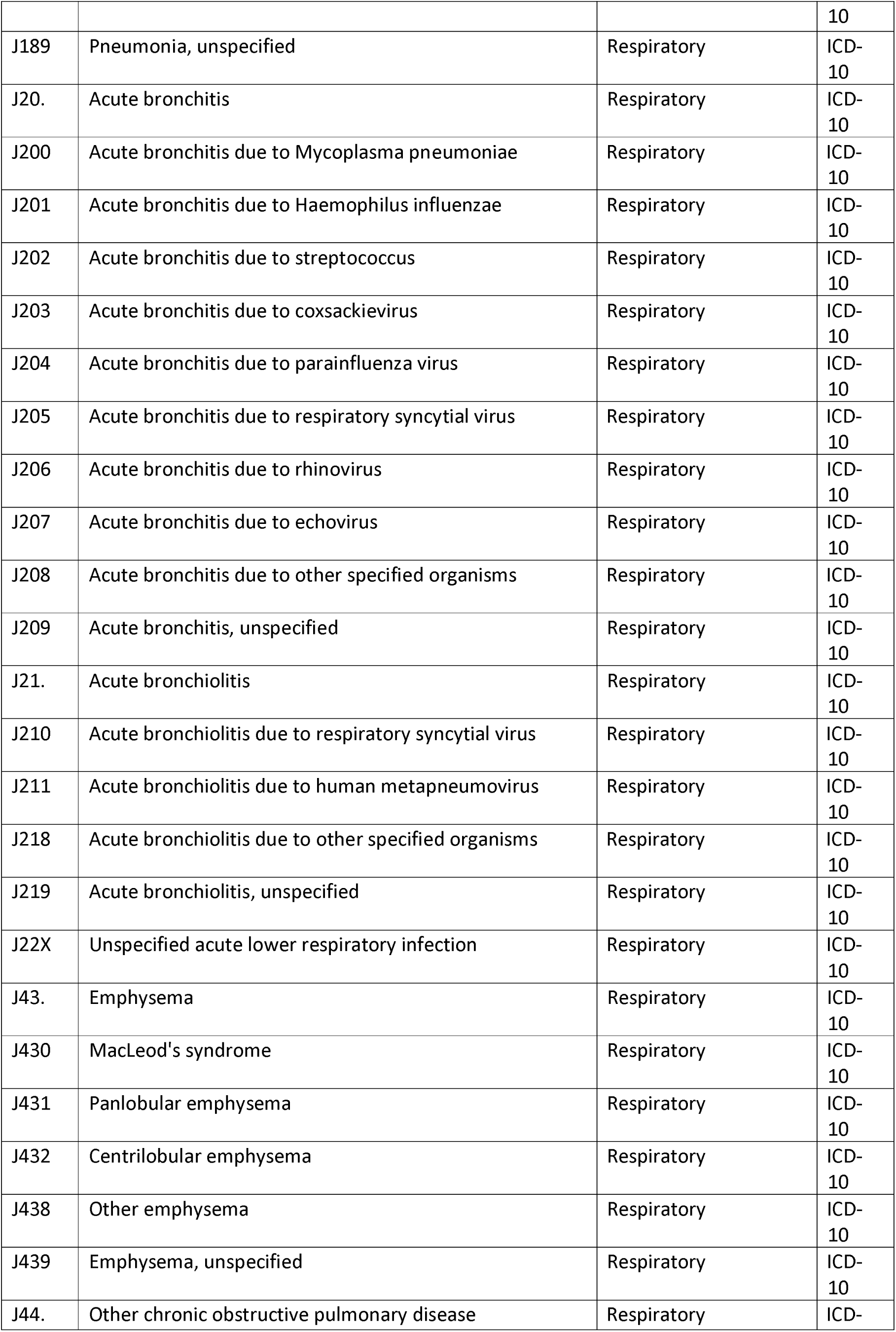
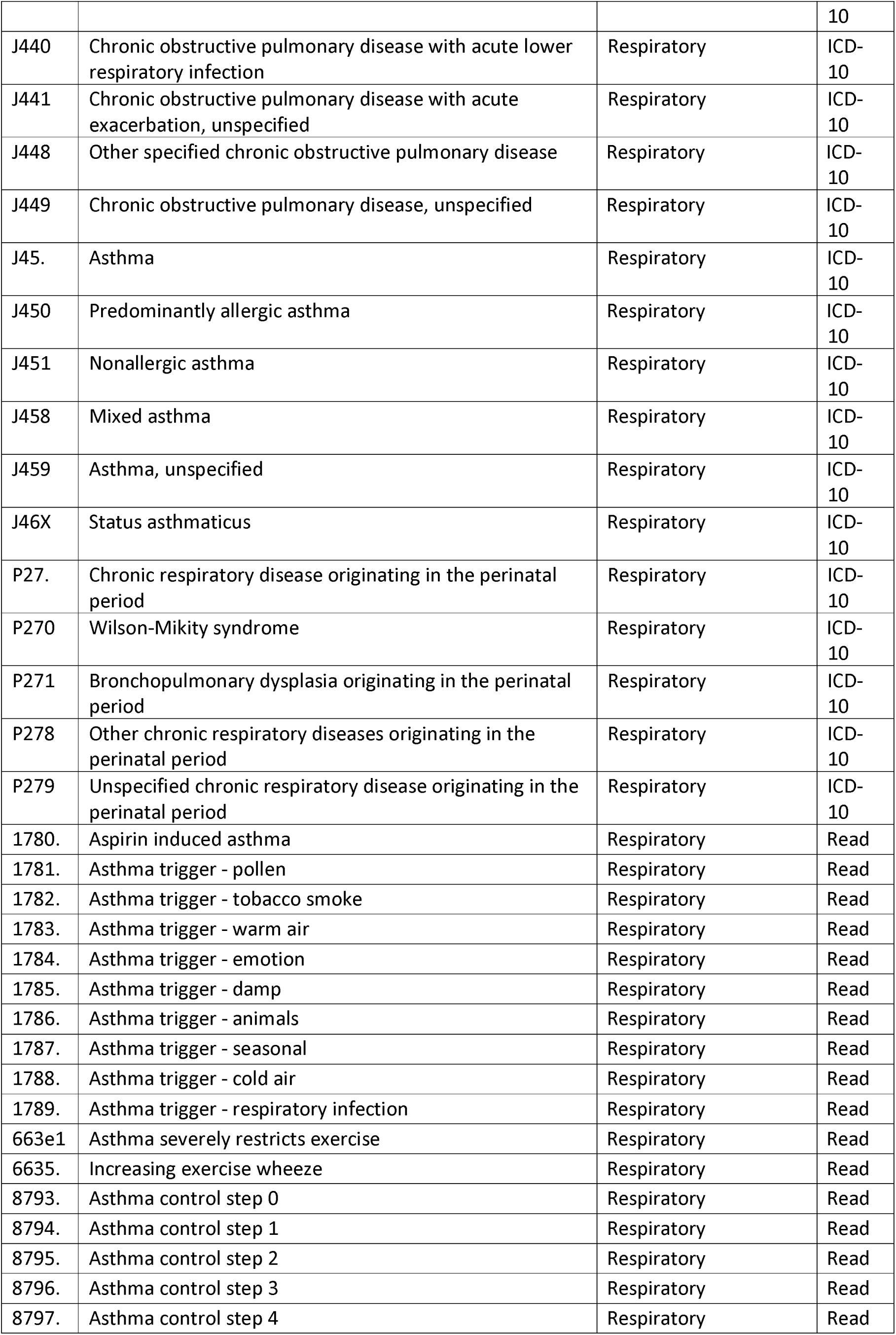
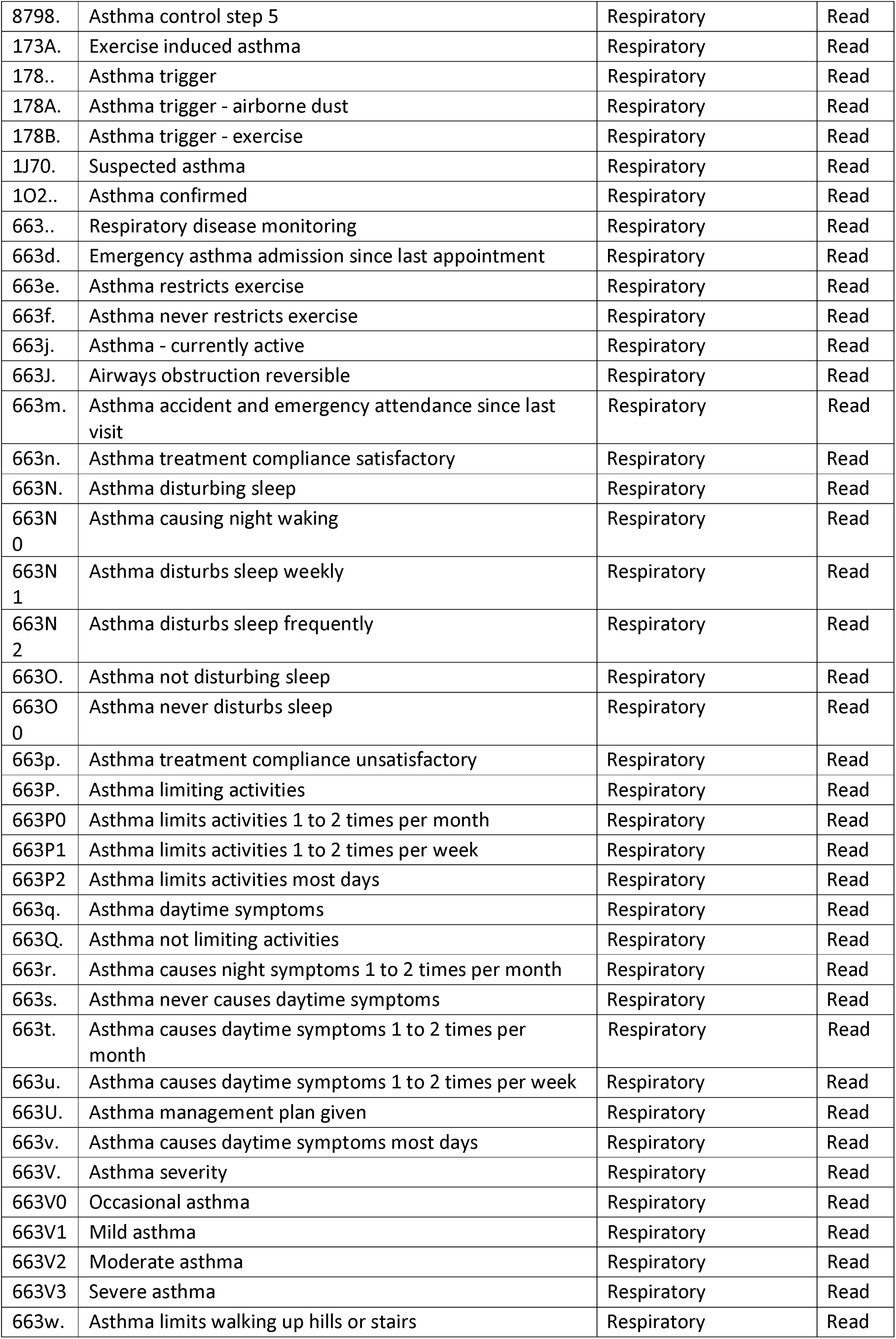
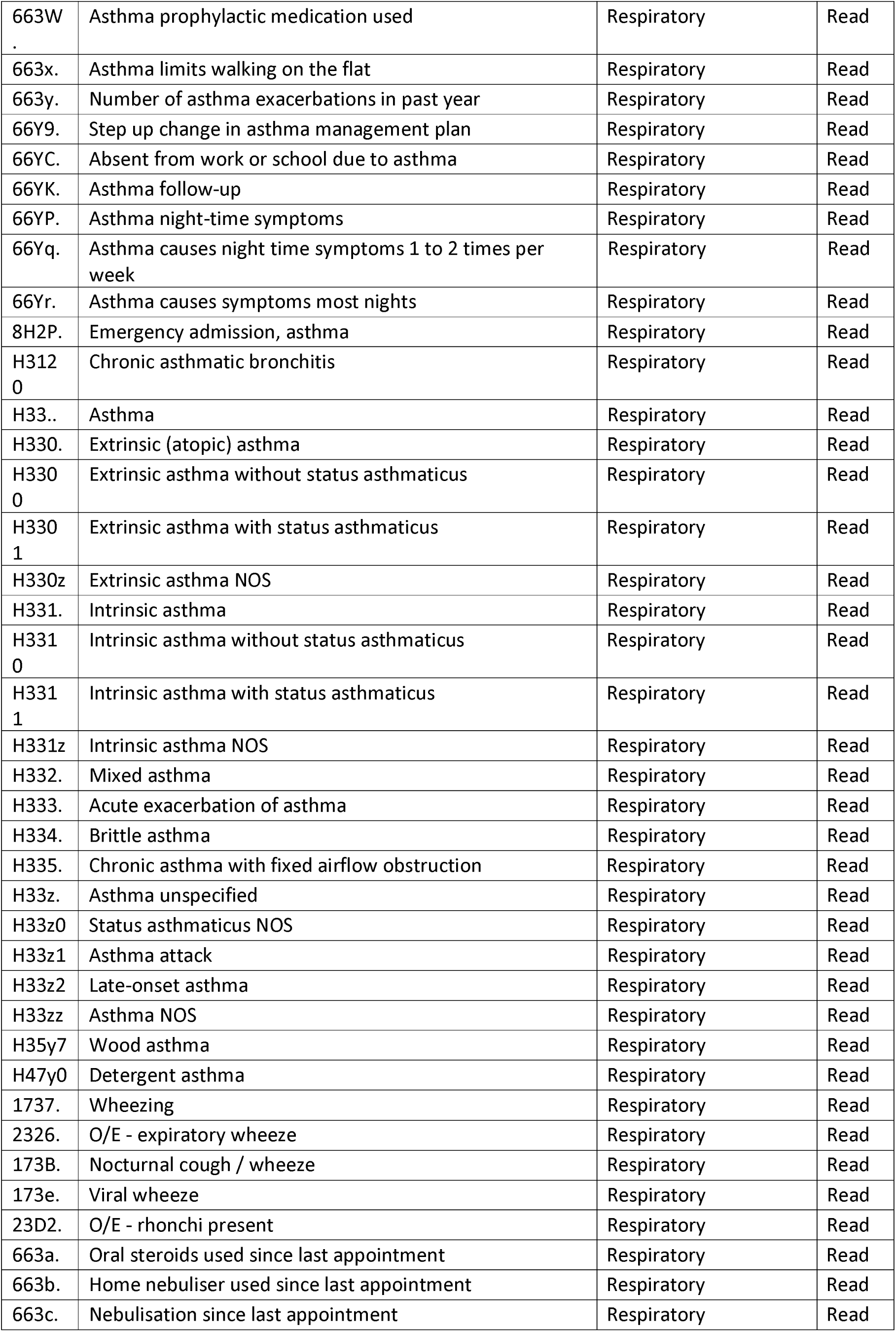
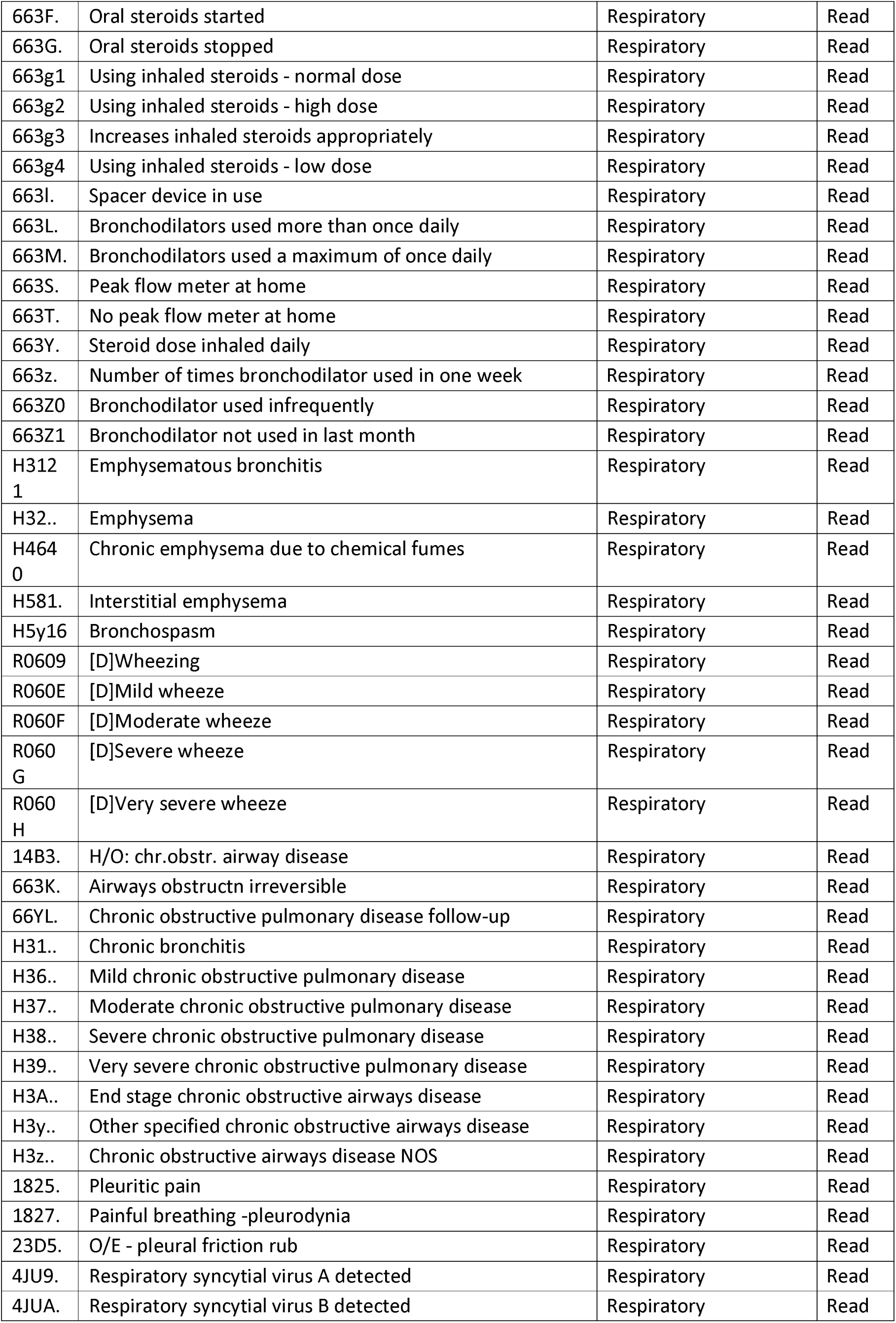
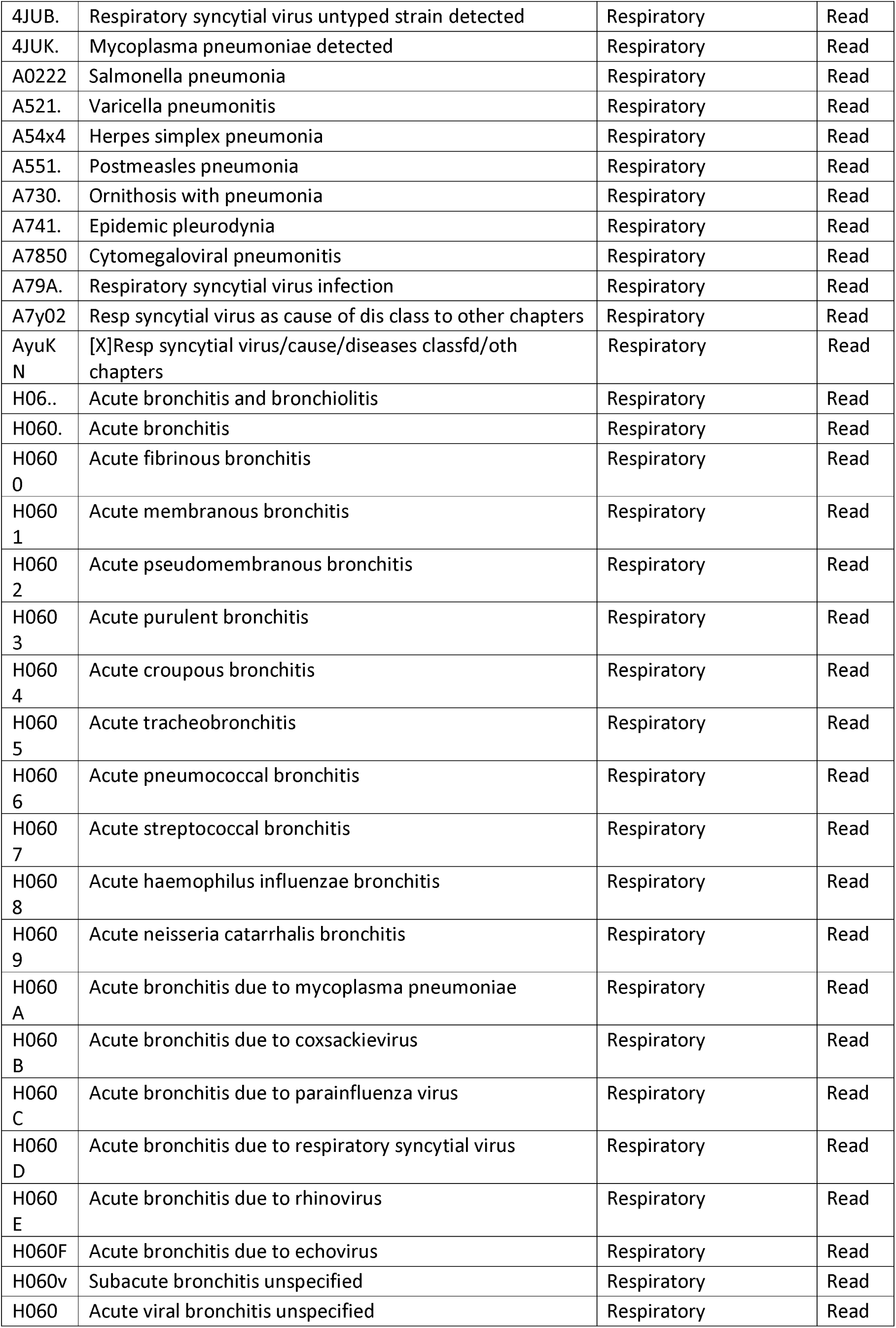
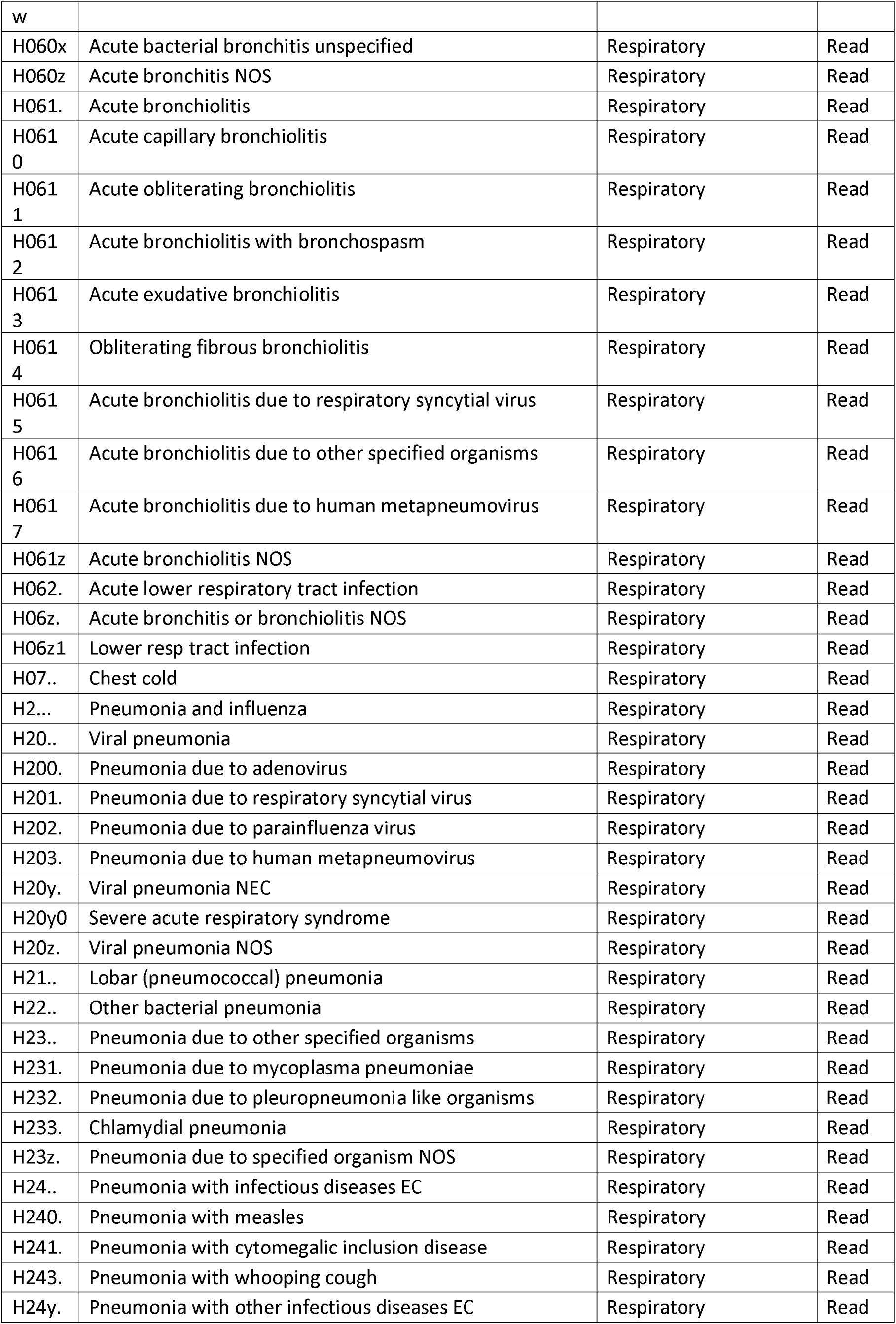
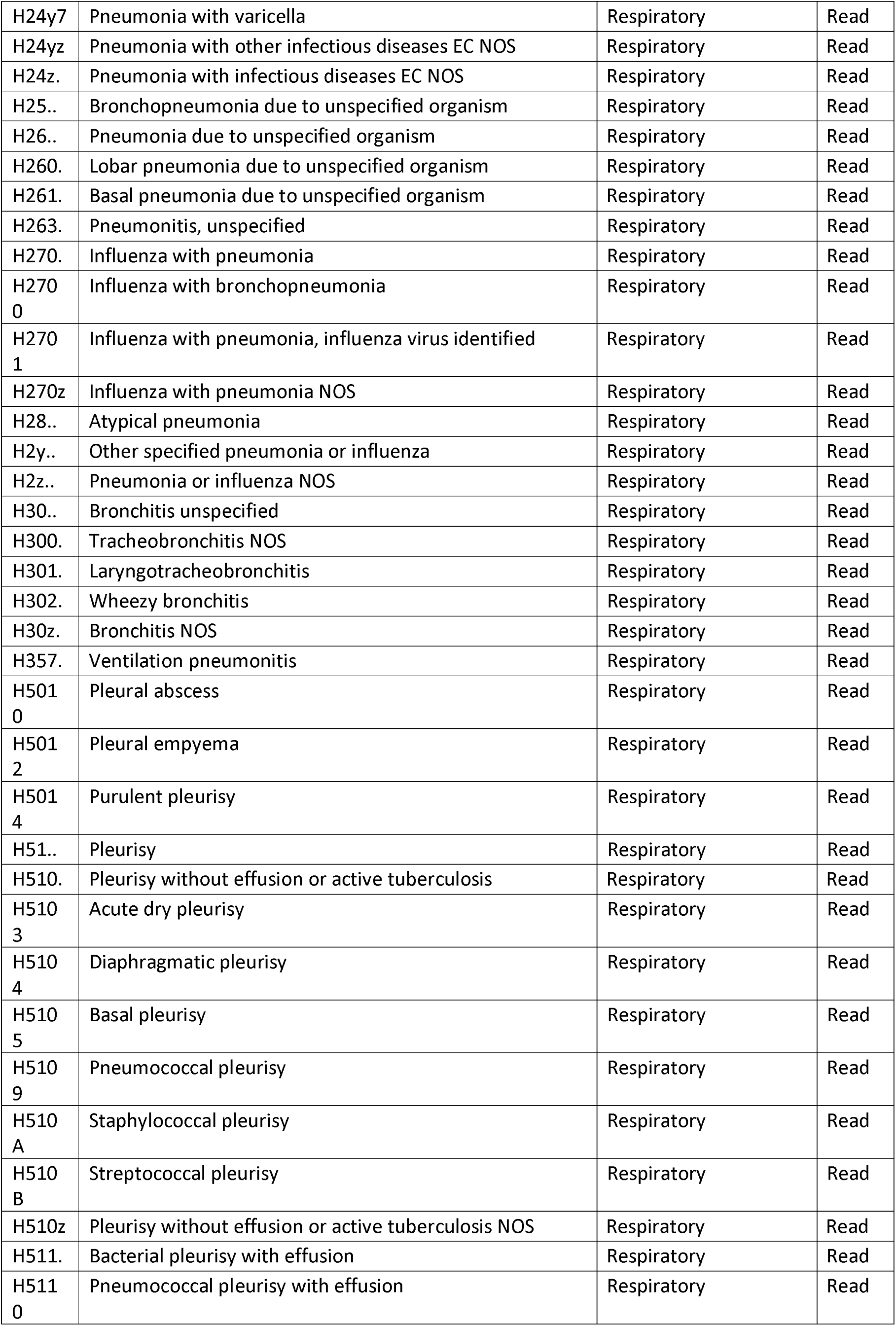
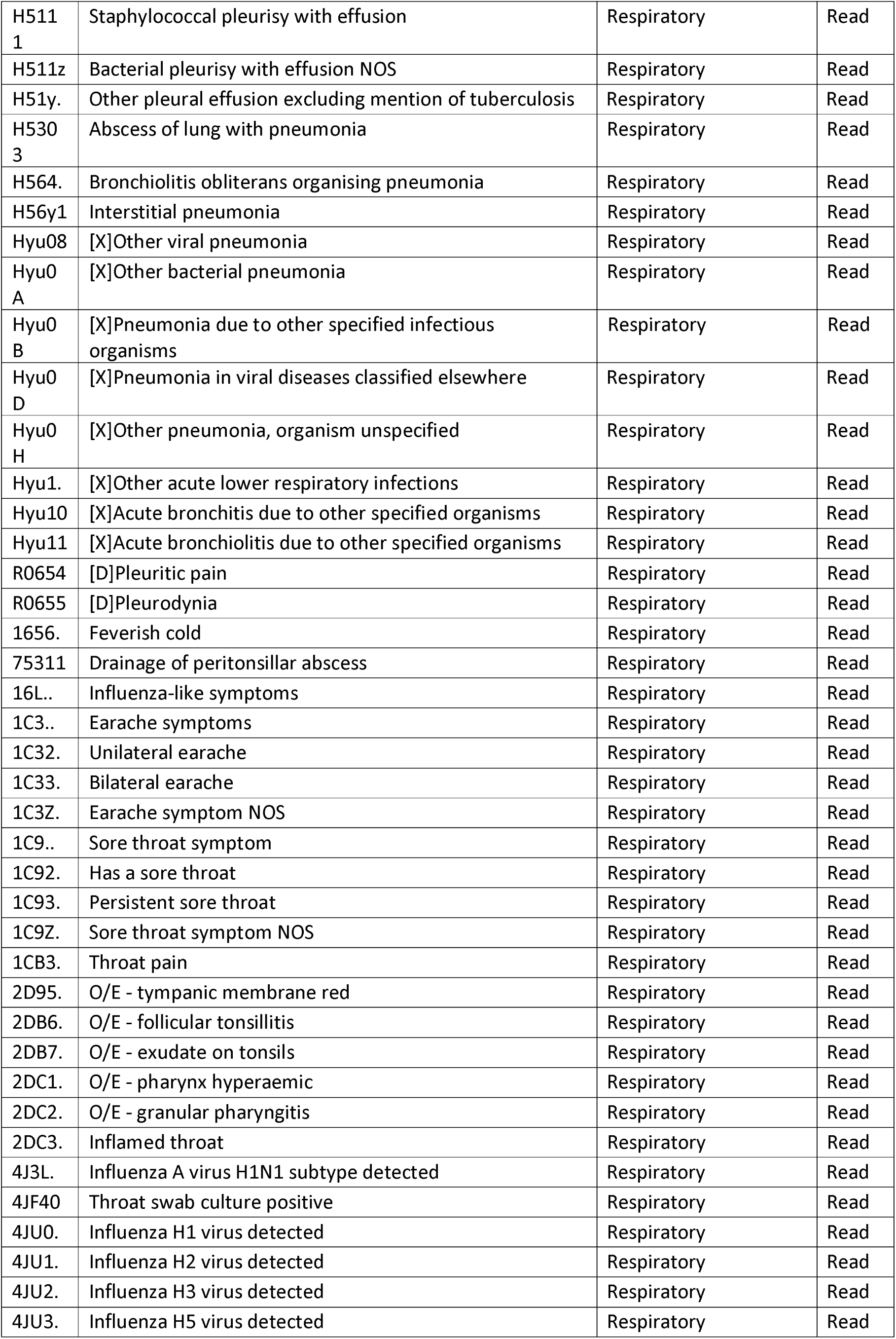
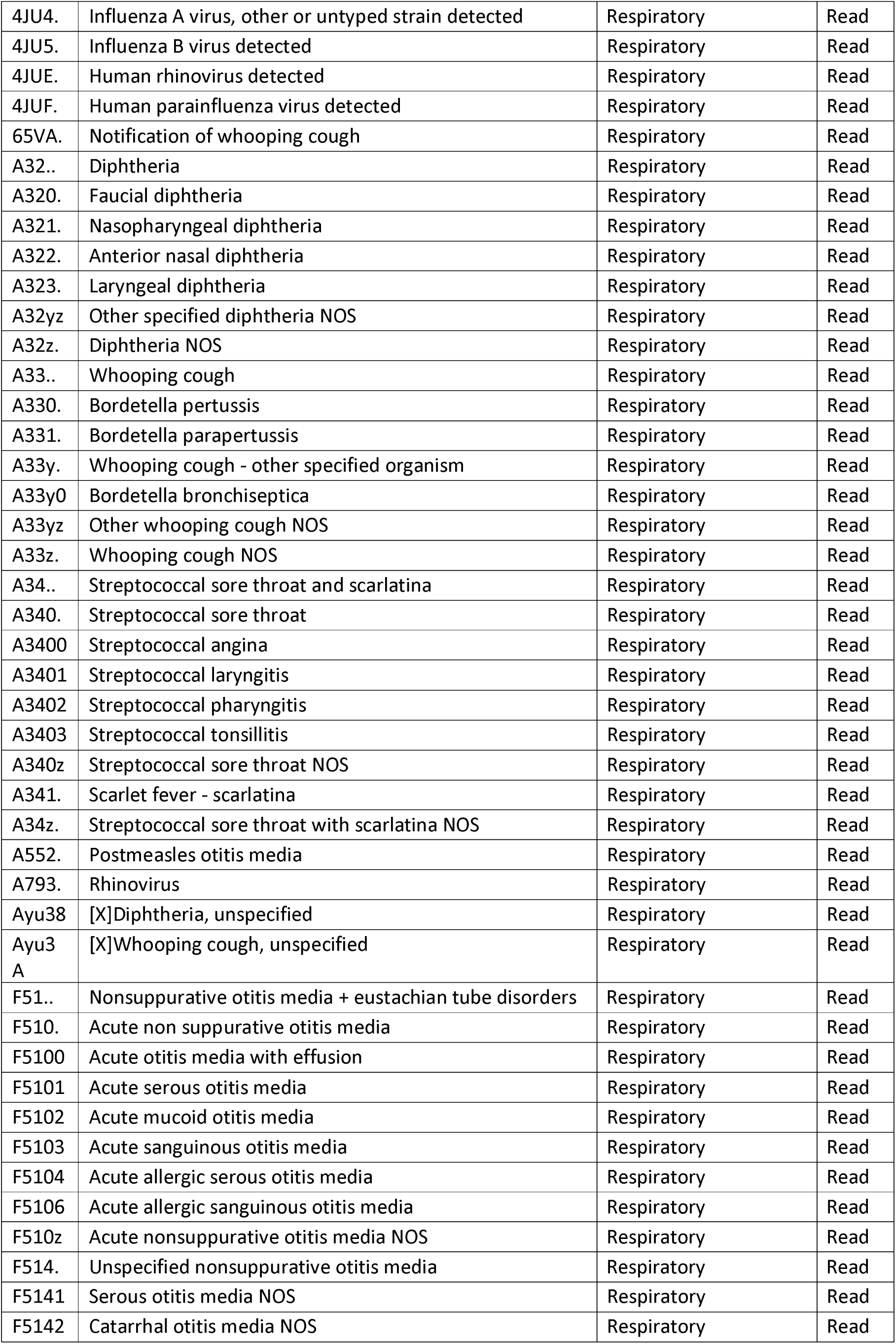
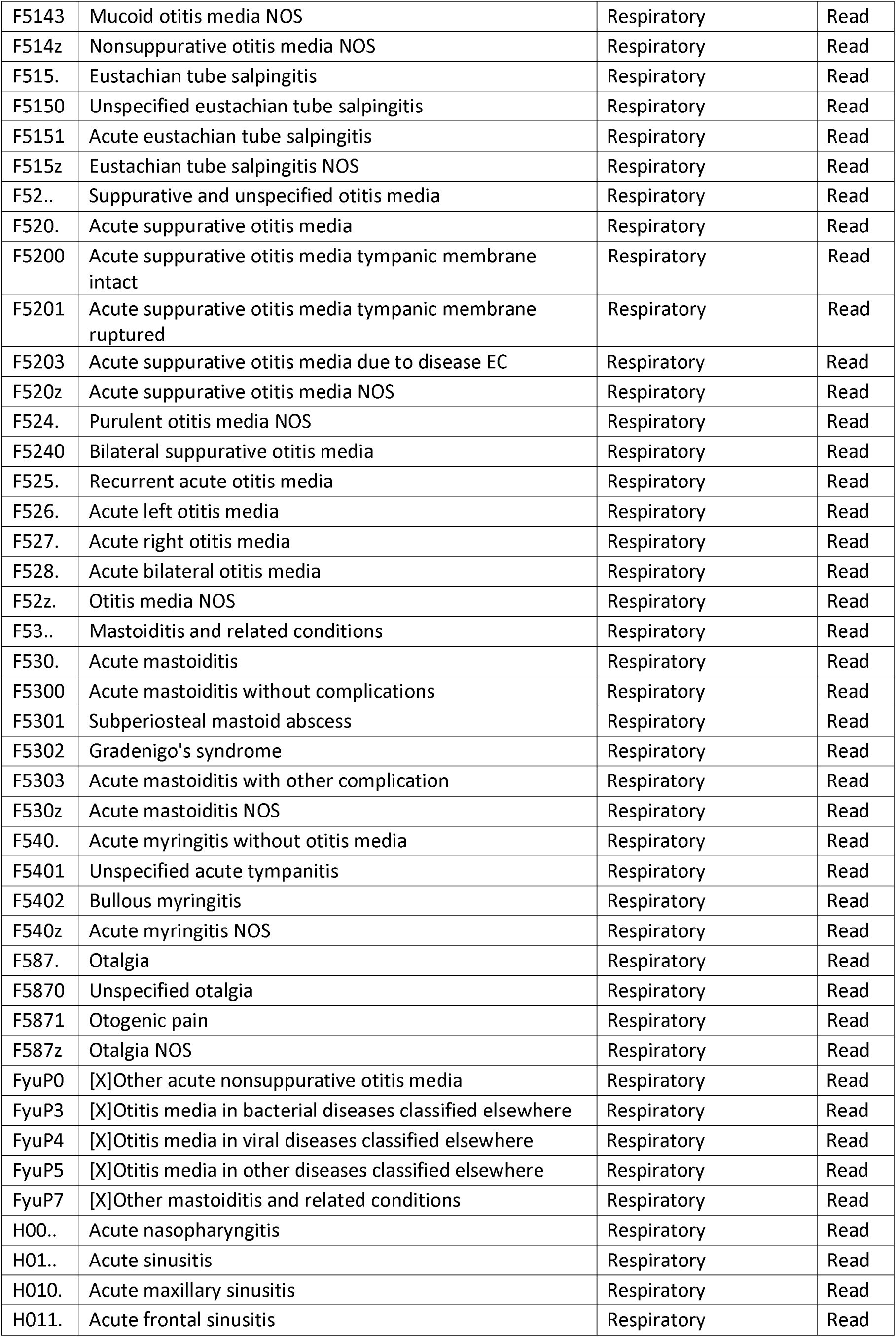
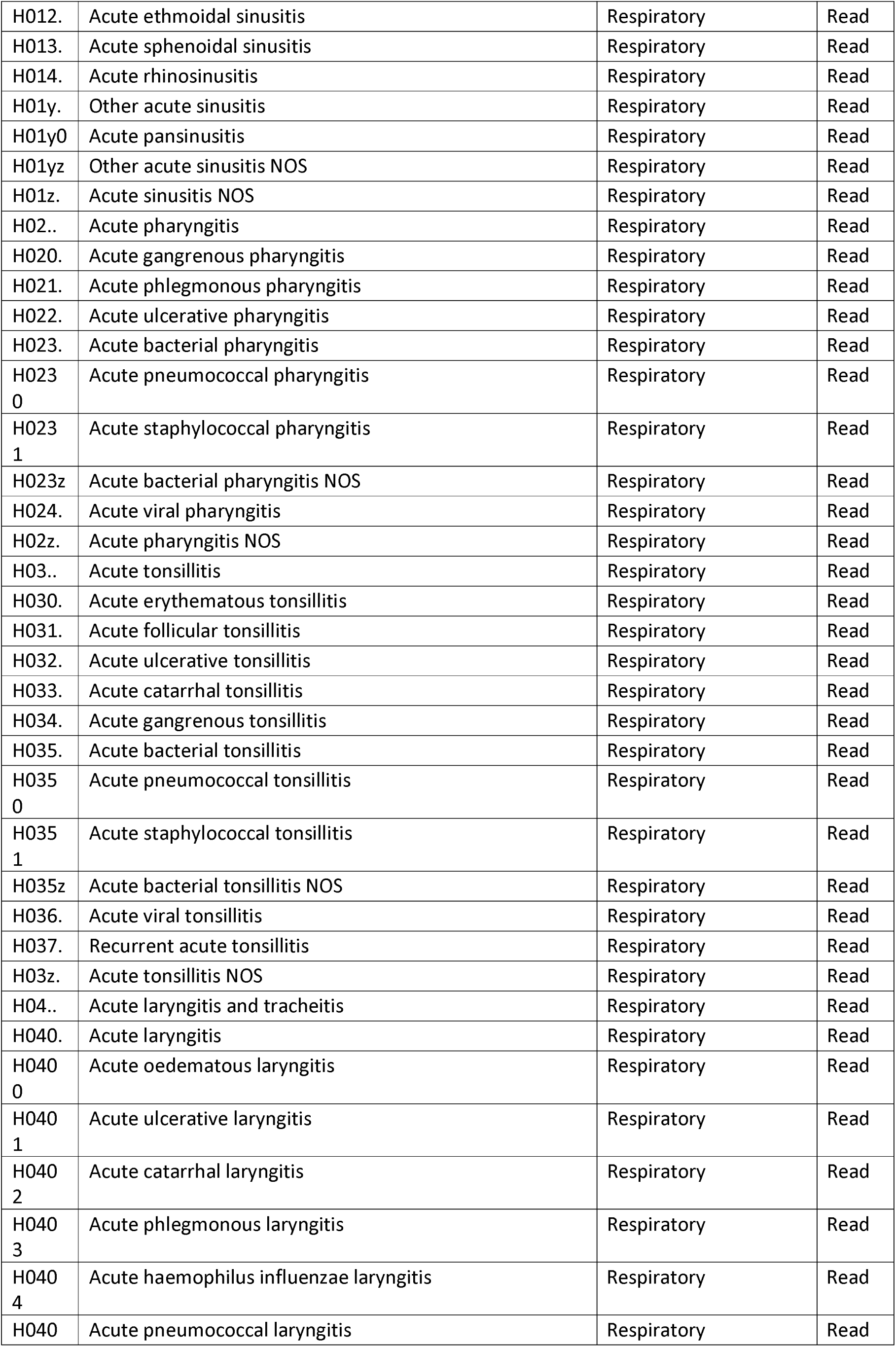
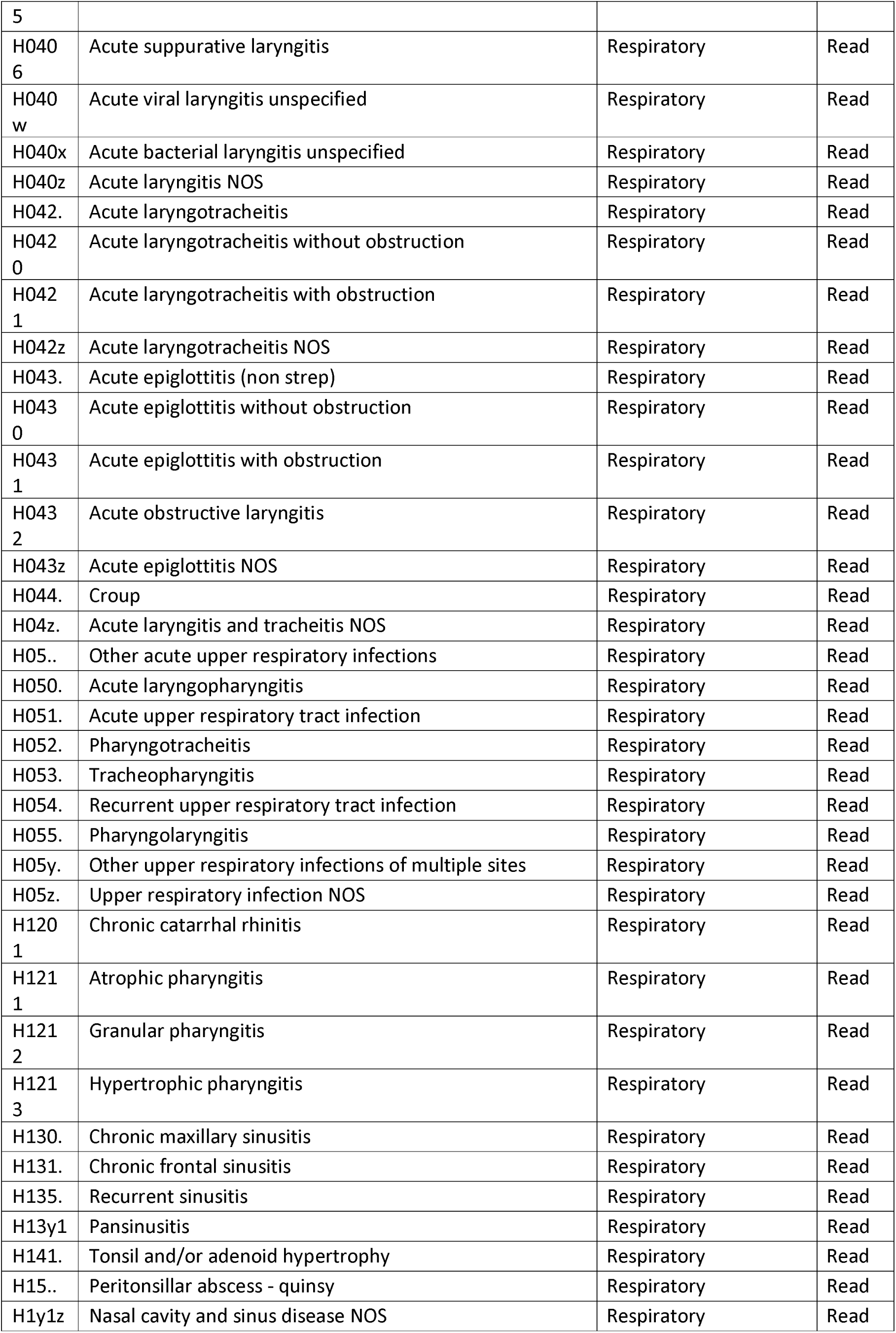
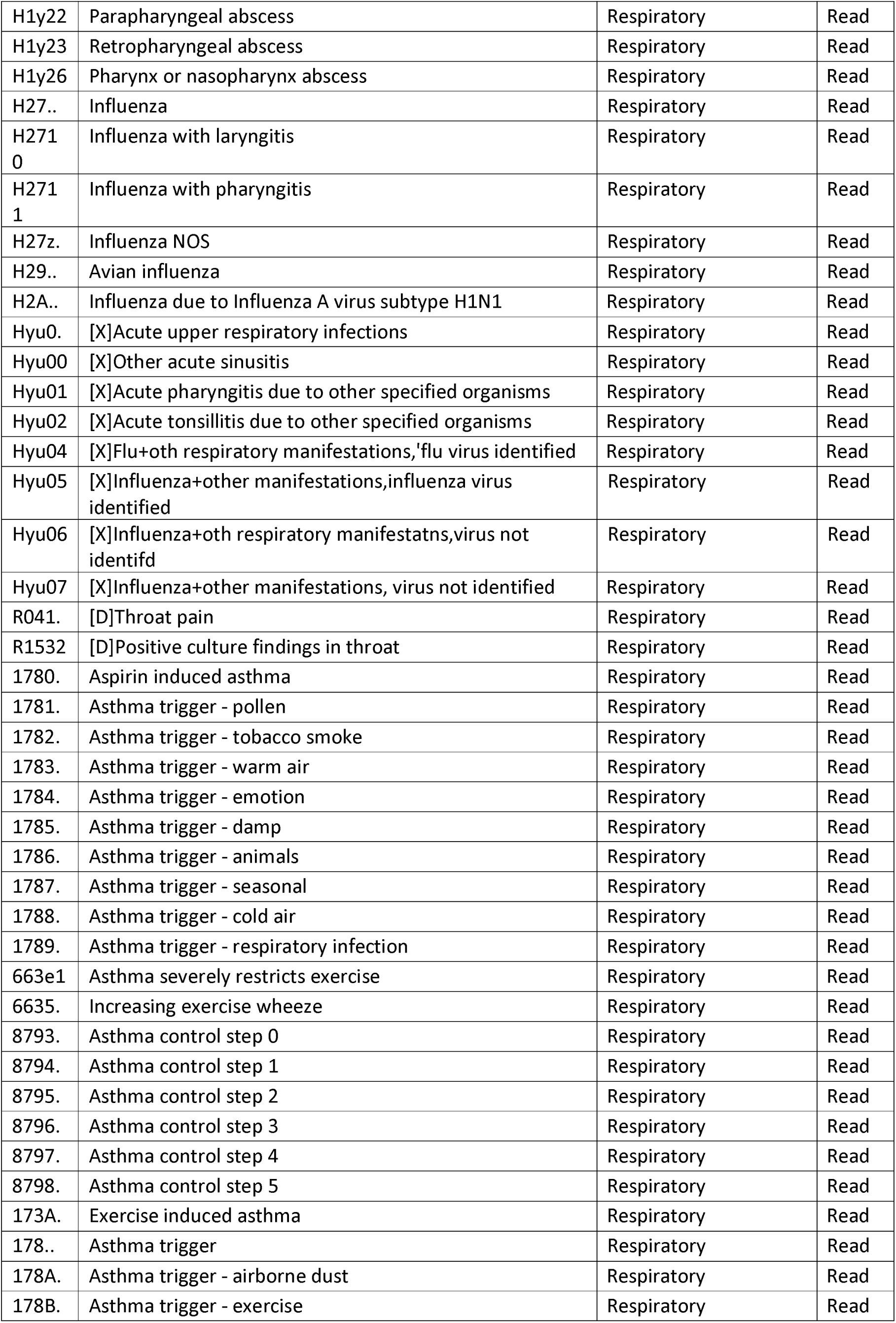
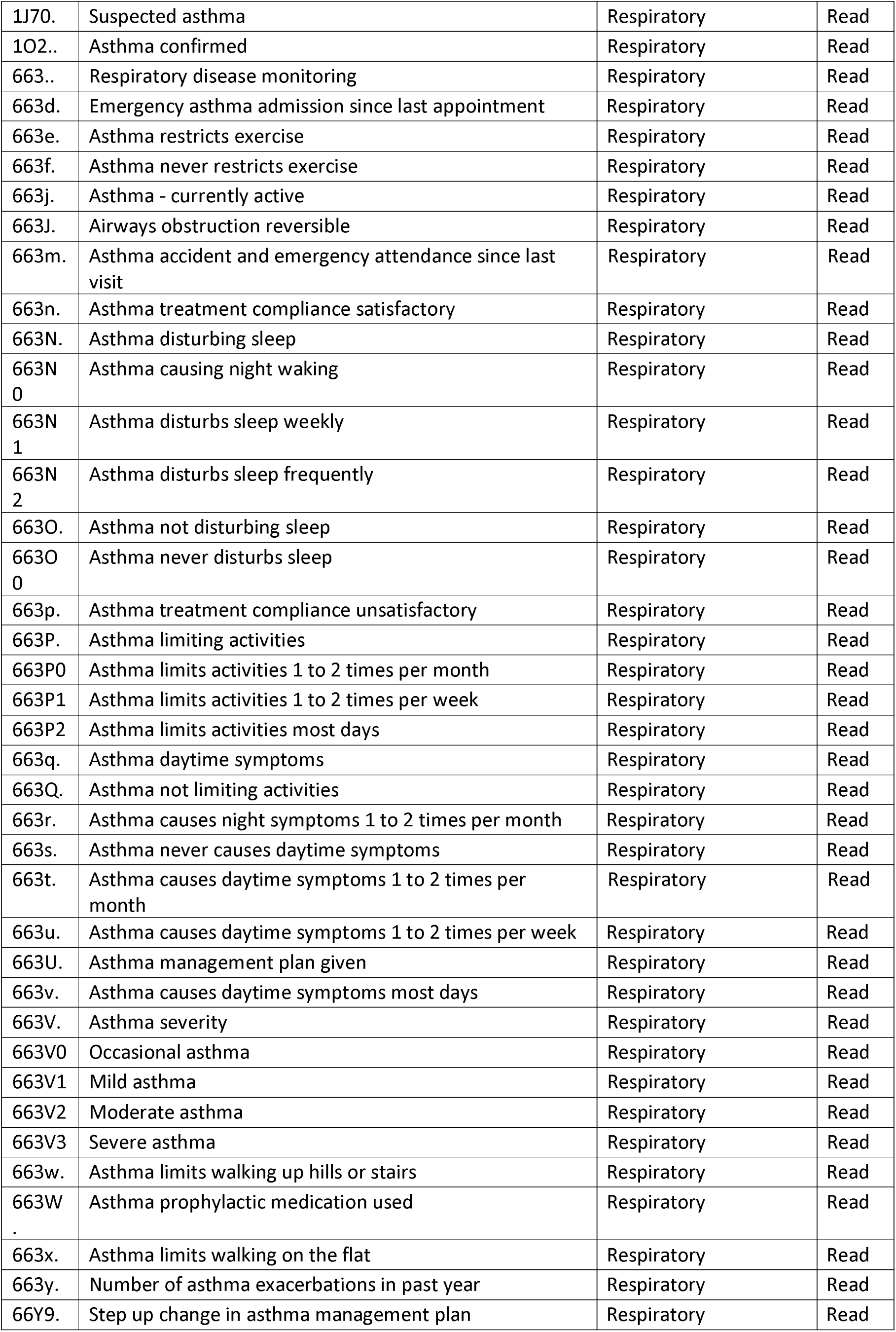
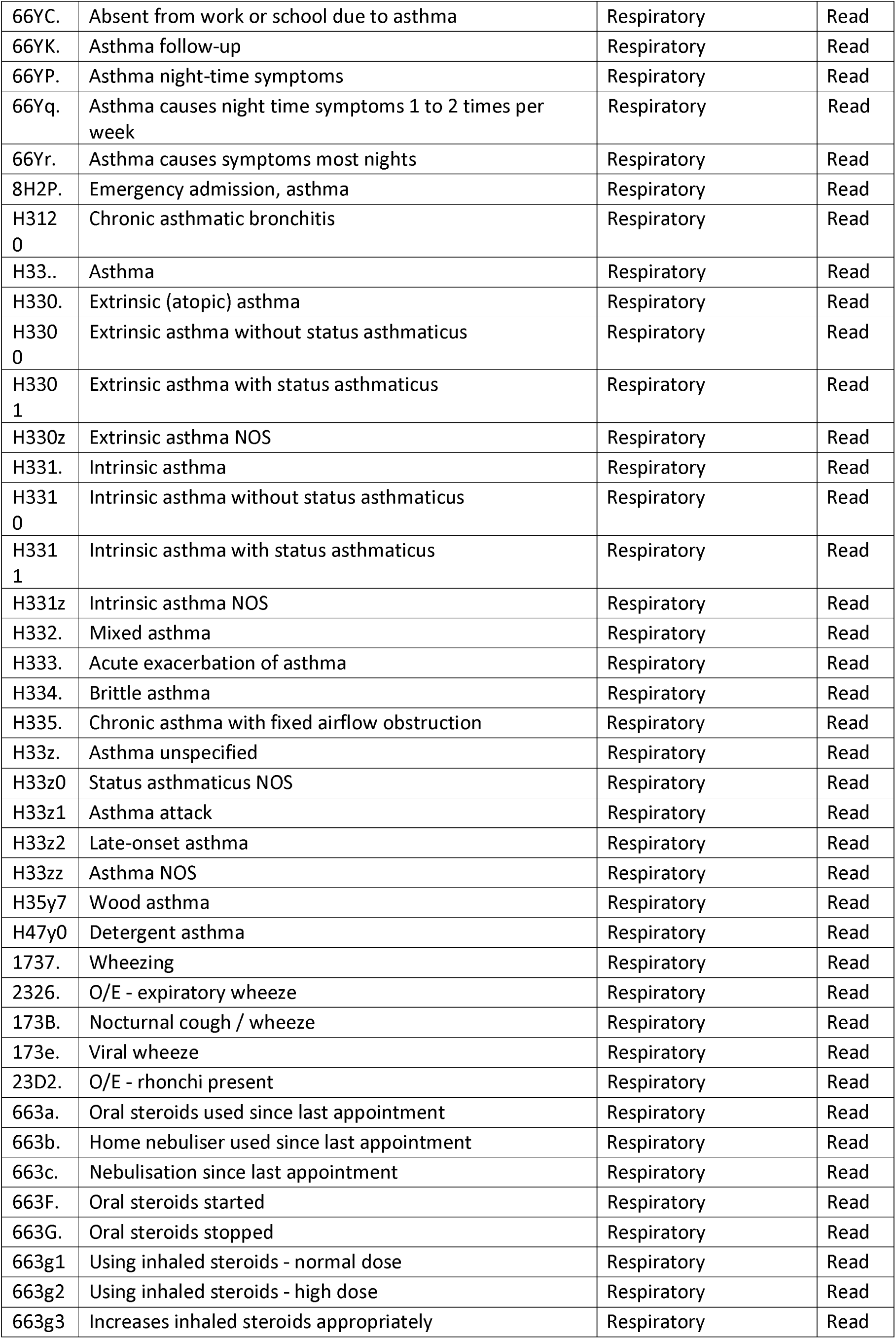
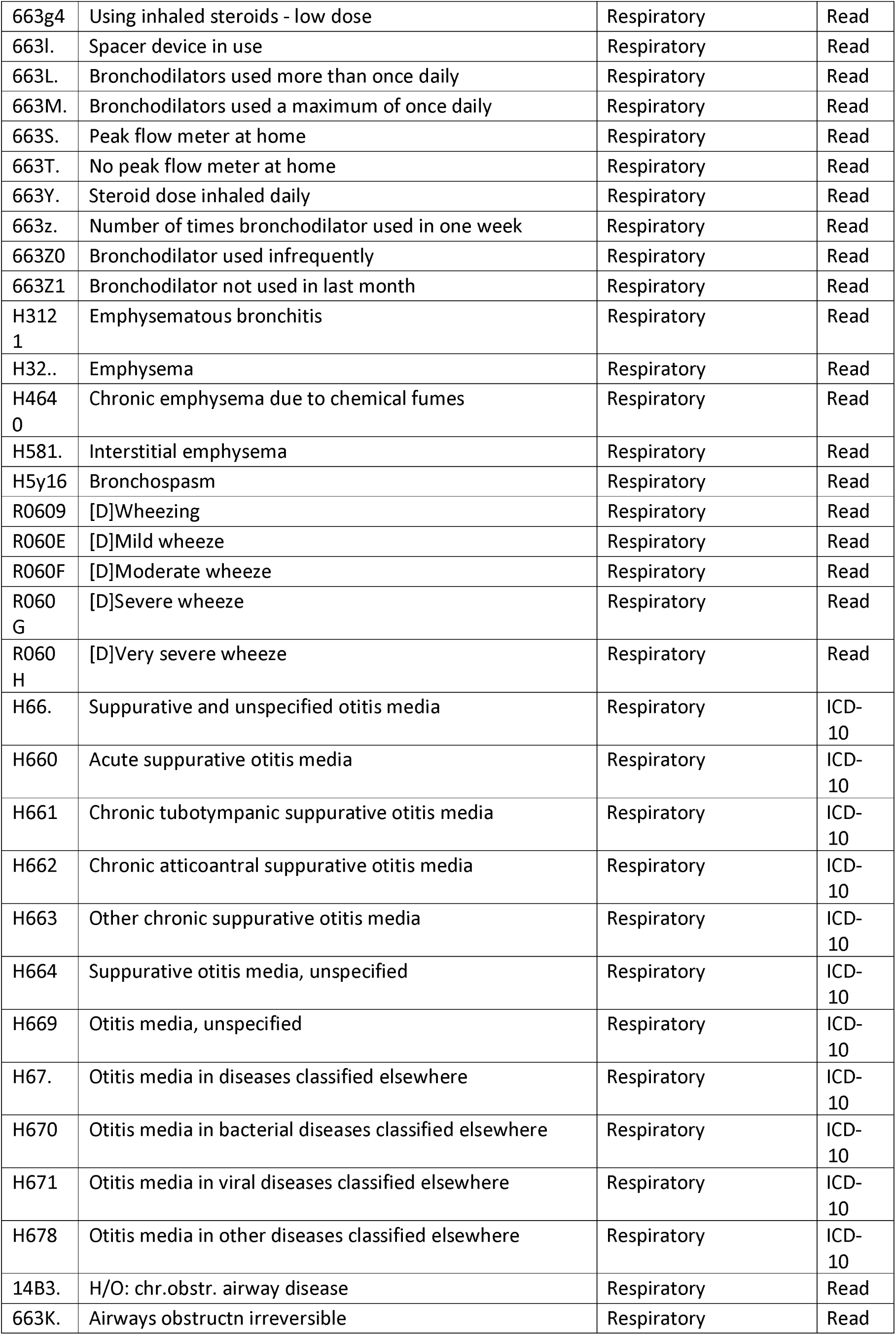
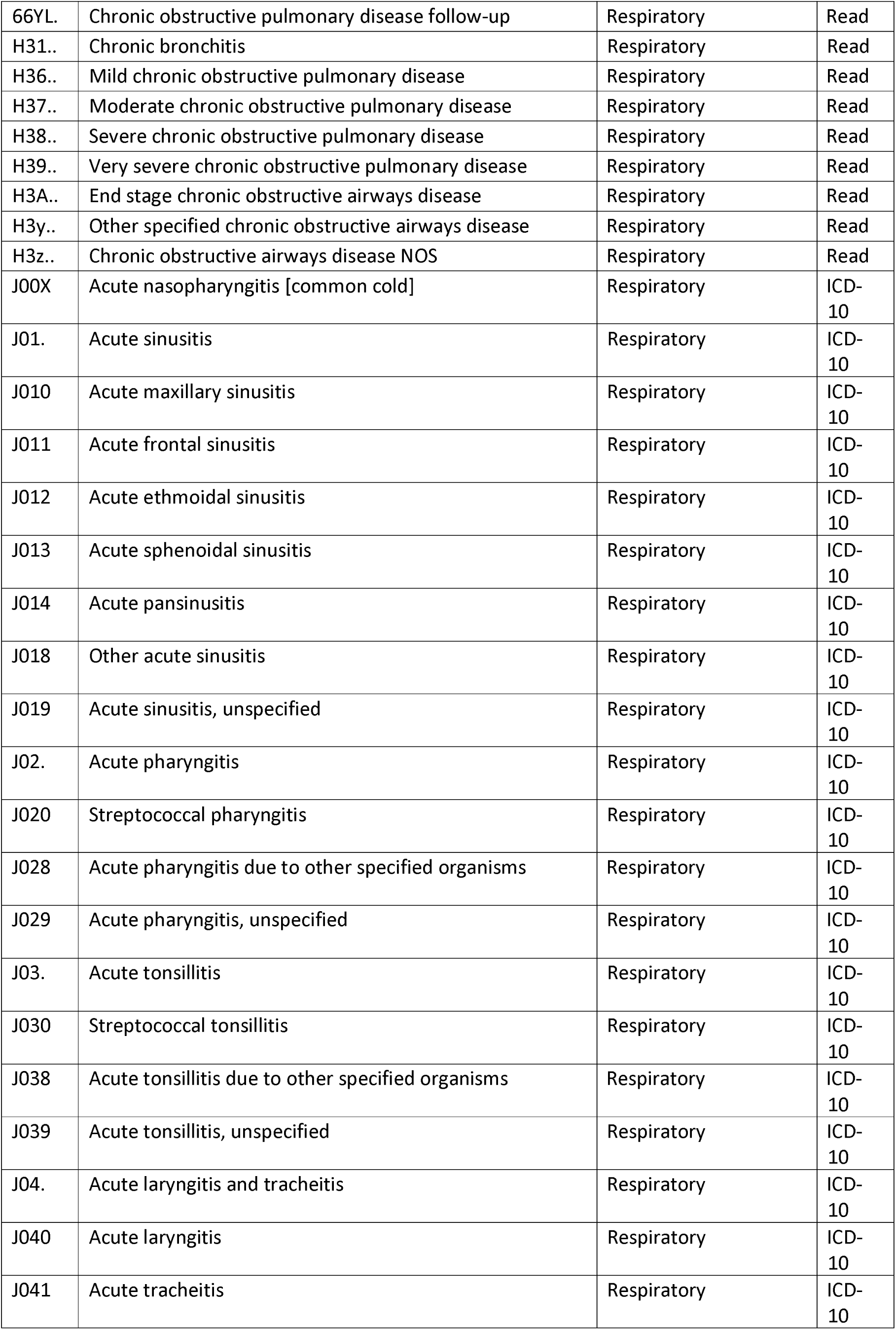
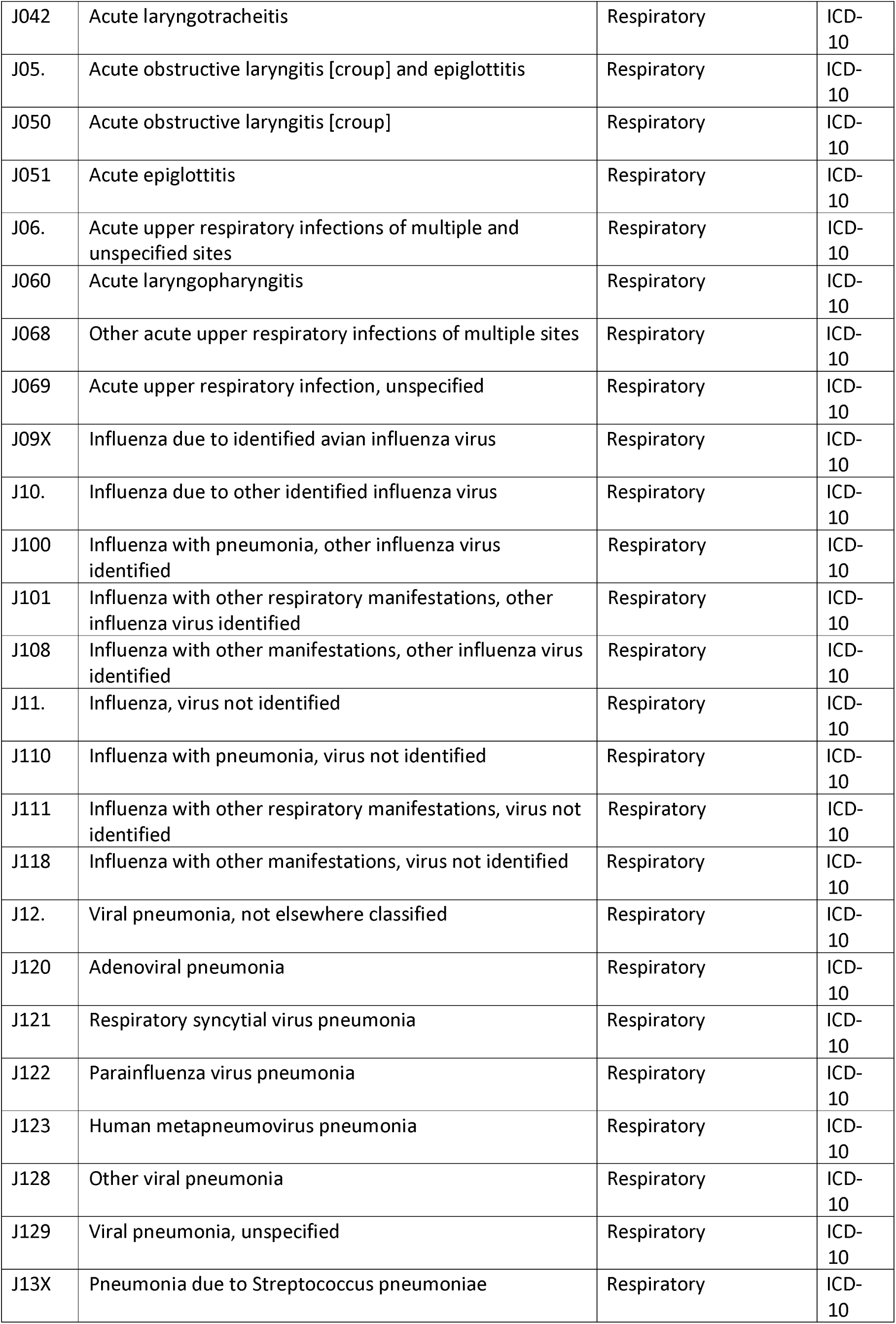
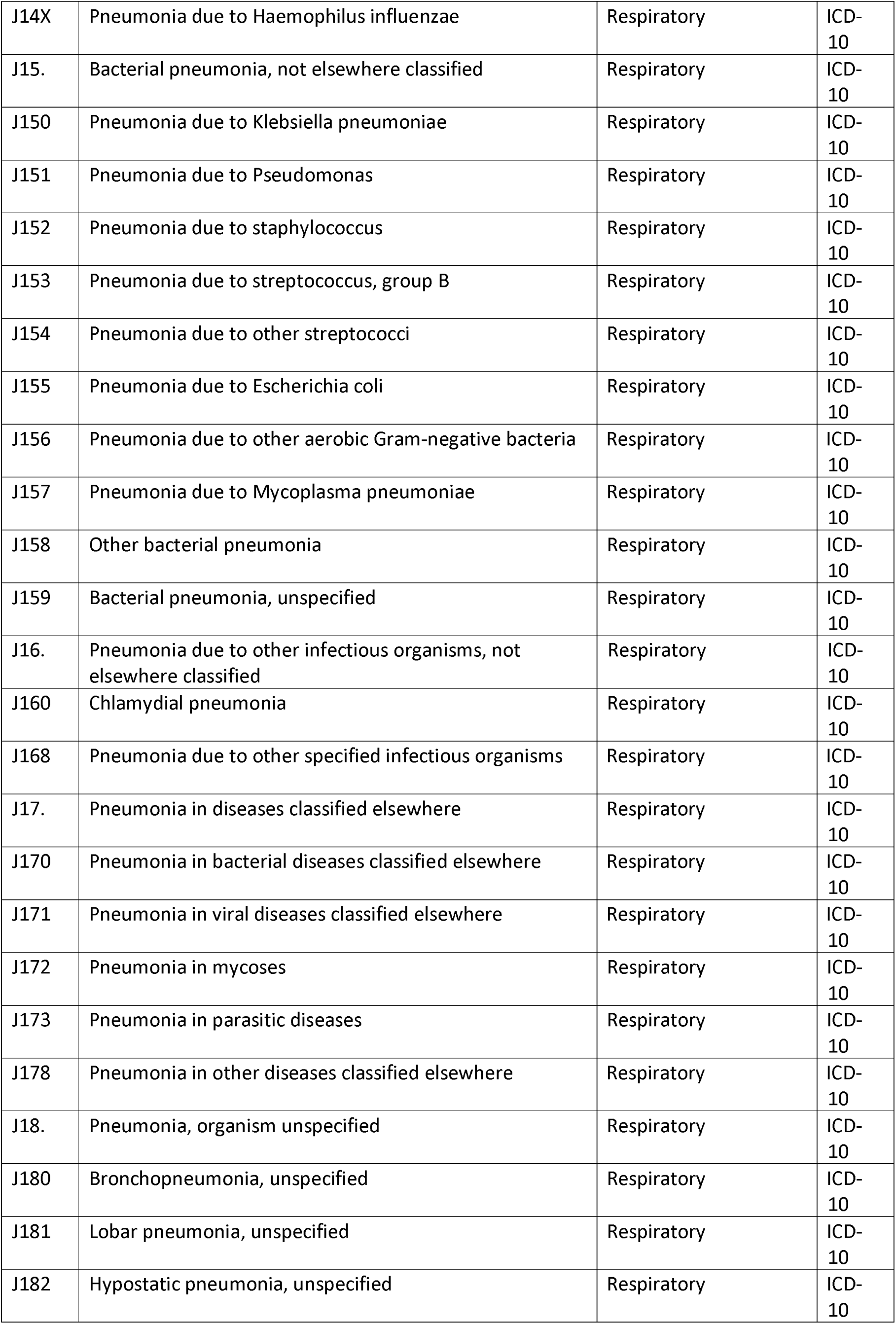
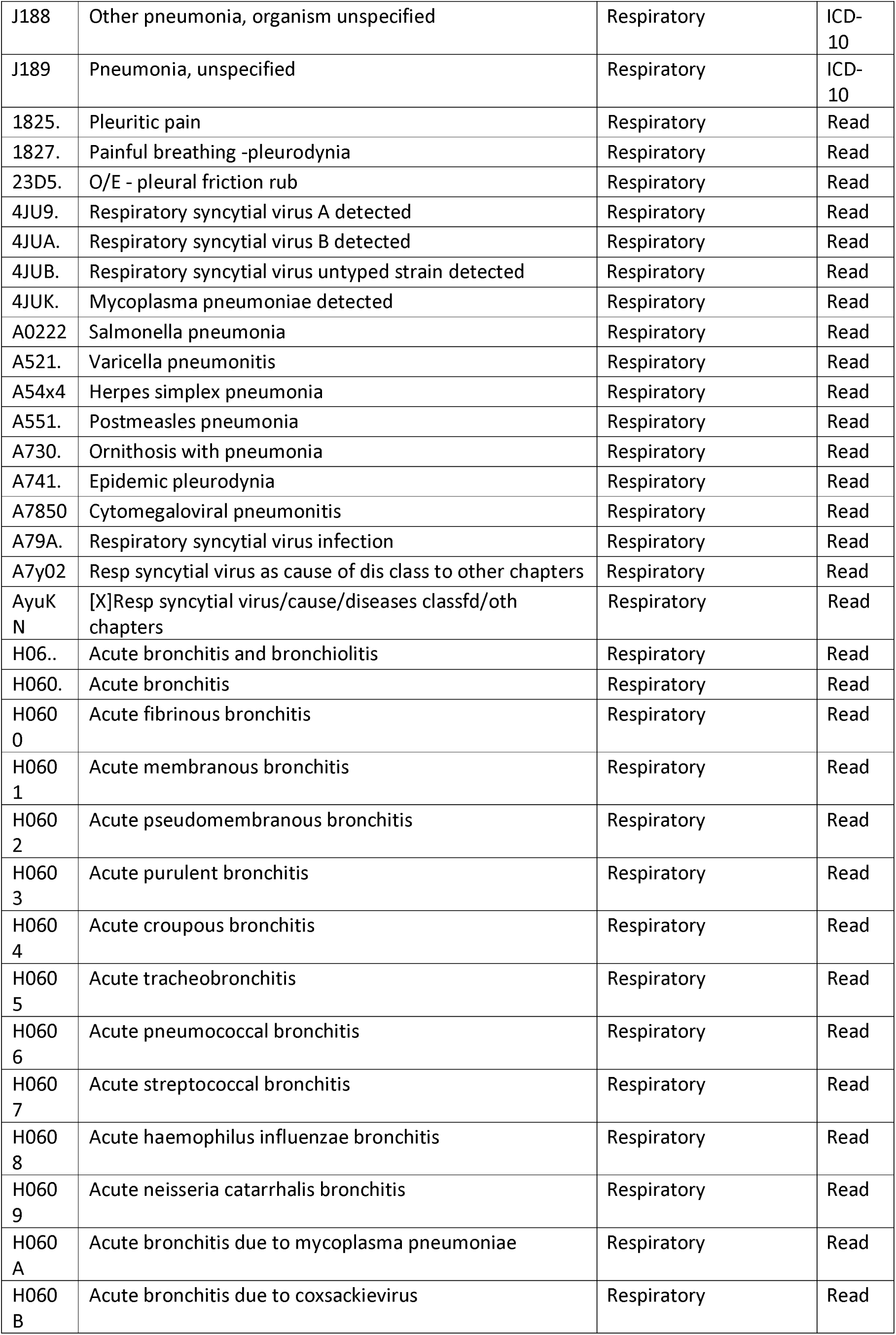
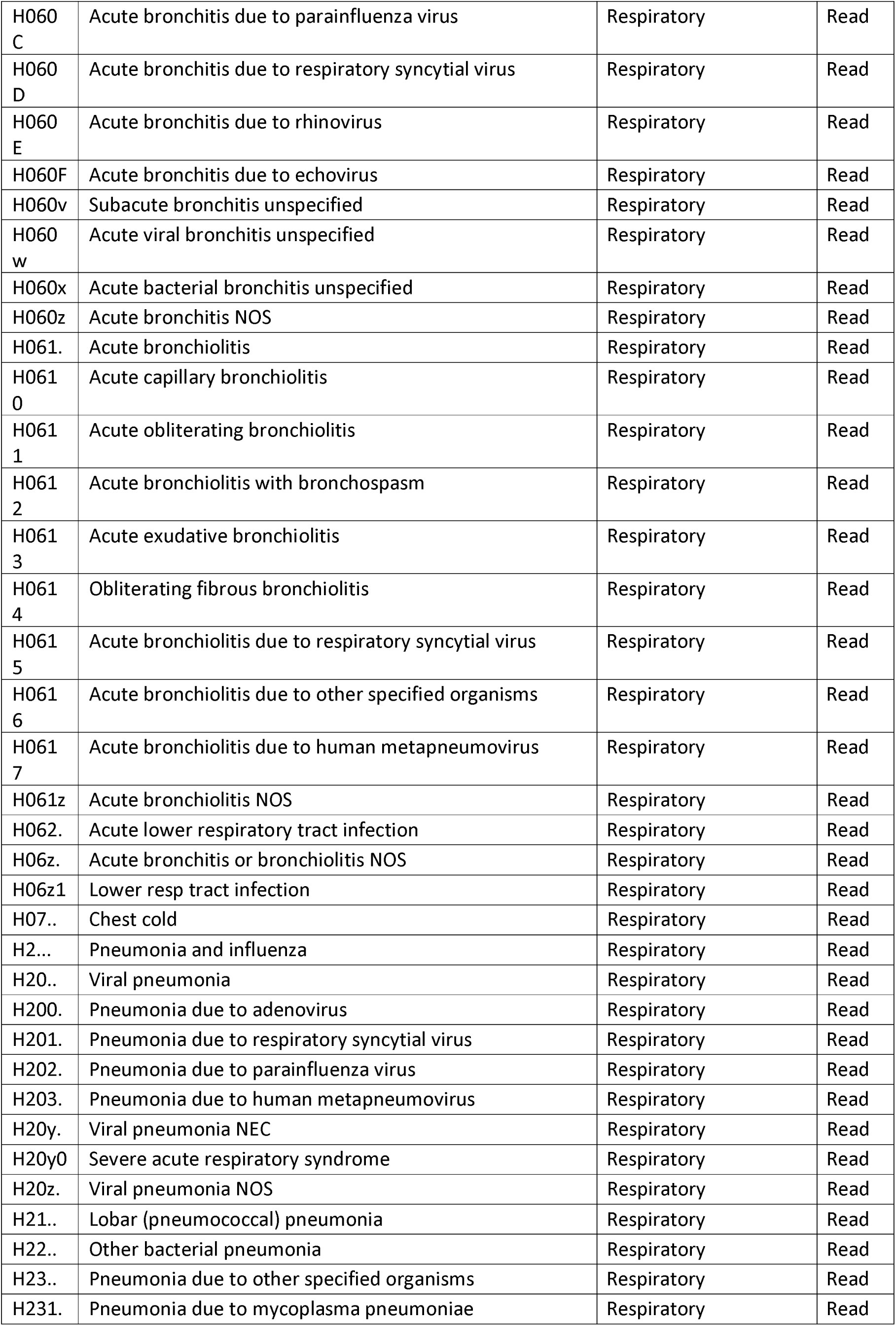
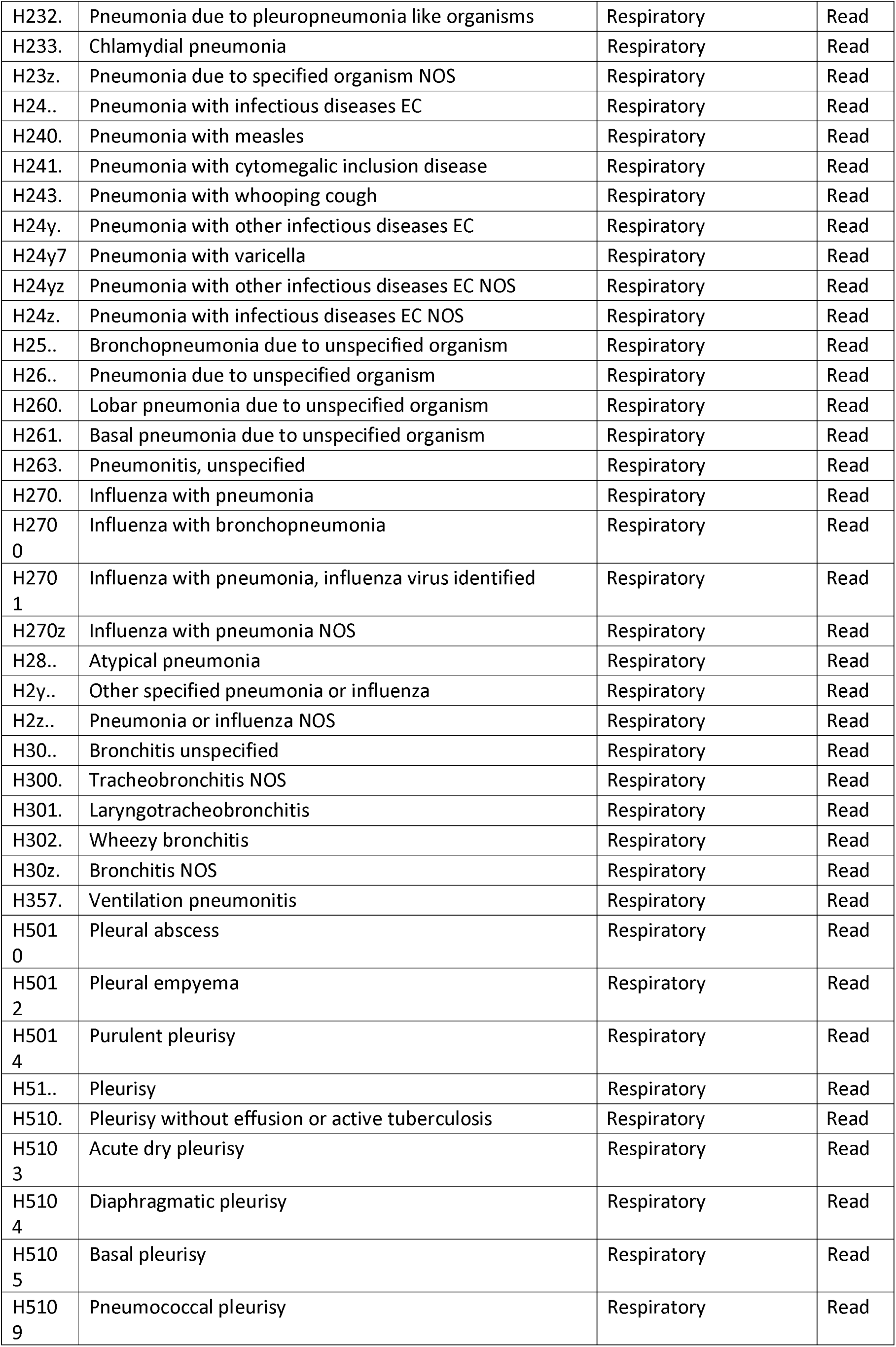
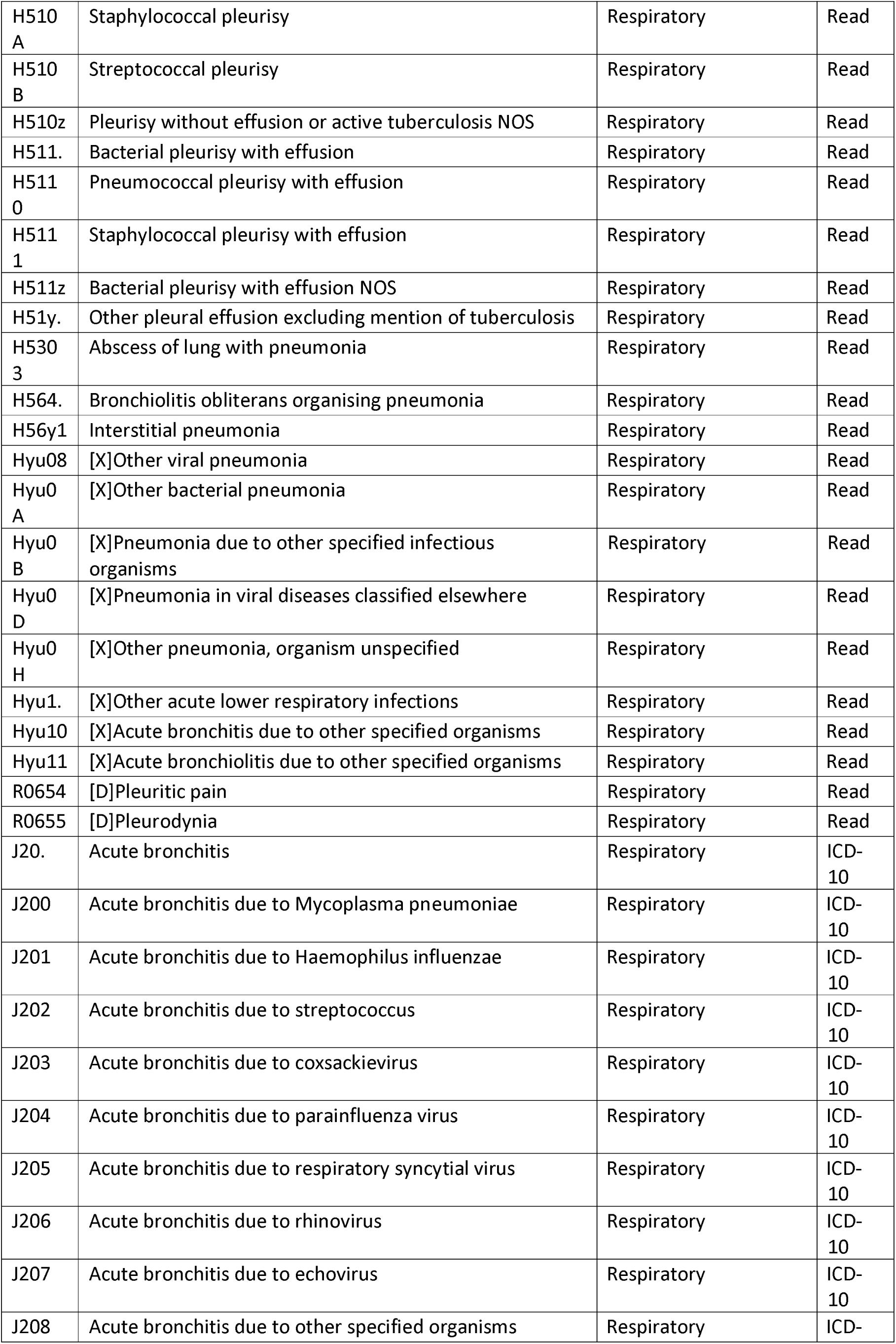
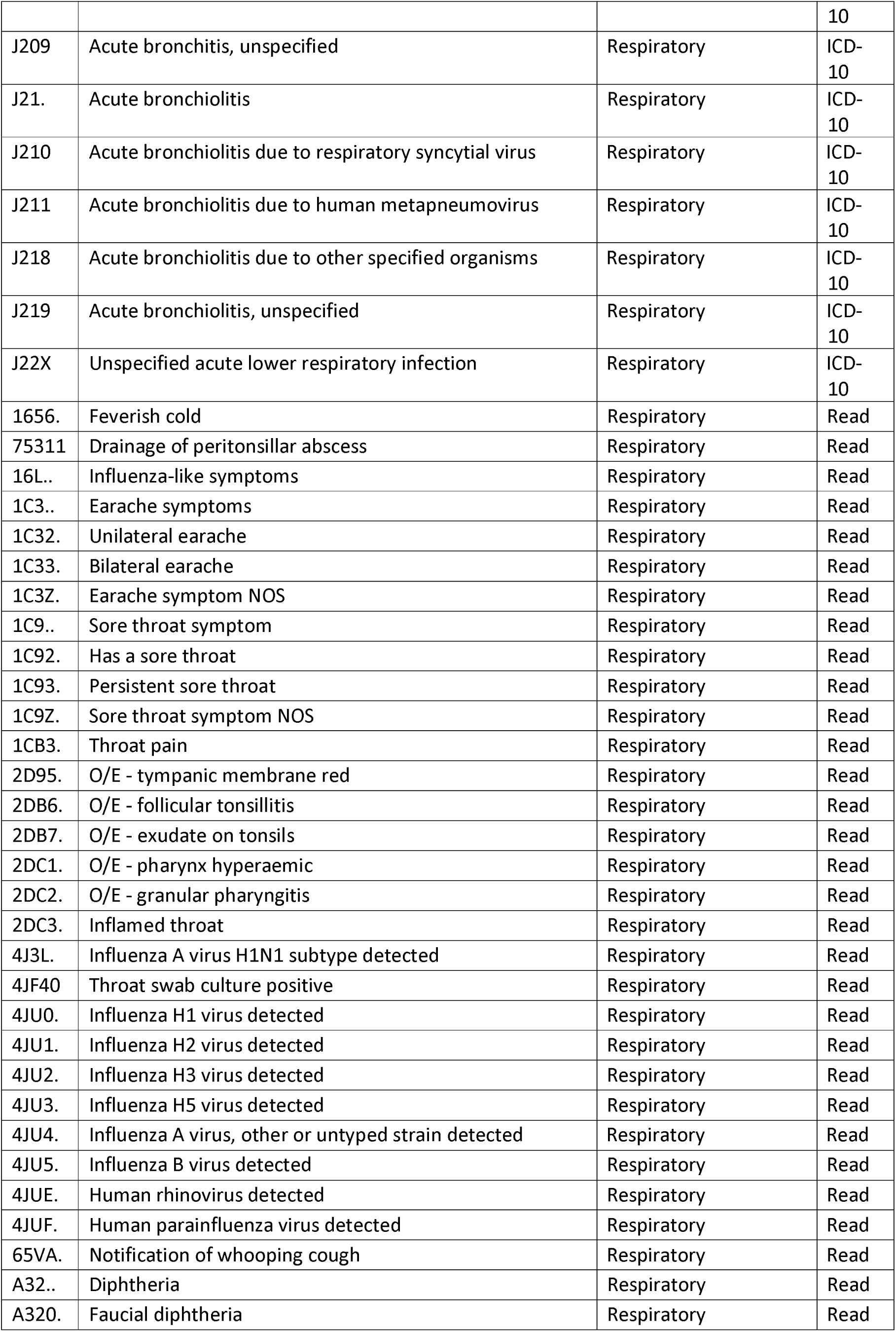
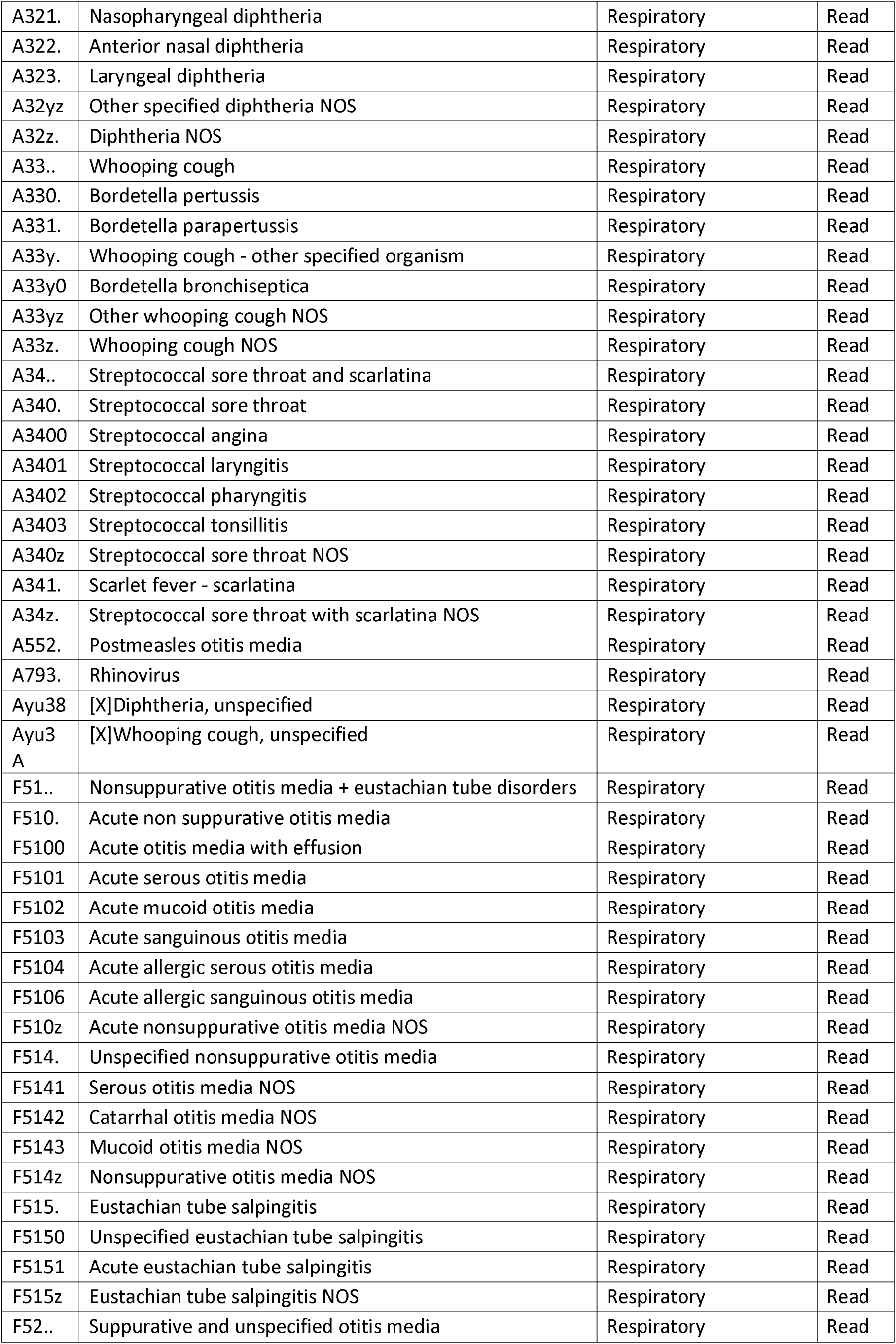
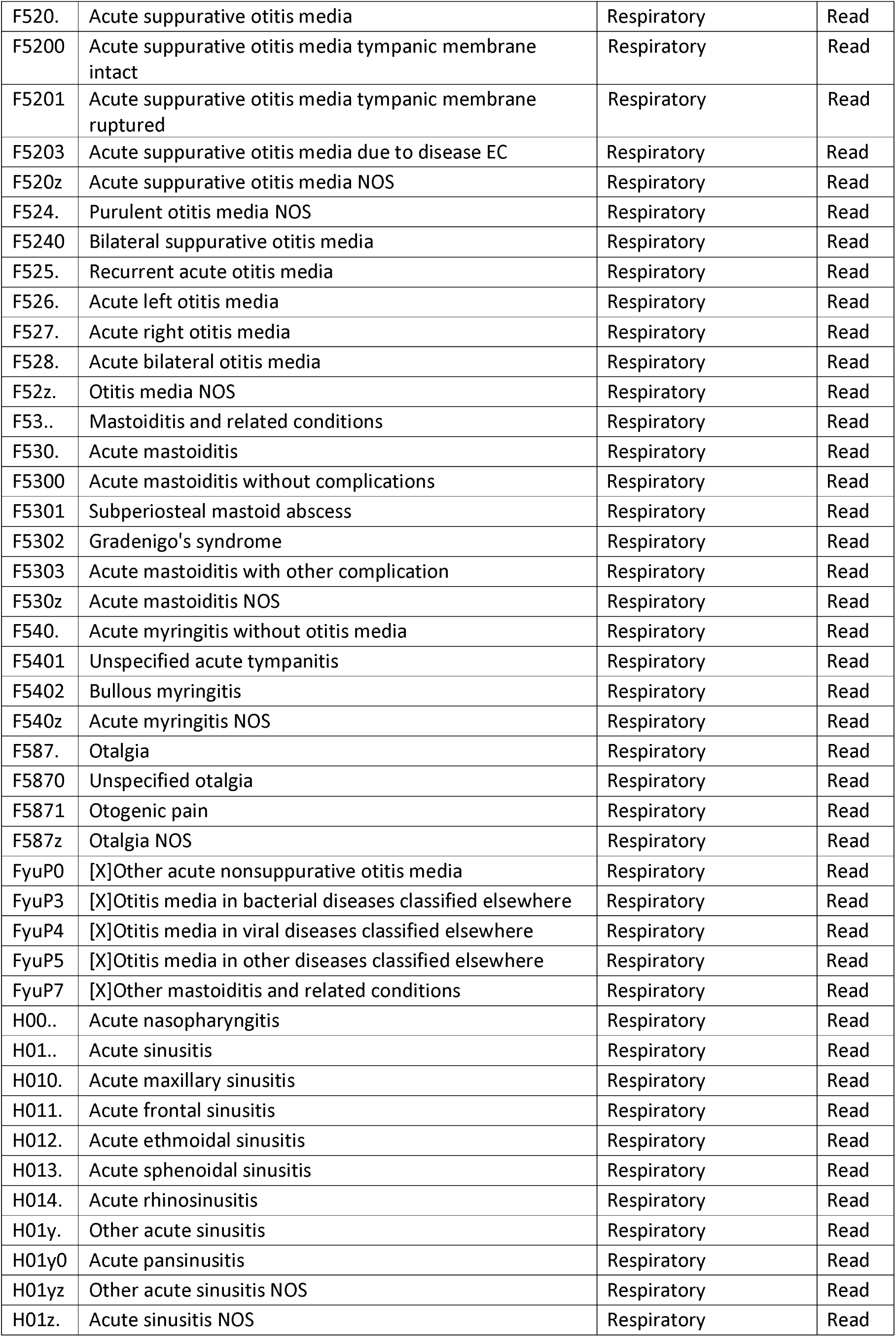
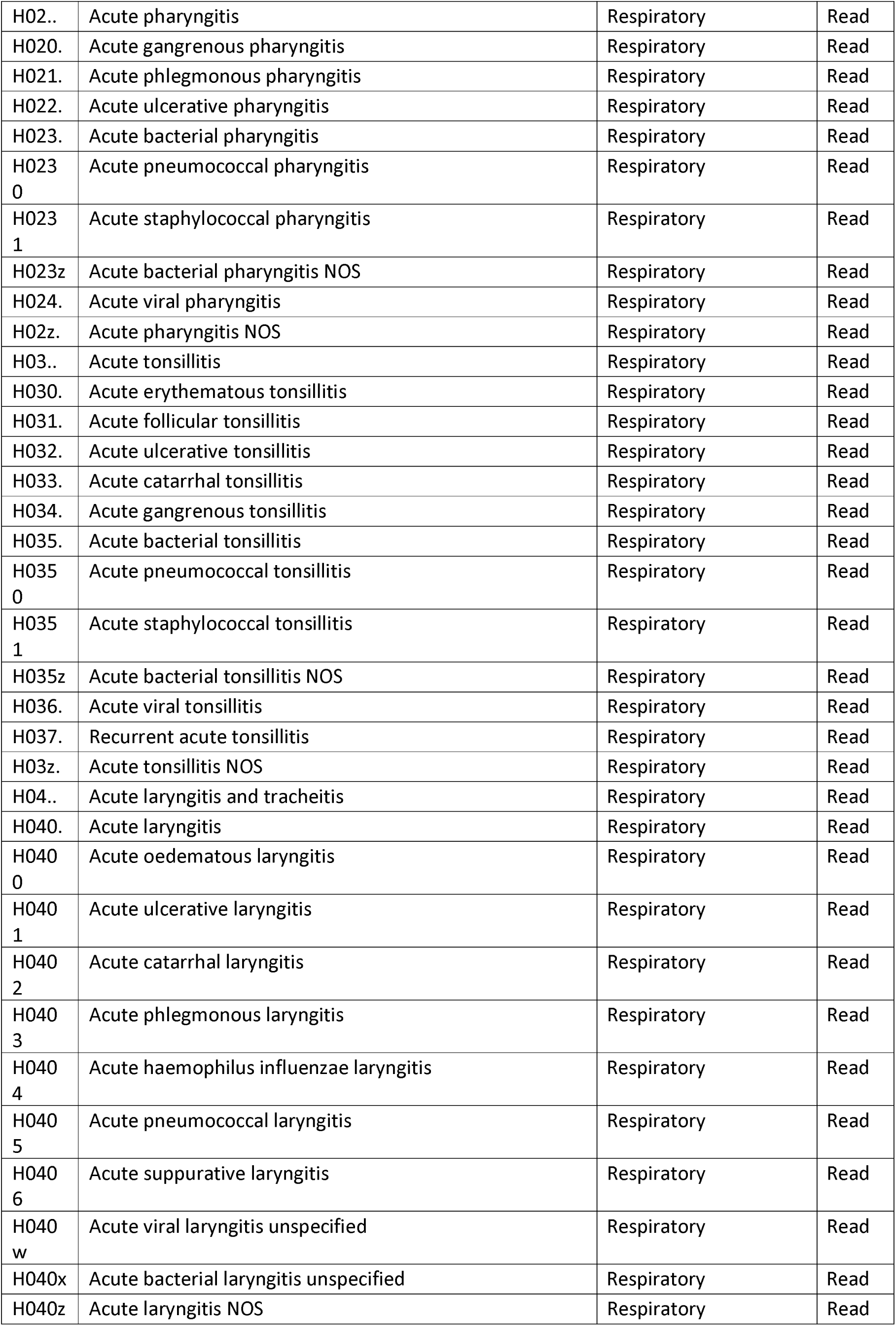
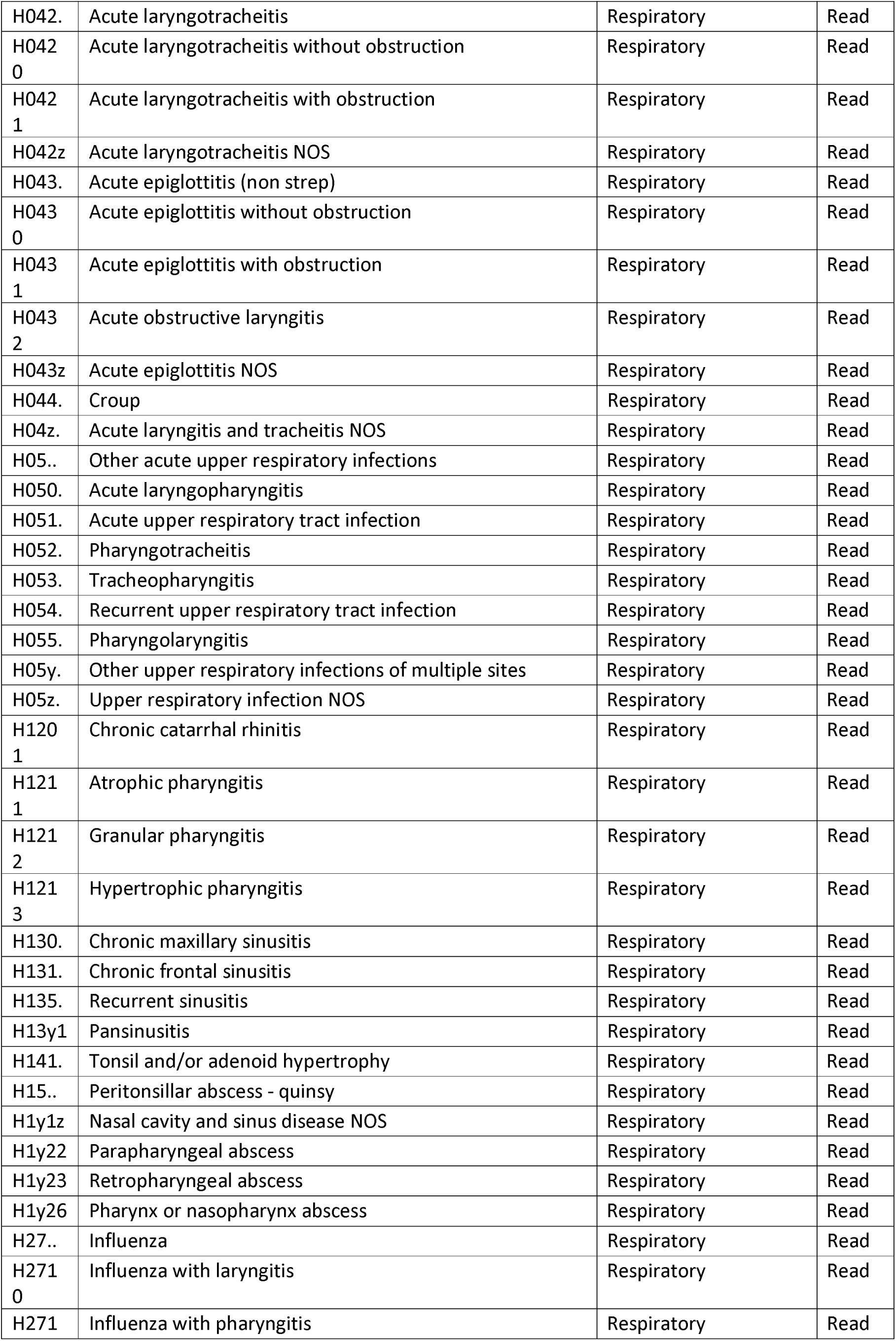
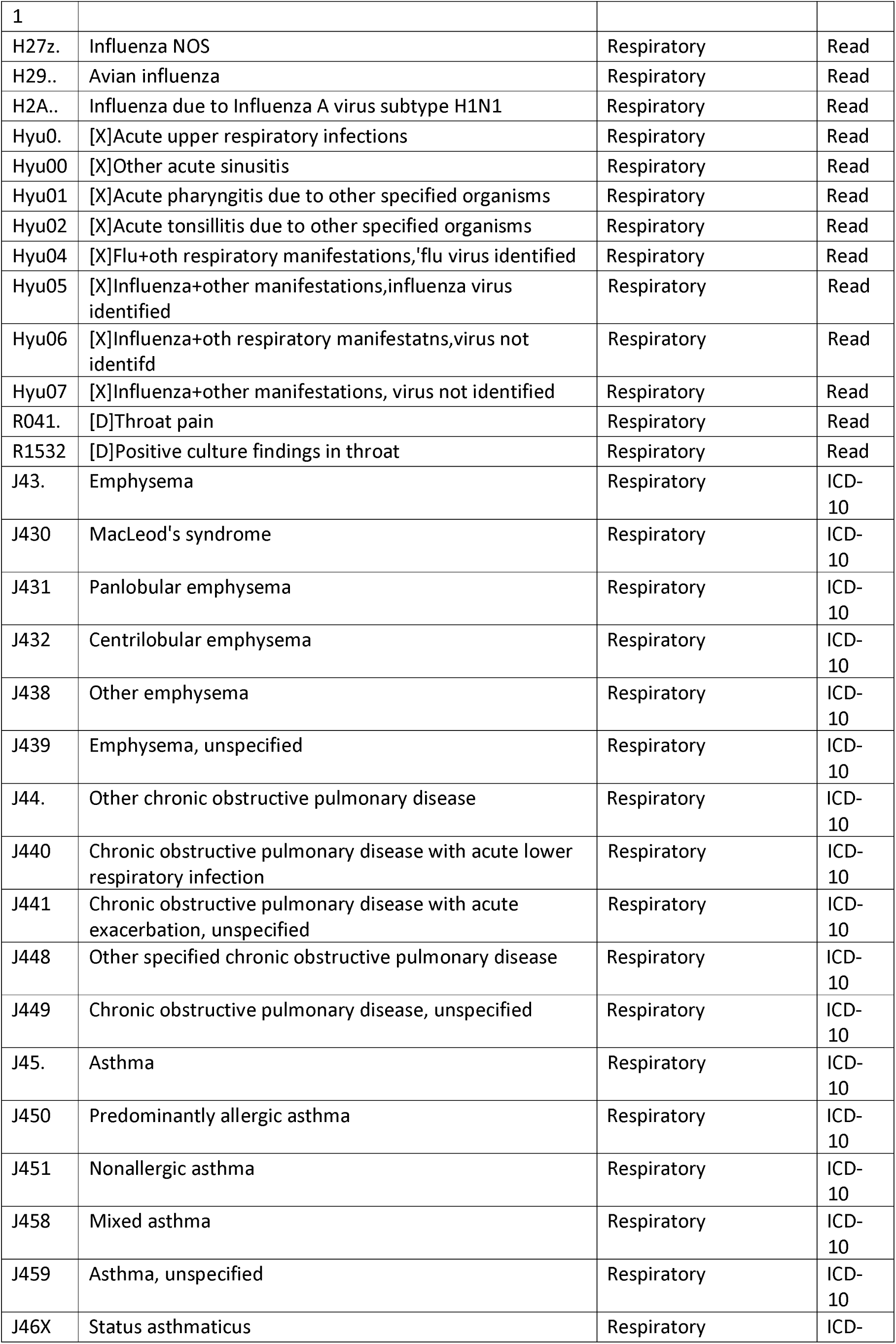
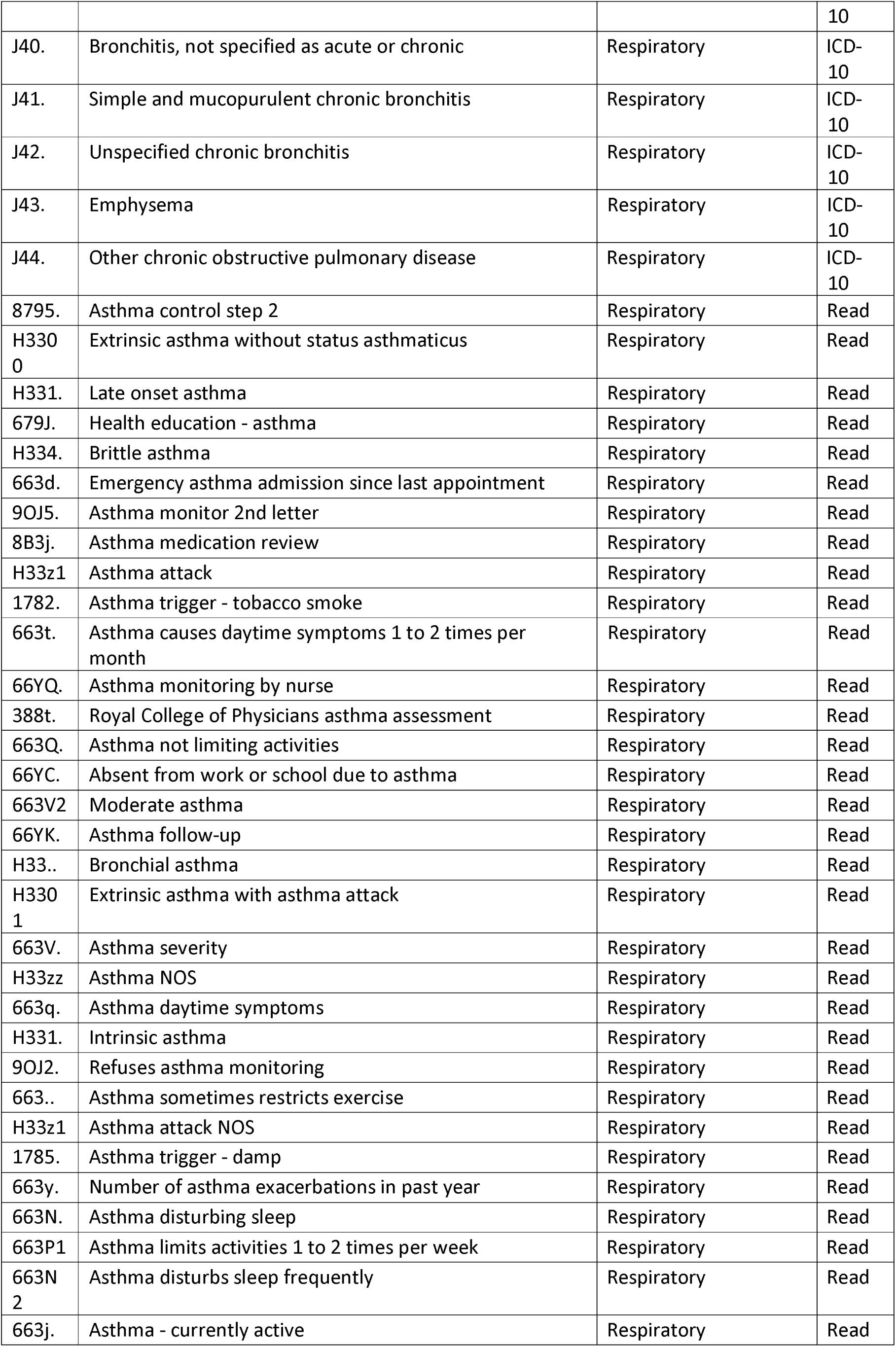
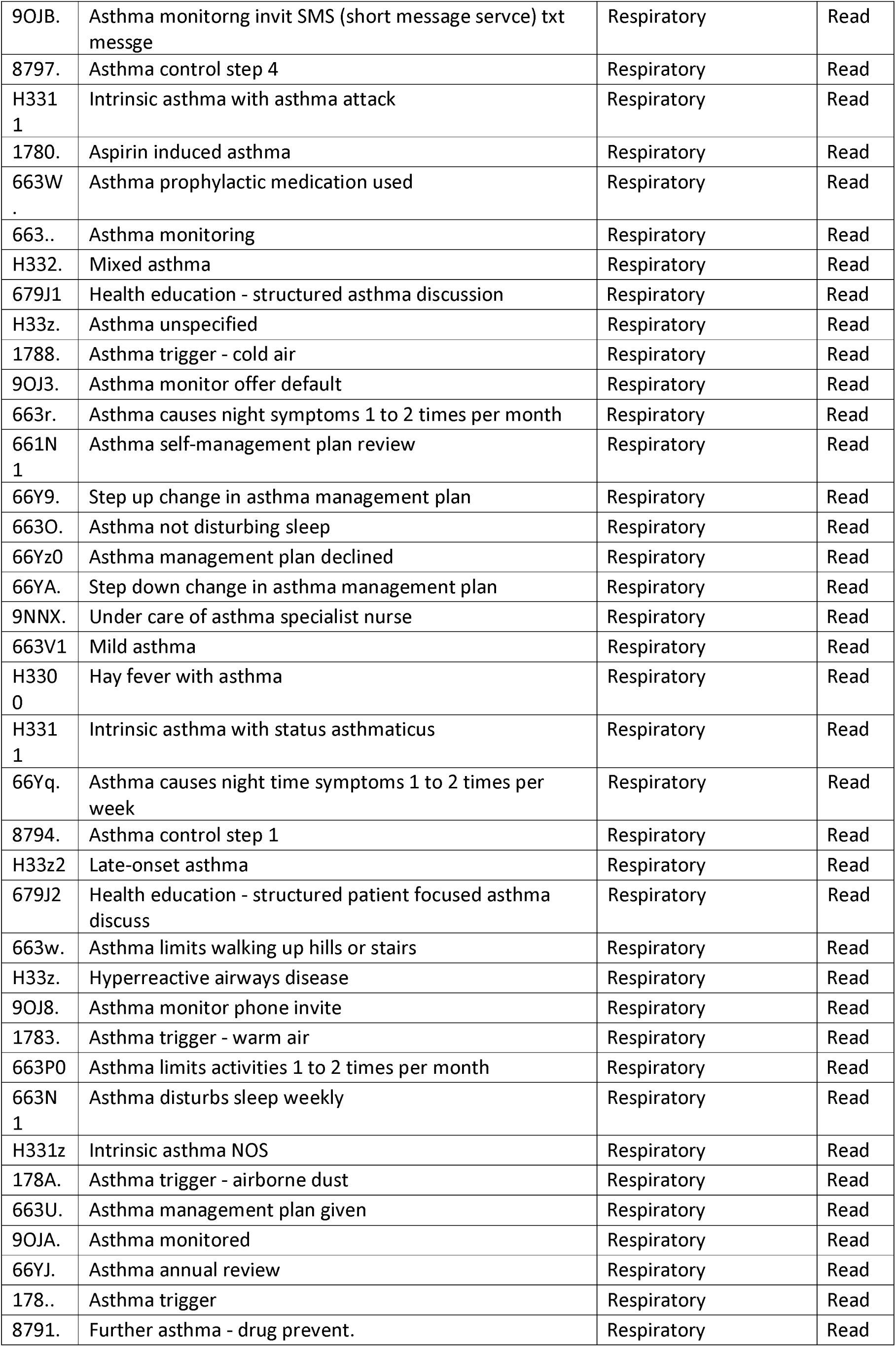
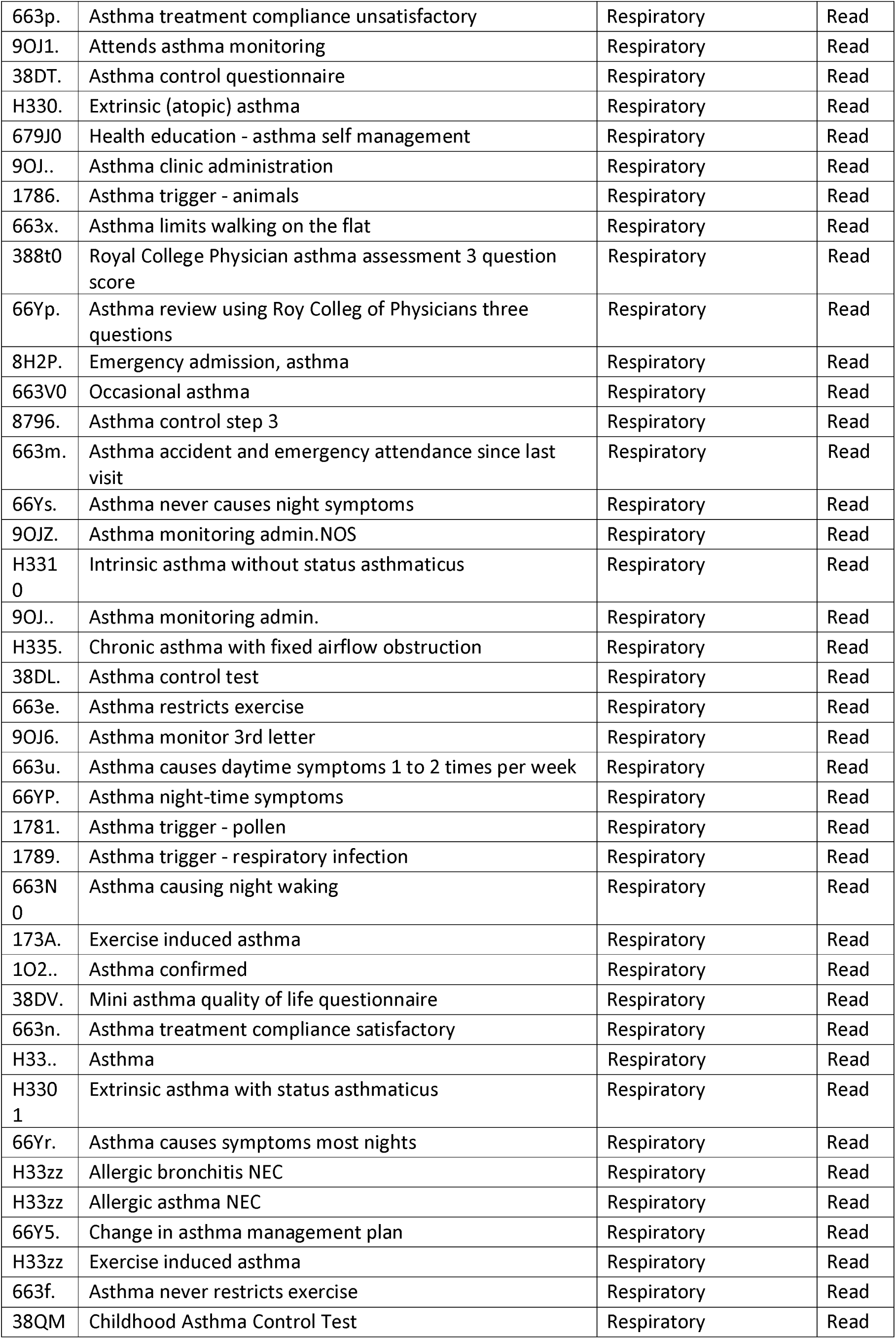
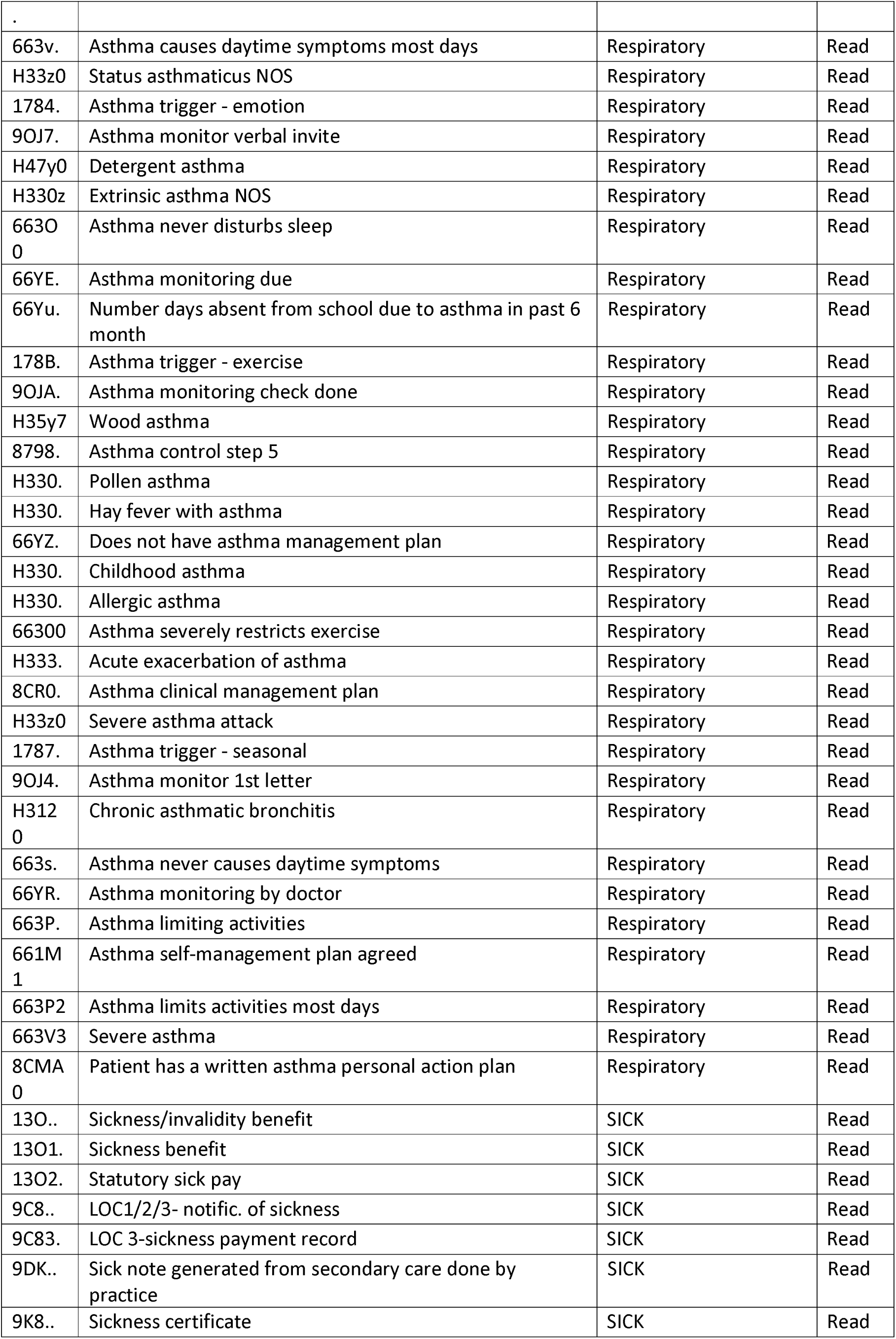
Table of Codes under investigation.

## References

1. Drew DA, Nguyen LH, Steves CJ, Menni C, Freydin M, Varsavsky T, et al. Rapid implementation of mobile technology for real-time epidemiology of COVID-19. Science [Internet]. 2020 Jun 19 [cited 2022 Mar 3];368(6497):1362–7. Available from: https://www.science.org/doi/abs/10.1126/science.abc0473

2. Kingstone T, Taylor AK, O’Donnell CA, Atherton H, Blane DN, Chew-Graham CA. Finding the “right” GP: a qualitative study of the experiences of people with long-COVID. BJGP Open [Internet]. 2020 Dec 1 [cited 2022 Mar 3];4(5):1–12. Available from: https://bjgpopen.org/content/4/5/bjgpopen20X101143

3. Taquet M, Geddes JR, Husain M, Luciano S, Harrison PJ. 6-month neurological and psychiatric outcomes in 236 379 survivors of COVID-19: a retrospective cohort study using electronic health records. The Lancet Psychiatry [Internet]. 2021 May 1 [cited 2022 Mar 6];8(5):416–27. Available from: http://www.thelancet.com/article/S2215036621000845/fulltext

4. Williamson EJ, Walker AJ, Bhaskaran K, Bacon S, Bates C, Morton CE, et al. Factors associated with COVID-19-related death using OpenSAFELY. Nature 2020 584:7821 [Internet]. 2020 Jul 8 [cited 2022 Mar 3];584(7821):430–6. Available from: https://www.nature.com/articles/s41586-020-2521-4

5. Goërtz YMJ, Herck M van, Delbressine JM, Vaes AW, Meys R, Machado FVC, et al. Persistent symptoms 3 months after a SARS-CoV-2 infection: the post-COVID-19 syndrome? ERJ Open Research [Internet]. 2020 Oct [cited 2022 Mar 3];6(4):00542–2020. Available from: /pmc/articles/PMC7491255/

6. Whittaker HR, Gulea C, Koteci A, Kallis C, Morgan AD, Iwundu C, et al. GP consultation rates for sequelae after acute covid-19 in patients managed in the community or hospital in the UK: population based study. BMJ [Internet]. 2021 Dec 29 [cited 2022 Mar 3];375. Available from: https://www.bmj.com/content/375/bmj-2021-065834

7. Danaei G, García Rodríguez LA, Cantero OF, Logan RW, Hernán MA. Electronic medical records can be used to emulate target trials of sustained treatment strategies. Journal of Clinical Epidemiology [Internet]. 2018 Apr 1 [cited 2022 Mar 3];96:12–22. Available from: http://www.jclinepi.com/article/S0895435617304997/fulltext

8. Hernán MA, Robins JM. Using Big Data to Emulate a Target Trial When a Randomized Trial Is Not Available. American Journal of Epidemiology [Internet]. 2016 Apr 15 [cited 2022 Mar 3];183(8):758–64. Available from: https://academic.oup.com/aje/article/183/8/758/1739860

9. Jones KH, Ford D v., Thompson S, Lyons RA. A Profile of the SAIL Databank on the UK Secure Research Platform. International Journal of Population Data Science [Internet]. 2019 Nov 20 [cited 2022 Mar 3];4(2). Available from: https://ijpds.org/article/view/1134

10. NHS Digital. Read Codes [Internet]. [cited 2022 Feb 16]. Available from: https://digital.nhs.uk/services/terminology-and-classifications/read-codes

11. IBM. Eclipse [Internet]. [cited 2022 Mar 3]. Available from: https://www.ibm.com/support/pages/java-sdk-downloads-eclipse

12. RStudio Team. RStudio | Open source & professional software for data science teams - RStudio [Internet]. Boston; 2021 [cited 2022 Mar 3]. Available from: https://www.rstudio.com/

13. Kassambara A, Kosinski M, Biecek P. survminer: Drawing Survival Curves using “ggplot2” [Internet]. 2021 [cited 2022 Mar 3]. Available from: https://cran.r-project.org/web/packages/survminer/index.html

14. Therneau T. A Package for Survival Analysis in R [Internet]. 2021 [cited 2022 Mar 3]. Available from: https://cran.r-project.org/web/packages/survival/index.html

15. Noah Greifer. cobalt: Covariate Balance Tables and Plots. 2022.

16. Rivera-Izquierdo M, Láinez-Ramos-Bossini AJ, de Alba IG-F, Ortiz-González-Serna R, Serrano-Ortiz Á, Fernández-Martínez NF, et al. Long COVID 12 months after discharge: persistent symptoms in patients hospitalised due to COVID-19 and patients hospitalised due to other causes-a multicentre cohort study. BMC medicine [Internet]. 2022 Dec [cited 2022 Mar 6];20(1). Available from: https://pubmed.ncbi.nlm.nih.gov/35193574/

17. Xie Y, Xu E, Bowe B, Al-Aly Z. Long-term cardiovascular outcomes of COVID-19. Nature Medicine 2022 [Internet]. 2022 Feb 7 [cited 2022 Mar 3];1–8. Available from: https://www.nature.com/articles/s41591-022-01689-3

18. Xie Y, Xu E, Al-Aly Z. Risks of mental health outcomes in people with covid-19: cohort study. BMJ [Internet]. 2022 Feb 16 [cited 2022 Mar 6];376:e068993. Available from: https://www.bmj.com/content/376/bmj-2021-068993

19. Rodgers SE, Demmler JC, Dsilva R, Lyons RA. Protecting health data privacy while using residence-based environment and demographic data. Health & Place. 2012 Mar 1;18(2):209–17.

20. Lyons RA, Ford D v., Moore L, Rodgers SE. Use of data linkage to measure the population health effect of non-health-care interventions. The Lancet. 2014 Apr 26;383(9927):1517–9.

